# Waning effectiveness of BNT162b2 and ChAdOx1 COVID-19 vaccines over six months since second dose: a cohort study using linked electronic health records

**DOI:** 10.1101/2022.03.23.22272804

**Authors:** Elsie MF Horne, William J Hulme, Ruth H Keogh, Tom M Palmer, Elizabeth J Williamson, Edward PK Parker, Amelia Green, Venexia Walker, Alex J Walker, Helen Curtis, Louis Fisher, Brian MacKenna, Richard Croker, Lisa Hopcroft, Robin Y Park, Jon Massey, Jessica Morley, Amir Mehrkar, Sebastian Bacon, David Evans, Peter Inglesby, Caroline E Morton, George Hickman, Simon Davy, Tom Ward, Iain Dillingham, Ben Goldacre, Miguel A Hernán, Jonathan AC Sterne

## Abstract

**Background:** The rate at which COVID-19 vaccine effectiveness wanes over time is crucial for vaccination policies, but is incompletely understood with conflicting results from different studies.

**Methods:** This cohort study, using the OpenSAFELY-TPP database and approved by NHS England, included individuals without prior SARS-CoV-2 infection assigned to vaccines priority groups 2-12 defined by the UK Joint Committee on Vaccination and Immunisation. We compared individuals who had received two doses of BNT162b2 or ChAdOx1 with unvaccinated individuals during six 4-week comparison periods, separately for subgroups aged 65+ years; 16-64 years and clinically vulnerable; 40-64 years and 18-39 years. We used Cox regression, stratified by first dose eligibility and geographical region and controlled for calendar time, to estimate adjusted hazard ratios (aHRs) comparing vaccinated with unvaccinated individuals, and quantified waning vaccine effectiveness as ratios of aHRs per-4-week period. The outcomes were COVID-19 hospitalisation, COVID-19 death, positive SARS-CoV-2 test, and non-COVID-19 death.

**Findings:** The BNT162b2, ChAdOx1 and unvaccinated groups comprised 1,773,970, 2,961,011 and 2,433,988 individuals, respectively. Waning of vaccine effectiveness was similar across outcomes and vaccine brands: e.g. in the 65+ years subgroup ratios of aHRs versus unvaccinated for COVID-19 hospitalisation, COVID-19 death and positive SARS-CoV-2 test ranged from 1.23 (95% CI 1.15-1.32) to 1.27 (1.20-1.34) for BNT162b2 and 1.16 (0.98-1.37) to 1.20 (1.14-1.27) for ChAdOx1. Despite waning, rates of COVID-19 hospitalisation and COVID-19 death were substantially lower among vaccinated individuals compared to unvaccinated individuals up to 26 weeks after second dose, with estimated aHRs <0.20 (>80% vaccine effectiveness) for BNT162b2, and <0.26 (>74%) for ChAdOx1. By weeks 23-26, rates of SARS-CoV-2 infection in fully vaccinated individuals were similar to or higher than those in unvaccinated individuals: aHRs ranged from 0.85 (0.78-0.92) to 1.53 (1.07-2.18) for BNT162b2, and 1.21 (1.13-1.30) to 1.99 (1.94-2.05) for ChAdOx1.

**Interpretation:** The rate at which estimated vaccine effectiveness waned was strikingly consistent for COVID-19 hospitalisation, COVID-19 death and positive SARS-CoV-2 test, and similar across subgroups defined by age and clinical vulnerability. If sustained to outcomes of infection with the Omicron variant and to booster vaccination, these findings will facilitate scheduling of booster vaccination doses.

## Introduction

Effectiveness of COVID-19 vaccines, first demonstrated in randomised trials,^1,2^ has been confirmed with longer follow-up in observational studies.^3–5^ However, neutralising antibody titres decrease with time since vaccination,^6–8^ and vaccine effectiveness against infection wanes over time.^5,9–16^ The extent of waning vaccine effectiveness against severe COVID-19 is less clear: studies have found no evidence of,^9,10,17^ modest,^5,11^ or substantial^12^ waning. Clarification of rates of waning effectiveness is needed to determine the frequency with which booster doses are needed and whether booster vaccination should be targeted at groups defined by age, clinical vulnerability or brand of primary vaccination.

Examination of waning COVID-19 vaccine effectiveness is difficult. The success of vaccine rollouts in many countries means that only a small and selected proportion of the population remains unvaccinated. Continuing uptake of vaccination further depletes this unvaccinated group over time. Vaccines were offered in priority order determined by age and clinical vulnerability, so that the longest follow up is in people at highest risk of severe COVID-19. Rapid changes in rates of SARS-CoV-2 infection over time, related to pandemic control measures and introduction of new variants, make it essential to account for the calendar date on which events occurred.

Many studies of waning COVID-19 vaccine effectiveness have used “test-negative case-control” (TNCC) designs, restricted to people tested for infection with SARS-CoV-2 and comparing those testing positive (cases) and negative (controls),^5,9,17,18^ or reported indirect evidence such as changing rates of COVID-19 with time since vaccination.^11,12^ The extent to which TNCC designs control bias due to confounding, or are biased because of the restriction to people who were tested, remains unclear.^19,20^

We conducted a cohort study within the OpenSAFELY-TPP database (https://opensafely.org), which includes detailed linked data on 24 million people registered with an English general practice (GP) using TPP SystmOne electronic health record (EHR) software. We compared rates of COVID-19 hospitalisation, COVID-19 and non-COVID-19 mortality, and infection with SARS-CoV-2, between adults fully vaccinated with the Pfizer-BioNTech BNT162b2 mRNA vaccine (BNT162b2) and the Oxford-AstraZeneca ChAdOx1 nCoV-19 AZD1222 (ChAdOx1), and those who were unvaccinated.

## Methods

### Data source

OpenSAFELY-TPP includes detailed pseudonymized primary care data linked (via National Health Service (NHS) number) with accident and emergency attendance, inpatient hospital spell records (NHS Digital’s Hospital Episode Statistics dataset), national SARS-CoV-2 testing records (Second Generation Surveillance System), and national death registry records. Vaccination status (National Immunisation Management System (NIMS)) is available in the primary care record. Healthcare worker status (recorded for vaccine recipients at the time of vaccination) is provided by NHS Digital’s COVID-19 data store.

### Study design

Individuals eligible for this cohort study, which was approved by NHS England, were adults who had been assigned to UK Joint Committee on Vaccination and Immunisation (JCVI) priority groups 2 to 12 (Supplementary Table 1) and registered with a primary care GP for ≥1 year before eligibility for their first vaccine dose (the “eligibility date”, based on JCVI group and age, Supplementary Table 2). Individuals were assigned to JCVI groups based on information in their linked EHR. They were excluded if their sex, geographical region, ethnicity or index of multiple deprivation were unknown; or they were resident in a care home at six weeks after their eligibility date. Full details are in Supplementary Figure 1.

We defined three groups who: (1) received two doses of BNT162b2; (2) received two doses of ChAdOx1; (3) were unvaccinated. Eligibility for the vaccinated groups was restricted to those who received their second vaccine dose during a 4-week “second vaccination period” (SVP) within analysis strata defined by JCVI group, eligibility date (for groups within which eligibility was based on age (Supplementary Table 2)), and English NHS region, defined using individuals’ GP address). The SVP was defined as the 28-day period during which the greatest number of individuals in the stratum received their second dose. Individuals were excluded from the vaccine groups if they: received their first dose before their eligibility date; had an interval between first and second dose of <6 or >14 weeks; or were flagged as a healthcare worker on their vaccination record. Individuals were assigned to the unvaccinated group if they had received no COVID-19 vaccine at the start of the SVP for their analysis stratum. Individuals were excluded from any group if they had: evidence of previous SARS-CoV-2 infection by the start of their SVP; ever been recorded as being resident in a care home; or evidence of having started an end-of-life care pathway.

Figure 1 depicts the study design. The analysis timescale was calendar time, which ensured that vaccinated and unvaccinated individuals were compared on the calendar day on which each outcome event occurred. Follow-up time for fully vaccinated individuals was split into six consecutive 4-week “comparison periods”, starting 2 weeks after receipt of second dose. Because each SVP was 4 weeks long and each vaccinated individual was followed up for 4 weeks per comparison period, there were 8 calendar weeks during which vaccinated individuals were followed in each comparison period. Vaccinated individuals entered and finished follow-up on the calendar dates corresponding to the start and end of their comparison period.

**Figure 1.**
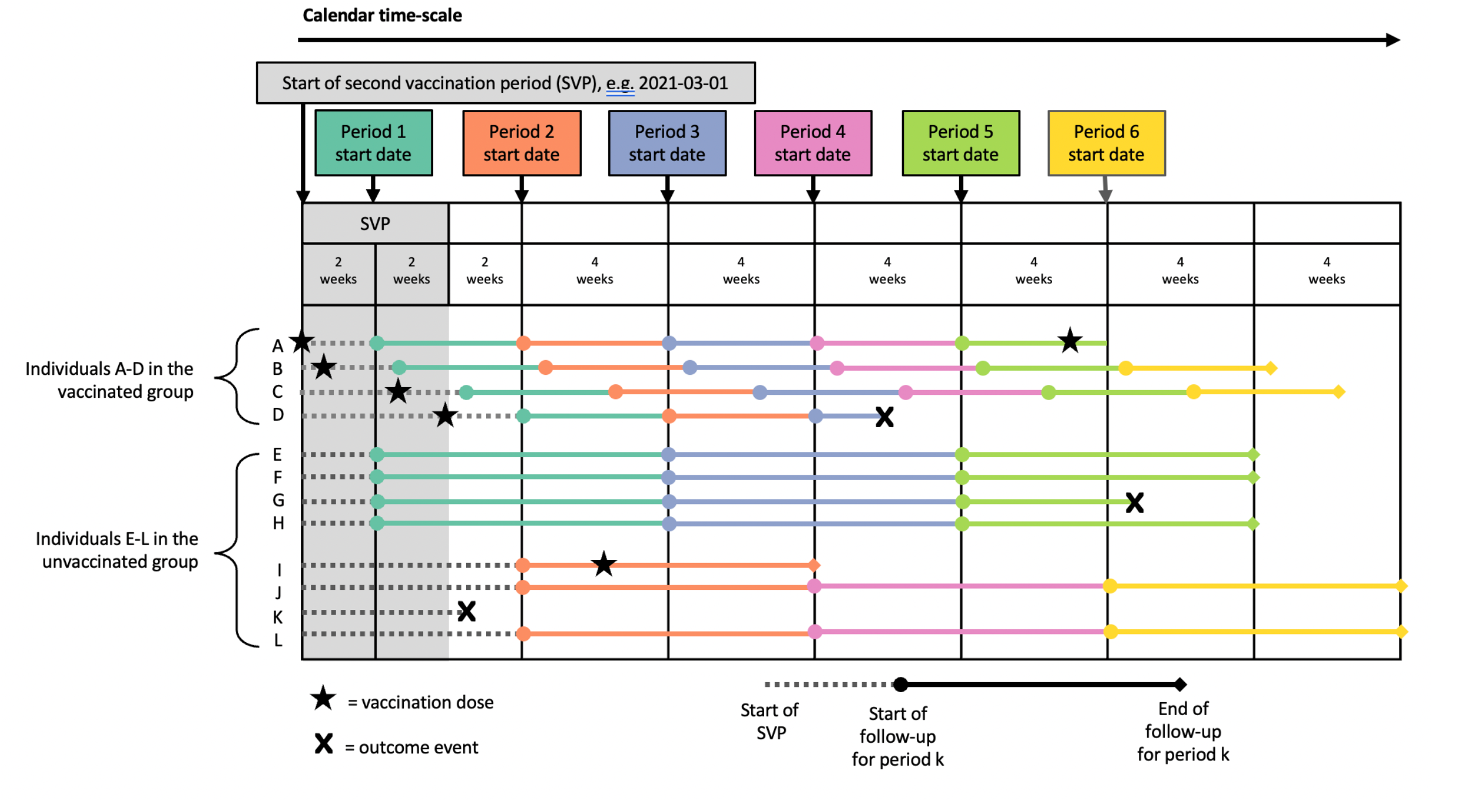
Illustrative example showing the definition of the 4-week comparison periods, within a stratum defined by JCVI group, eligibility date and region. The horizontal lines represent follow-up time for four vaccinated and eight unvaccinated individuals, and the colours of the lines correspond to the six comparison periods. Individuals A and D received their second vaccinations on the first and last day of the second vaccination period (SVP), respectively.

To avoid overlap in follow-up of unvaccinated individuals between comparison periods, follow up time for unvaccinated individuals was assigned at random to start follow up either 2 or 6 weeks after the start of the SVP, and split into the three consecutive 8-week calendar periods during which vaccinated individuals were followed in each comparison period. Unvaccinated individuals assigned to start at 2 weeks were followed during comparison periods 1, 3 and 5, while those assigned to at 6 weeks were followed during comparison periods 2, 4 and 6.

### Outcomes

The outcomes were COVID-19 hospitalisation (identified using HES inpatient hospital records), COVID-19 death, positive SARS-CoV-2 test, and non-COVID-19 death. We also investigated test-seeking behaviour by comparing rates of testing for SARS-CoV-2 between the vaccine groups. COVID-19 and non-COVID-19 deaths (death certificates with and without a COVID-19 code) were based on death registry data from the Office for National Statistics. SARS-CoV-2 tests were identified using SGSS records and based on swab date. Both polymerase chain reaction (PCR) and lateral flow tests were included, without differentiation between symptomatic and asymptomatic infection. All outcomes were defined by the date of their first occurrence during the comparison follow-up period. Where there was no positive SARS-CoV-2 test, but a record of COVID-19 hospitalisation and/or COVID-19 death, the date of positive SARS-CoV-2 test was imputed as the date of COVID-19 hospitalisation or date of COVID-19 death.

### Potential confounding factors

The following potential confounders were defined at a single time (typically the day before the start of comparison period 1; full details in Supplementary Table 3): age; sex (male or female); English Index of Multiple Deprivation (IMD, grouped by quintiles); ethnicity (Black, Mixed, South Asian, White, Other, as per the UK census); number of SARS-CoV-2 tests between 18 May 2020 (when widespread testing became available) and eligibility for first vaccine dose; and receipt of one or more flu vaccines in the five years prior to COVID-19 vaccine eligibility. The other potential confounders were updated at the start of each comparison period: body mass index (BMI); current pregnancy; care home residence; housebound; shielding; history of chronic heart disease, kidney disease, liver disease, neurological disease or respiratory disease; history of diabetes, learning disability, serious mental illness; current immunosuppression or history of permanent immunosuppression; and number of conditions in the clinically “at risk” classification (according to national prioritisation guidelines). After exclusions for missing values in demographic variables, there were no missing values in the remaining variables as they were defined as presence or absence of codes in the EHR.

### Statistical analysis

For each comparison period we estimated hazard ratios (HRs) comparing: (1) BNT162b2 vs unvaccinated; (2) ChAdOx1 vs unvaccinated; (3) BNT162b2 vs ChAdOx1. HRs for comparison periods with <3 events in either group were not estimated. For each individual, follow-up ended at the earliest of the outcome of interest, deregistration from the GP, death, or 15^th^ December 2021. However, for SARS-CoV-2 test, follow-up ended at the “outcome of interest” only when the test result was positive. Fully vaccinated individuals who received a booster dose, and unvaccinated individuals who received a first dose, were excluded from subsequent comparison periods, but follow-up within comparison periods was not censored after these events.

To estimate hazard ratios, we fitted Cox regression models with baseline hazards stratified by JCVI group, eligibility date and region used to define the SVPs, and with the covariates described above. To avoid issues with model convergence, binary covariates were excluded from the model if there were <3 events in any cell of the table defined by cross tabulating the covariate with vaccine group and comparison period. For categorical covariates with more than two levels, levels were merged until either all levels had greater than three events, or there was only one level, in which case the variable was excluded. This process was carried out independently for each outcome. Age within strata was modelled as linear, with quadratic terms additionally included for strata with age range >5 years. We used meta-regression to quantify waning effectiveness as estimated ratios of HRs per comparison period.

All analyses were done independently in four vaccine priority subgroups: (A) aged ≥65 years and in JCVI groups 2-5 (“65+”); (B) aged 16-64 years and clinically vulnerable (JCVI groups 4 or 6; “16-64 CV”); (C) aged 40-64 years (JCVI groups 7-10; most people in this subgroup received ChAdOx1; “40-64”) and (D) aged 18-39 years (JCVI groups 11-12; this subgroup only received BNT162b2; “18-39”). The 65+ subgroup included individuals who were clinically vulnerable, while the 40-64 and 18-39 subgroups did not.

This study followed STROBE-RECORD reporting guidelines. Any counts below six were redacted or rounded for disclosure control. The funders had no role in the study design, collection, analysis, and interpretation of data; in the writing of the report; and in the decision to submit the article for publication.

## Results

Of 13,923,580 individuals satisfying initial eligibility criteria (Supplementary Figure 2), 4,780,020 received second doses of BNT162b2 or ChAdOx1 during the SVP for their stratum (Supplementary Figures 3-17) and 2,596,920 were unvaccinated at the start of their SVP. Of these, 1,773,970, 2,961,011 and 2,433,988 were included in the first comparison period BNT162b2, ChAdOx1 and unvaccinated groups respectively. Table 1 and Supplementary Table 4 show summary statistics for these three groups by subgroup. Compared to vaccinated individuals, unvaccinated individuals were less likely to be white; live in a more affluent area; have had a flu vaccine in the previous five years or have tested for SARS-CoV-2 before their eligibility date. The distribution of other characteristics between vaccine groups differed according to subgroup. For example, in the 65+ subgroup, those vaccinated with BNT162b2 were older than those vaccinated with ChAdOx1 (74 years [IQR 11] versus 71 years [IQR 5], while the converse was true in the 40-64 subgroup (44 years [IQR 9] versus 55 years [IQR 9])

**Table 1.** Summary statistics for selected characteristics across subgroups and vaccination groups

|  |  | ≥ 65 years |  |  | 16-64 years and clinically vulnerable |  |  | 40-64 years* |  |  | 18-39 years* |  |
| --- | --- | --- | --- | --- | --- | --- | --- | --- | --- | --- | --- | --- |
| Characteristic |  | BNT162b2 | ChAdOx1 | Unvaccinated | BNT162b2 | ChAdOx1 | Unvaccinated | BNT162b2 | ChAdOx1 | Unvaccinated | BNT162b2 | Unvaccinated |
| Total |  | 653,435 | 841,239 | 152,294 | 374,137 | 651,574 | 310,445 | 62,652 | 1,463,607 | 623,998 | 683,746 | 1,347,251 |
| Age | Median (IQR) | 74 (11) | 71 (5) | 72 (10) | 54 (15) | 52 (17) | 40 (20) | 44 (9) | 55 (9) | 48 (11) | 30 (11) | 30 (9) |
| Sex | Female | 347,806 (53%) | 435,501 (52%) | 82,283 (54%) | 182,013 (49%) | 331,328 (51%) | 159,164 (51%) | 31,797 (51%) | 687,844 (47%) | 257,530 (41%) | 334,338 (49%) | 605,337 (45%) |
|  | Male | 305,629 (47%) | 405,738 (48%) | 70,011 (46%) | 192,124 (51%) | 320,246 (49%) | 151,281 (49%) | 30,855 (49%) | 775,763 (53%) | 366,468 (59%) | 349,408 (51%) | 741,914 (55%) |
| IMD (1 is most deprived) | 1 | 83,667 (13%) | 110,976 (13%) | 37,216 (24%) | 82,809 (22%) | 147,215 (23%) | 116,493 (38%) | 9,239 (15%) | 190,626 (13%) | 176,288 (28%) | 112,749 (16%) | 425,972 (32%) |
|  | 2 | 109,349 (17%) | 146,207 (17%) | 34,104 (22%) | 76,936 (21%) | 137,960 (21%) | 74,883 (24%) | 11,247 (18%) | 254,639 (17%) | 147,847 (24%) | 139,567 (20%) | 336,285 (25%) |
|  | 3 | 143,949 (22%) | 191,476 (23%) | 32,185 (21%) | 80,052 (21%) | 137,458 (21%) | 54,955 (18%) | 13,453 (21%) | 324,757 (22%) | 127,699 (20%) | 153,740 (22%) | 265,524 (20%) |
|  | 4 | 156,352 (24%) | 196,207 (23%) | 27,553 (18%) | 71,761 (19%) | 120,773 (19%) | 38,540 (12%) | 14,362 (23%) | 342,778 (23%) | 101,526 (16%) | 144,067 (21%) | 192,439 (14%) |
|  | 5 | 160,118 (25%) | 196,373 (23%) | 21,236 (14%) | 62,579 (17%) | 108,168 (17%) | 25,574 (8%) | 14,351 (23%) | 350,807 (24%) | 70,638 (11%) | 133,623 (20%) | 127,031 (9%) |
| Ethnicity | White | 629,163 (96%) | 809,144 (96%) | 120,320 (79%) | 339,971 (91%) | 577,192 (89%) | 223,741 (72%) | 53,182 (85%) | 1,353,356 (92%) | 458,941 (74%) | 590,465 (86%) | 965,781 (72%) |
|  | Black | 3,327 (1%) | 4,051 (0%) | 8,683 (6%) | 5,432 (1%) | 12,204 (2%) | 24,709 (8%) | 1,153 (2%) | 18,105 (1%) | 42,229 (7%) | 8,933 (1%) | 69,018 (5%) |
|  | South Asian | 14,910 (2%) | 19,812 (2%) | 14,842 (10%) | 20,971 (6%) | 46,262 (7%) | 41,049 (13%) | 5,503 (9%) | 57,161 (4%) | 70,931 (11%) | 54,744 (8%) | 168,380 (12%) |
|  | Mixed | 1,981 (0%) | 2,672 (0%) | 2,696 (2%) | 3,568 (1%) | 6,921 (1%) | 9,785 (3%) | 893 (1%) | 12,150 (1%) | 17,327 (3%) | 11,511 (2%) | 44,563 (3%) |
|  | Other | 4,054 (1%) | 5,560 (1%) | 5,753 (4%) | 4,195 (1%) | 8,995 (1%) | 11,161 (4%) | 1,921 (3%) | 22,835 (2%) | 34,570 (6%) | 18,093 (3%) | 99,509 (7%) |
| BMI (kg/m<sup>2</sup>) | <30 | 476,707 (73%) | 585,656 (70%) | 113,867 (75%) | 193,295 (52%) | 345,661 (53%) | 193,942 (62%) | 50,414 (80%) | 1,125,365 (77%) | 518,164 (83%) | 593,256 (87%) | 1,218,459 (90%) |
|  | 30-34.9 | 121,472 (19%) | 168,102 (20%) | 25,490 (17%) | 77,654 (21%) | 129,559 (20%) | 45,655 (15%) | 9,080 (14%) | 252,974 (17%) | 80,099 (13%) | 63,146 (9%) | 92,905 (7%) |
|  | 35-39.9 | 38,613 (6%) | 59,172 (7%) | 8,673 (6%) | 40,365 (11%) | 66,161 (10%) | 21,492 (7%) | 2,981 (5%) | 81,552 (6%) | 24,479 (4%) | 25,221 (4%) | 32,948 (2%) |
|  | 40+ | 16,643 (3%) | 28,309 (3%) | 4,264 (3%) | 62,823 (17%) | 110,193 (17%) | 49,356 (16%) | 177 (0%) | 3,716 (0%) | 1,256 (0%) | 2,123 (0%) | 2,939 (0%) |
| Morbidity count | 0 | 260,080 (40%) | 410,906 (49%) | 78,325 (51%) | 16,632 (4%) | 33,291 (5%) | 15,121 (5%) | 61,987 (99%) | 1,447,217 (99%) | 617,296 (99%) | 679,441 (99%) | 1,337,471 (99%) |
|  | 1 | 211,422 (32%) | 256,688 (31%) | 42,833 (28%) | 251,713 (67%) | 454,041 (70%) | 243,640 (78%) | 648 (1%) | 15,963 (1%) | 6,530 (1%) | 4,224 (1%) | 9,569 (1%) |
|  | ≥ 2 | 181,933 (28%) | 173,645 (21%) | 31,136 (20%) | 105,792 (28%) | 164,242 (25%) | 51,684 (17%) | 17 (0%) | 427 (0%) | 172 (0%) | 81 (0%) | 211 (0%) |
| Number of SARS-CoV-2 tests** | 0 | 551,722 (84%) | 715,240 (85%) | 136,606 (90%) | 262,334 (70%) | 449,231 (69%) | 236,241 (76%) | 44,433 (71%) | 1,056,234 (72%) | 532,962 (85%) | 366,973 (54%) | 1,014,417 (75%) |
|  | 1 | 66,092 (10%) | 84,845 (10%) | 9,043 (6%) | 70,100 (19%) | 124,905 (19%) | 43,578 (14%) | 11,137 (18%) | 252,000 (17%) | 54,887 (9%) | 144,497 (21%) | 176,505 (13%) |
|  | 2 | 19,413 (3%) | 22,687 (3%) | 2,958 (2%) | 23,318 (6%) | 42,502 (7%) | 14,956 (5%) | 3,475 (6%) | 73,497 (5%) | 16,589 (3%) | 63,261 (9%) | 70,777 (5%) |
|  | ≥ 3 | 16,208 (2%) | 18,467 (2%) | 3,687 (2%) | 18,385 (5%) | 34,936 (5%) | 15,670 (5%) | 3,607 (6%) | 81,876 (6%) | 19,560 (3%) | 109,015 (16%) | 85,552 (6%) |
| Flu vaccine past 5 years | yes | 598,623 (92%) | 747,953 (89%) | 59,585 (39%) | 284,323 (76%) | 459,204 (70%) | 91,427 (29%) | 14,920 (24%) | 585,023 (40%) | 34,172 (5%) | 104,104 (15%) | 107,067 (8%) |
| Shielding | yes | 121,846 (19%) | 116,974 (14%) | 23,208 (15%) | 101,785 (27%) | 196,031 (30%) | 67,509 (22%) | 56 (0%) | 1,183 (0%) | 895 (0%) | 237 (0%) | 1,461 (0%) |

Summary statistics calculated two weeks after date of second dose for vaccinated groups and two weeks after start of second vaccination period for unvaccinated group
\* And not clinically vulnerable
\*\* Number of tests in between 18 May 2020 (when widespread testing became available) and the first eligibility date for the subgroup.

The cumulative incidence of first dose by weeks 23-26 in previously unvaccinated individuals was 17%, 27%, 12% and 13% in the 65+, 16-64 CV, 40-64 and 18-39 subgroups respectively (Supplementary Figure 18). The UK vaccination programme initially offered third doses only 6 months (23-26 weeks) after the second dose,^21^ but among the 18-39 subgroup 35% and 26% of those in the BNT162b2 and ChAdOx1 groups respectively had received a third dose by week 20, because the required time since second dose was reduced to three months in mid-December 2021 due to concerns about the Omicron variant.^22,23^

Unadjusted and adjusted hazard ratios (aHRs) are shown in Supplementary Tables 6 and 7, respectively, and compared in Supplementary Figures 19-24 with aHRs for each covariate shown in Supplementary Tables 9-18. For the models comparing individuals fully vaccinated with BNT162b2 and ChAdOx1 to unvaccinated individuals, the unadjusted and adjusted HRs were generally similar. Where they differed, patterns were outcome-and subgroup-specific. Unadjusted and adjusted HRs were similar for comparisons of BNT162b2 with ChAdOx1.

Figure 2 shows aHRs (with 95% CI) for BNT162b2 or ChAdOx1 vaccination versus no vaccination across the six comparison periods. The slopes of the dashed lines correspond to ratios of aHRs per comparison period (also shown in Supplementary Table 8). The missing aHRs could not be estimated because there were too few events in one or both groups (Supplementary Table 5).

**Figure 2.**
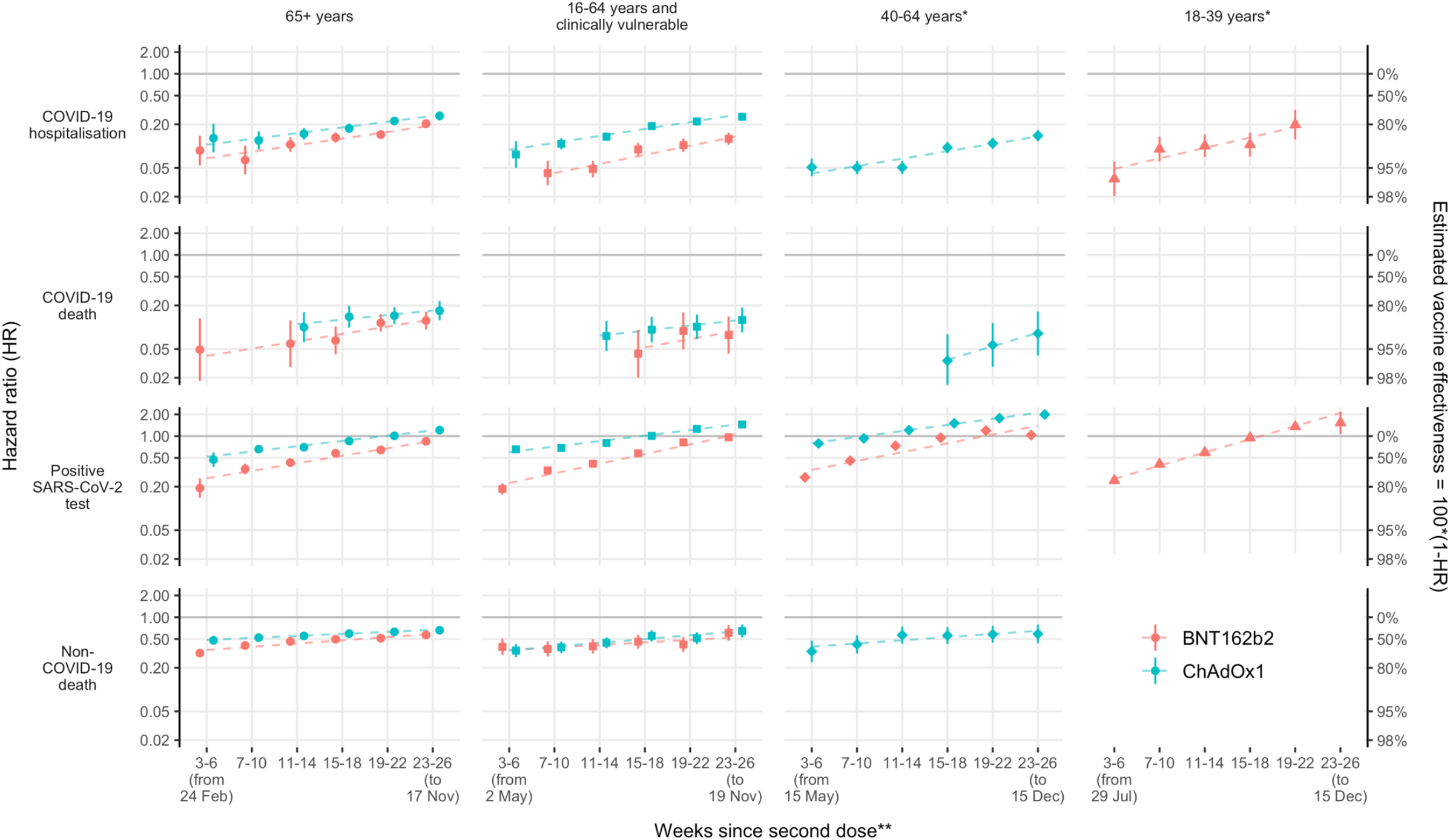
Adjusted hazard ratios comparing BNT162b2 and ChAdOx1 with unvaccinated individuals in each comparison period. Estimates for BNT162b2 in the 40-64 age group are omitted for all outcomes except positive SARS-CoV-2 test due to low event counts. The slopes of the dashed lines are the ratios of hazard ratios across comparison periods, fitted using meta-regression. *And not clinically vulnerable. **All follow-up dates are in 2021.

Follow up in the 65+ subgroup began on 24 February 2021 (when the Alpha variant was dominant) and ended on 17 November 2021. Follow-up for the 16-64 CV, 40-64 and 18-39 subgroups began on 2 May, 15 May and 29 July 2021 respectively: the Delta variant was dominant in England by 1 June 2021. The latest follow up was on 15 December 2021, before the Omicron variant became dominant.

There were 1,731, 4,611 and 9,154 COVID-19 hospitalisations in BNT162b2 recipients, ChAdOx1 recipients and unvaccinated individuals, respectively (Supplementary Table 5). Estimated aHRs comparing BNT162b2 and ChAdOx1 with unvaccinated in the 65+ subgroup were 0.09 (95% CI 0.05-0.14) and 0.13 (0.08-0.20) respectively during weeks 3-6 after second dose, waning to 0.20 (0.18-0.24) and 0.26 (0.23-0.30) respectively during weeks 23-26 (Figure 2). The ratios of aHRs per period were similar for BNT162b2 and ChAdOx1: 1.23 (1.15-1.32) and 1.20 (1.14-1.27) respectively. Estimated aHRs comparing BNT162b2 and ChAdOx1 with unvaccinated in the 16-64 CV subgroup were 0.04 (0.03-0.06) and 0.11 (0.09-0.13) respectively during weeks 7-10, waning to 0.13 (0.10-0.15) and 0.25 (0.22-0.29) respectively by weeks 23-26. The ratios of aHRs per period were 1.34 (1.20-1.48) and 1.25 (1.19-1.30) for BNT162b2 and ChAdOx1 respectively. Estimated aHRs comparing ChAdOx1 with unvaccinated in the 40-64 subgroup waned from 0.05 (0.04-0.07) during weeks 3-6 to 0.14 (0.12-0.17) during weeks 23-26 (ratio of aHRs 1.27 (1.16-1.38) per period). aHRs for BNT162b2 could not be estimated in the 40-46 subgroup because there were too few COVID-19 hospitalisations. Estimated aHRs comparing BNT162b2 with unvaccinated in the 18-39 subgroup waned from 0.04 (0.02-0.06) during weeks 3-6 to 0.20 (0.12-0.32) during weeks 19-22 (ratio of aHRs 1.39 (1.15-1.68) per period).

There were 265, 449, and 831 COVID-19 deaths in BNT162b2 recipients, ChAdOx1 recipients and unvaccinated individuals, respectively (Supplementary Table 5). Estimated aHRs comparing BNT162b2 and ChAdOx1 with unvaccinated in the 65+ subgroup were 0.06 (95% CI 0.03-0.12) and 0.10 (0.06-0.16) respectively during weeks 11-14, waning to 0.20 (0.18-0.24) and 0.26 (0.23-0.30) respectively by weeks 23-26 (Figure 2). The ratios of aHRs per period were 1.26 (1.08-1.48) and 1.16 (.98-1.37) for BNT162b2 and ChAdOx1 respectively. Estimated aHRs comparing BNT162b2 and ChAdOx1 with unvaccinated in the 16-64 CV subgroup were 0.04 (0.02-0.09) and 0.09 (0.06-0.14) during weeks 11-14, waning to 0.08 (0.04-0.14) and 0.13 (0.08-0.19) respectively by weeks 23-26. The ratios of aHRs per period were 1.30 (0.77-2.18) and 1.18 (0.97-1.43) for BNT162b2 and ChAdOx1 respectively. Estimated aHRs comparing ChAdOx1 with unvaccinated in the 40-64 subgroup waned from 0.03 (0.01-0.08) during weeks 11-14 to 0.08 (0.04-0.16) during weeks 23-26. (ratio of aHRs 1.53 (0.89-2.64) per period.

There were 60,171, 180,848 and 118,559 positive SARS-CoV-2 tests in BNT162b2 recipients, ChAdOx1 recipients and unvaccinated individuals, respectively (Supplementary Table 5). For BNT162b2 compared with unvaccinated, estimated aHRs during weeks 3-6 ranged across subgroups from 0.19 (95% CI 0.14-0.26) to 0.27 (0.23-0.31) during weeks 3-6, waning to approximately 1 or greater than 1 by weeks 23-26 (Figure 2, Supplementary Table 7). For ChAdOx1 compared with unvaccinated, estimated aHRs ranged across subgroups from 0.47 (0.38-0.59) to 0.79 (0.76-0.82) and waned to 1.21 (1.13-1.30), 1.45 (1.40-1.50) and 1.99 (1.94-2.05) in the 65+, 16-16 CV and 40-46 subgroups respectively. Rates of waning were similar to those for COVID-19 hospitalisation and COVID-19 death (Supplementary Table 8).

There were 7,318, 7,646, and 3,237 non-COVID-19 deaths in BNT162b2 recipients, ChAdOx1 recipients and unvaccinated individuals, respectively (Supplementary Table 5). Across subgroups, estimated aHRs during weeks 3-6 ranged from 0.32 (95% CI 0.28-0.36) to 0.39 (0.30-0.50) for BNT162b2, and 0.34 (0.24-0.47) to 0.48 (0.42-0.55) for ChAdOx1 (Figure 2, Supplementary Table 7). By weeks 23-26, these ranged from 0.57 (0.51-0.64) to 0.61 (0.47-0.79) for BNT162b2, and 0.59 (0.44-0.79) to 0.66 (0.58-0.76) for ChAdOx1. Rates of waning were lower than for the other outcomes (maximum ratio of aHRs 1.13 (1.07-1.19), Supplementary Table 8).

There were 1,316,223, 2,762,572 and 681,674 SARS-CoV-2 tests in BNT162b2 recipients, ChAdOx1 recipients and unvaccinated individuals, respectively (only the first test in each comparison period counted; Supplementary Table 5). Across subgroups, rates of testing during weeks 3-6 were broadly similar in vaccinated and unvaccinated individuals (aHRs ranged from 0.88 (95% CI 0.87-0.90) to 1.73 (1.69-1.78) for BNT162b2, and from 0.84 (0.82-0.85) to 1.40 (1.39-1.41) for ChAdOx1 (Supplementary Figure 25, Supplementary Tables 7 and 8). By weeks 23-26, rates of testing were substantially higher in vaccinated than unvaccinated individuals; aHRs ranged from 2.99 (2.94-3.05) to 7.05 (6.33-7.84) for BNT162b2 and from 3.80 (3.73-3.87) to 8.95 (8.80-9.09) for ChAdOx1.

Estimated aHRs comparing BNT162b2 with ChAdOx1 recipients consistently favoured BNT162b2 (Figure 3 and Supplementary Table 7, ratios of aHRs in Supplementary Table 8). For the earliest comparison period during which they were estimable, aHRs across subgroups ranged from 0.37 (95% CI 0.24-0.55) to 0.45 (0.29-0.71) for COVID-19 hospitalisation during weeks 7-11; 0.35 (0.21-0.58) to 0.45 (0.19-1.05) for COVID-19 death during weeks 15-18; and 0.35 (0.29-0.41) to 0.62 (0.47-0.81) for positive SARS-CoV-2 test during weeks 3-6. Because the rate of waning was slightly higher for BNT162b2 than ChAdOx1, the aHRs were attenuated by weeks 23-26 (range across subgroups from 0.47 (0.39-0.57) to 0.73 (0.65-0.83) for COVID-19 hospitalisation; 0.59 (0.32-1.07) to 0.70 (0.53-0.94) for COVID-19 death; and 0.60 (0.55-0.67) to 0.76 (0.73-0.79) for positive SARS-CoV-2 test).

**Figure 3.**
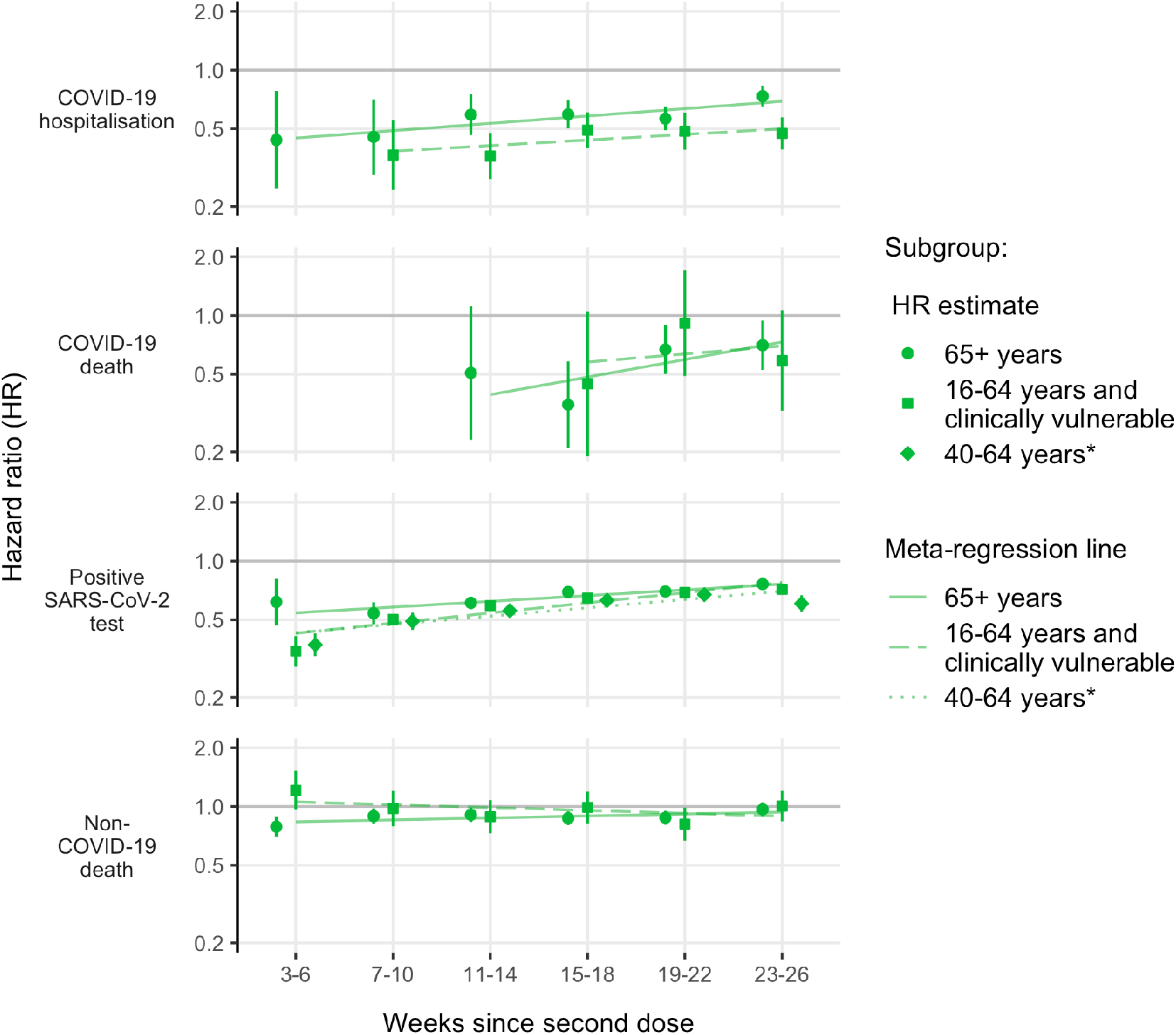
Adjusted hazard ratios comparing BNT162b2 with ChAdOx1. Hazard ratios <1 favour BNT162b2. The slopes of the lines correspond to ratios of hazard ratios across comparison periods, estimated using meta-regression. *And not clinically vulnerable

## Discussion

This cohort study estimated the effectiveness and comparative effectiveness of the BNT162b2 and ChAdOx1 vaccines during six 4-week periods after receipt of second dose. Rates of COVID-19 hospitalisation and COVID-19 death were consistently and substantially lower among fully vaccinated individuals compared to those who remained unvaccinated, up to 26 weeks after second vaccination, and consistently lower among individuals fully vaccinated with BNT162b2 than ChAdOx1. However, by 23-26 weeks rates of SARS-CoV-2 infection (ascertained through freely available national routine testing) in fully vaccinated individuals were similar to or higher than those in unvaccinated individuals. Rates of non-COVID-19 death were consistently lower among fully vaccinated than unvaccinated individuals.

When quantified as ratios of aHRs, the waning of estimated effectiveness was strikingly similar across risk groups, except that estimated effectiveness waned fastest in the 18-39 subgroup (those at lowest risk of severe COVID-19, all vaccinated with BNT162b2). In those subgroups in which the two vaccines could be compared, estimated effectiveness was initially greater for BNT162b2 than for ChAdOx1, but effectiveness waned somewhat faster for BNT162b2 than ChAdOx1, so that the two brands’ comparative effectiveness became more similar over time. Estimated aHRs for COVID-19 hospitalisation and COVID-19 death remained <0.20 (at least 80% vaccine effectiveness) for BNT162b2 and <0.26 (at least 74% vaccine effectiveness) for ChAdOx1 during weeks 23-26 after second vaccination.

A systematic review of the duration of effectiveness of COVID-19 vaccines^13^ included 18 studies, the majority of which evaluated BNT162b2 or Moderna-mRNA-1273 vaccines. Estimated vaccine effectiveness decreased by 10 percentage points (95% CI 6-15) for severe COVID-19 (hospitalisation or death due to COVID-19), and by 21 percentage points (95% CI 14-30) for SARS-CoV-2 infection, from 1 to 6 months after full vaccination. Estimates varied substantially between studies with reductions comparable to those in our study reported by only two studies.^9,14^ The review concluded that the decline in vaccine effectiveness against severe COVID-19 was less than for SARS-CoV-2 infection and symptomatic disease, whereas we found that rates of waning, quantified as ratios of HRs, were similar for both outcomes across the four subgroups and that estimated vaccine effectiveness against COVID-19 hospitalisation and COVID-19 death remained high 23-26 weeks after receipt of the second dose.

As in this study, Andrews and colleagues analysed NHS England EHR data to investigate of the duration of protection by COVID-19 vaccines against symptomatic and severe COVID-19.^5^ They used a TNCC design, restricted to individuals who were tested for SARS-CoV-2 infection. In the 65+ and 40-64 CV subgroups, identically defined in the two studies, estimated vaccine effectiveness against COVID-19 hospitalisation and COVID-19 death was consistently 3-6% lower in this study than that of Andrews and colleagues, who concluded (by contrast with our study) that waning was greater for ChAdOx1 than BNT162b2, and greater among older adults and those in a clinical risk group. While those authors found continuing vaccine effectiveness against symptomatic COVID-19 up to 26 weeks since receipt of the second vaccine dose, we found rates of SARS-COV-2 infection to be similar to or higher than those in unvaccinated individuals by that time.

Our study is based on whole-population data analysed within the OpenSAFELY Trusted Research Environment, which has stringent disclosure controls to protect patient privacy. The large study size and large numbers of outcome events led to precise estimates of vaccine effectiveness according to vaccine brand and time since second vaccine dose. We accounted for risk-dependent vaccine allocation by separating the cohort into subgroups based on JCVI group,^26^ and conducting analyses within strata defined by JCVI group, eligibility date for primary vaccination and geographical region. Our analyses also excluded individuals with a pre-vaccine rollout record of SARS-CoV-2 infection and accounted for rapid changes in COVID-19 incidence with calendar time, censoring due to occurrence of outcome events, and attenuation of comparison groups because receipt of receipt of first vaccine dose by unvaccinated individuals and third dose by fully vaccinated individuals.

Our study has several limitations. First, as in any observational study, our estimates could be affected by confounding by unmeasured factors. However, the detailed linked data analysed permitted adjustment for a wide range of potential confounding factors. Second, patients registered with a GP who have moved or emigrated (or whose death was not recorded)^25^ may contribute person-time but not events. Because the BNT162b2 and ChAdOx1 groups are defined by recent vaccination, these “ghost” patients are more likely to be present in the unvaccinated group, leading to bias in estimates of waning. Also, healthcare workers could be identified and excluded from the vaccinated groups because this information was recorded at the time of vaccination, but not from the unvaccinated group. This limitation should not affect results for the 65+ subgroup, most of whom are retired, or comparisons between BNT162b2 and ChAdOx1. Third, consistent with an Australian survey,^24^ we found that unvaccinated individuals had tested less frequently than vaccinated individuals during the pre-vaccine rollout period when widespread testing was available, and were considerably less likely to be tested during follow-up. Fourth, differential depletion of susceptible people in the unvaccinated groups over time may lead to attenuation of HRs even when true vaccine effectiveness does not change. However, such bias is likely to be minimal when vaccine effectiveness is high.^27^

Our results have immediate implications for COVID-19 vaccination policies. When quantified as ratios of HRs, the rate at which estimated vaccine effectiveness waned was strikingly consistent (there was little variation around the fitted rates of waning displayed in Figure 2) and (by contrast with other studies) similar across subgroups defined by age and clinical vulnerability. If sustained to outcomes of infection with the Omicron variant and to booster vaccination, these findings will facilitate scheduling of booster vaccination doses. By 26 weeks after second dose, rates of infection with SARS-CoV-2 were similar to or higher in fully vaccinated than unvaccinated individuals, implying that vaccination has only transient impacts on transmissibility of SARS-CoV-2. This may arise partly because a desirable consequence of vaccination is to facilitate greater social mixing because of the reduced risk of severe COVID-19. Protection against COVID-19 hospitalisation and COVID-19 death was substantial up to 26 weeks after second vaccination, even in older and clinically vulnerable individuals. Finally, cessation of freely-available population-based testing programmes is likely to limit applications of the TNCC design, which have to date provided rapid estimates of vaccine effectiveness. By contrast, cohort approaches based on detailed linked EHR data, such as were used in this study, should remain feasible for severe COVID-19 outcomes.

## Data Availability

All data were linked, stored and analysed securely within the OpenSAFELY platform: https://opensafely.org/. Data include pseudonymised data such as coded diagnoses, medications and physiological parameters. No free text data are included. All code is shared openly for review and re-use under MIT open license https://github.com/opensafely/covid-ve-change-over-time. Detailed pseudonymised patient data is potentially re-identifiable and therefore not shared. Codelists are available at https://www.opencodelists.org/.

## Contributors

JACS and MAH conceived the study. JACS, MAH, EMFH, WJH, RK, TP and EJW designed the study. EMFH, WJH, SB, ID, BG contributed to data curation. EMFH, WJH, and JACS conducted the statistical analysis. JACS and BG acquired funding for the study. WJH, JM, AM, BG contributed to project administration. SB and ID contributed resources for the study. SB, and ID created and maintained software. SB, DE, PI, CEM, GH, SD, TW, and ID, created and maintained software. JACS, WJH, JM, AM, and BG supervised the study. AM, and BG were responsible for work relating to information governance. EMFH and JACS wrote the first draft of the manuscript. All authors contributed to reviewing and editing of the manuscript. All authors had final responsibility for the decision to submit for publication.

## Data sharing

Data management and analyses were conducted in Python version 3.8.10 and R version 4.0.2. All data were linked, stored and analysed securely within the OpenSAFELY platform: https://opensafely.org/. Data include pseudonymised data such as coded diagnoses, medications and physiological parameters. No free text data are included. All code is shared openly for review and re-use under MIT open license https://github.com/opensafely/covid-ve-change-over-time. Detailed pseudonymised patient data is potentially re-identifiable and therefore not shared. Codelists are available at https://www.opencodelists.org/.

## Acknowledgements

This work was supported by the This work was supported by the Longitudinal Health and Wellbeing COVID-19 National Core Study (UKRI Medical Research Council MC_PC_20030 and MC_PC_20059), Asthma UK, and NIHR grant MR/V015737/1. The OpenSAFELY software platform is funded by Wellcome and by the Data and Connectivity COVID-19 National Core Study, led by Health Data Research UK in partnership with the Office for National Statistics and funded by UK Research and Innovation (grant ref MC_PC_20058). TPP provided technical expertise and infrastructure within their data centre pro bono in the context of a national emergency.

BG’s work on better use of data in healthcare more broadly is currently funded in part by: NIHR Oxford Biomedical Research Centre, NIHR Applied Research Collaboration Oxford and Thames Valley, the Mohn-Westlake Foundation, NHS England, and the Health Foundation; all DataLab staff are supported by BG’s grants on this work. RHK was funded by UK Research and Innovation (Future Leaders Fellowship MR/S017968/1). VW and TMP were supported by the MRC Integrative Epidemiology Unit, which receives funding from the UKRI Medical Research Council and the University of Bristol (MC_UU_00011/1 and MC_UU_00011/3). EPKP was funded by UK Research and Innovation (COVID-19 data analysis secondment MR/W021420/1). EW holds grants from MRC. JACS is supported by the NIHR Bristol Biomedical Research Centre and by Health Data Research UK.

The views expressed are those of the authors and not necessarily those of the NIHR, NHS England, Public Health England or the Department of Health and Social Care.

This research was funded in part by Wellcome. A CC BY or equivalent licence is applied to author accepted manuscript arising from this submission, in accordance with the grant’s open access conditions.

We are very grateful for all the support received from the EMIS and TPP Technical Operations team throughout this work, and for generous assistance from the information governance and database teams at NHS England / NHSX.

## Information governance and ethical approval

NHS England is the data controller for OpenSAFELY-TPP; TPP is the data processor; all study authors using OpenSAFELY have the approval of NHS England. This implementation of OpenSAFELY is hosted within the TPP environment which is accredited to the ISO 27001 information security standard and is NHS IG Toolkit compliant.^28,29^

Patient data has been pseudonymised for analysis and linkage using industry standard cryptographic hashing techniques; all pseudonymised datasets transmitted for linkage onto OpenSAFELY are encrypted; access to the platform is via a virtual private network (VPN) connection, restricted to a small group of researchers; the researchers hold contracts with NHS England and only access the platform to initiate database queries and statistical models; all database activity is logged; only aggregate statistical outputs leave the platform environment following best practice for anonymisation of results such as statistical disclosure control for low cell counts.^30^

The OpenSAFELY research platform adheres to the obligations of the UK General Data Protection Regulation (GDPR) and the Data Protection Act 2018. In March 2020, the Secretary of State for Health and Social Care used powers under the UK Health Service (Control of Patient Information) Regulations 2002 (COPI) to require organisations to process confidential patient information for the purposes of protecting public health, providing healthcare services to the public and monitoring and managing the COVID-19 outbreak and incidents of exposure; this sets aside the requirement for patient consent.^31^

Taken together, these provide the legal bases to link patient datasets on the OpenSAFELY platform. GP practices, from which the primary care data are obtained, are required to share relevant health information to support the public health response to the pandemic, and have been informed of the OpenSAFELY analytics platform.

This study was approved by the Health Research Authority (REC reference 20/LO/0651) and by the LSHTM Ethics Board (reference 21863).

