## Supplementary Material for "Waning effectiveness of BNT162b2 and ChAdOx1 COVID-19 vaccines over six months since second dose: a cohort study using linked electronic health records"

### Contents

**List of Tables** 1

**List of Figures** 1

**Supplementary Methods** 1

**Supplementary Results** 7

9 Hazard ratios corresponding to covariates in the adjusted BNT162b2 vs unvaccinated model in the 65+ years subgroup. 36
10 Hazard ratios corresponding to covariates in the adjusted ChAdOx1 vs unvaccinated model in the 65+ years subgroup. 38
11 Hazard ratios corresponding to covariates in the adjusted BNT162b2 vs ChAdOx1 model in the 65+ years subgroup. 40
12 Hazard ratios corresponding to covariates in the adjusted BNT162b2 vs unvaccinated model in the 16-64 years and clinically vulnerable subgroup. . . . . . 42
13 Hazard ratios corresponding to covariates in the adjusted ChAdOx1 vs unvaccinated model in the 16-64 years and clinically vulnerable subgroup. . . . . . 44
14 Hazard ratios corresponding to covariates in the adjusted BNT162b2 vs ChAdOx1 model in the 16-64 years and clinically vulnerable subgroup. . . . . . 46
15 Hazard ratios corresponding to covariates in the adjusted BNT162b2 vs unvaccinated model in the 40-64 years and not clinically vulnerable subgroup. . . . . . 48
16 Hazard ratios corresponding to covariates in the adjusted ChAdOx1 vs unvaccinated model in the 40-64 years and not clinically vulnerable subgroup. . . . . . 50
17 Hazard ratios corresponding to covariates in the adjusted BNT162b2 vs ChAdOx1 model in the 40-64 years and not clinically vulnerable subgroup. . . . . . 52
18 Hazard ratios corresponding to covariates in the adjusted BNT162b2 vs unvaccinated model in the 18-39 years and not clinically vulnerable subgroup. . . . . . 54

### Supplementary Methods

**Supplementary Table 1:** Priority groups for primary vaccination advised by the Joint Committee on Vaccination and Immunisation

| Priority group | Risk group |
|---|---|
| 1 | Residents in a care home for older adults<br>Staff in a care home for older adults |
| 2 | All those 80 years of age and over<br>Frontline health and social care workers |
| 3 | All those 75 years of age and over |
| 4 | All those 70 years of age and over<br>Individuals aged 16 to 69 in a high risk group[a] |
| 5 | All those 65 years of age and over |
| 6 | Individuals aged 16 to 65 years in an at-risk group[a] |
| 7 | All those 60 years of age and over |
| 8 | All those 55 years of age and over |
| 9 | All those 50 years of age and over |
| 10 | All those 40 years of age and over |
| 11 | All those 30 years of age and over |
| 12 | All those 18 years of age and over |

[a] See COVID-19: the green book, chapter 14a for definitions of the high-risk and at-risk groups.

**Supplementary Table 2:** Eligibility dates for first vaccine dose

| Grouped eligibility date | Exact eligibility date | JCVI groups | Age range | Reference |
|---|---|---|---|---|
| 2020-12-08 | 2020-12-08 | 01, 02, 03 | - | source |
| 2021-01-18 | 2021-01-18 | 04 | - | source |
| 2021-02-15 | 2021-02-15 | 05, 06 | - | source |
| 2021-02-22 | 2021-02-22 | 07 | 64 | source |
| 2021-03-01 | 2021-03-01 | 07 | 60-63 | source |
| 2021-03-08 | 2021-03-08 | 08 | 56-59 | source |
| | 2021-03-09 | 08 | 55 | source |
| 2021-03-19 | 2021-03-19 | 09 | 50-54 | source |
| 2021-04-13 | 2021-04-13 | 10 | 45-49 | source |
| 2021-04-26 | 2021-04-26 | 10 | 44 | source |
| | 2021-04-27 | 10 | 42-43 | source |
| | 2021-04-30 | 10 | 40-41 | source |
| 2021-05-13 | 2021-05-13 | 11 | 38-39 | source |
| | 2021-05-19 | 11 | 36-37 | source |
| 2021-05-21 | 2021-05-21 | 11 | 34-35 | source |
| | 2021-05-25 | 11 | 32-33 | source |
| | 2021-05-26 | 11 | 30-31 | source |
| 2021-06-08 | 2021-06-08 | 12 | 25-29 | source |
| 2021-06-15 | 2021-06-15 | 12 | 23-24 | source |
| | 2021-06-16 | 12 | 21-22 | source |
| | 2021-06-18 | 12 | 18-20 | source |

JCVI: Joint Committee on Vaccination and Immunisation

**Supplementary Table 3:** Variable definitions

| Group | Name | Description | Encoding | Date defined |
|---|---|---|---|---|
| Exclusion criteria | Evidence of prior COVID-19 infection | Date of code corresponding to COVID-19 hospitalisation, positive SARS-CoV-2 test, or probable COVID-19 code in GP record. | Date | First occurence |
| | End-of-life care pathway initiated | Date of code corresponding to end-of-life care or midazolam injection used in end-of-life care in patient's record. | Date | First occurence prior to SVP |
| | Resident in care home | Date of code corresponding to care home in patient's record. | Date | First occurence prior to SVP |
| Strata | JCVI group | Priority groups defined by the JCVI expert advisory group | 2-12 | Age defined on 31 March 2021 for individuals eligible in phase 1 and 1 July 2021 for individuals eligible in phase 2. Individuals defined as clinically extremely vulnerable on 18 Januray 2021, and at-risk on 15 February 2021. |
| | Eligibility date | Date on which JCVI group (or age range within group) became eligible for 1st dose of COVID-19 vaccination. | Date | - |
| | Region | NHS region based on practice address. | East of England; Midlands; London; North East; Yorkshire; North West; South East; South West. | Eligibility date + 6 weeks. |
| Potential confounders (demographic) | Sex | - | M; F. | - |
| | Age | Age in whole years. | Numeric[a] | $start_{1i}$ |
| | Ethnicity | From primary care records or SUS if missing from primary care. | Black; Mixed; South Asian; White; Other. | Eligibility date + 6 weeks |
| | Index of Multiple Deprivation (IMD) | IMD quintile based on patient address | 1; 2; 3; 4; 5. | Eligibility date + 6 weeks |

**Supplementary Table 3:** Variable definitions *(continued)*

| Group | Name | Description | Encoding | Date defined |
|---|---|---|---|---|
| Potential confounders (clinical) | Body Mass Index (BMI) | Based on numeric BMI data from primary care records. | Under 30kg/$m^2$ or not recorded; 30-34.9; 35-39.9; 40+ kg/$m^2$. | $\text{start}_{ki}$ |
| | Chronic heart disease; Chronic respiratory disease; Chronic liver disease; Chronic kidney disease; Diabetes; Chronic neurological disease; Immunosuppressed; Learning disability including Down's syndrome; Serious mental illness. | Any code in primary care record before date defined. | 0; 1. | $\text{start}_{ki}$ |
| | Multimorbidity | Number of comorbid conditions in different organ systems. | 0; 1; 2+ | $\text{start}_{ki}$ |
| | Influenza vaccination | Any record in past 5 years. | 0; 1. | Eligibility date |
| | Clinically extremely vulnerable | UK government COVID-19 shielding criteria met. | 0; 1. | $\text{start}_{ki}$ |
| | Pregnancy | Pregnancy recorded in 36 weeks prior to date and no delivery code recorded more recently that pregnancy code. | 0; 1. | $\text{start}_{ki}$ |
| | Care home | Code corresponding to care home in patient's record before evaluation date. | 0; 1. | $\text{start}_{ki}$ |
| | Housebound | Code indicating that the individual is housebound. | 0; 1. | $\text{start}_{ki}$ |
| | Number of SARS-CoV-2 tests. | SARS-CoV-2 tests were identified using SGSS records and based on swab date. Both polymerase chain reaction (PCR) and lateral flow tests will be included, without differentiation between symptomatic and asymptomatic infection. | Integer | Between 18 May 2020 (when widespread testing became available in England) and the earliest date of eligibility for first dose in the subgroup. |

JCVI: Joint Committee on Vaccination and Immunisation; $\text{start}_{ki}$ is the start date of comparison period $k$ for individual $i$.
[a] Age was modelled separately within each strata. Age within strata was modelled as linear, with quadratic terms additionally included for strata with age range >5 years.

**Box A: Initial eligibility criteria.**
**Inclusion criteria:**
- Registered at a GP surgery (using TPP's SystemOne clinical information system) for at least one year prior to the date at which they became eligible for vaccination.
- Assigned to JCVI groups 2 to 12

**Exclusion criteria:**
- Aged under 16 or over 120 on 31 March 2021
- Sex unknown
- Geographical region, ethnicity or deprivation unknown on eligibility date + 6 weeks.
- Evidence of SARS-CoV-2 infection recorded on or before eligibility date + 6 weeks.
- Recorded as being in care home on or before eligibility date + 6 weeks

**Box B: Eligibility criteria for identification of second vaccination period (SVP).**
**Inclusion criteria:**
- Received second dose of BNT162b2 or ChAdOx1 between eligibility date + 6 weeks and eligibility date + 16 weeks (first and second doses must be same brand).

**Exclusion criteria:**
- First dose received before eligibility date.
- Interval of less than 6 or more than 14 weeks between first and second doses.
- Flagged as healthcare worker on vaccination record.

**Box C: Inclusion criteria for vaccinated group:**
- Second dose of BNT162b2 or ChAdOx1 received during the SVP.

**Box D: Inclusion criteria for unvaccinated group:**
- No doses of vaccine received before the start of the SVP.

**Box E: Exclusion criteria for both groups:**
- Evidence of SARS-CoV-2 infection recorded on or before start of SVP.
- Recorded as being in care home on or before start of the SVP.
- End-of-life care pathway initiated on or before start of the SVP.

**Box F: Individual *i* excluded from comparison period *k* with outcome *z* if any of the following events have occurred on or before the start of their follow-up for comparison period *k* (start_fu_{ki}):**
- Third dose of vaccine received for individuals in the vaccinated group, or first dose of vaccine received for individuals in the unvaccinated group.
- Outcome *z*.*
- Death of any cause.
- De-registration from all practices using using TPP's SystemOne clinical information system.

**Supplementary Figure 1:** Study eligibility criteria.

**Supplementary Results**

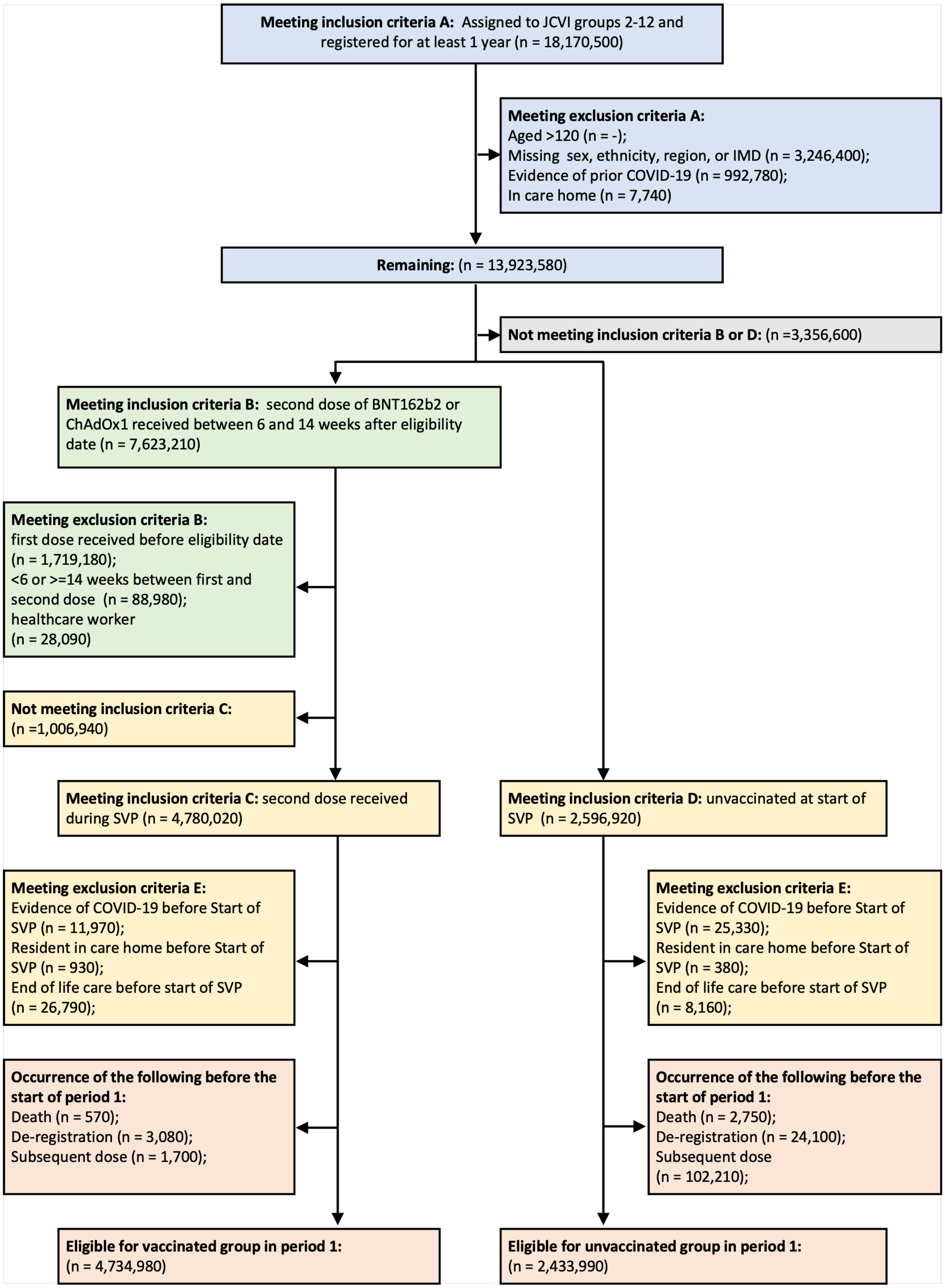

**Supplementary Figure 2:** Flow of individuals into study

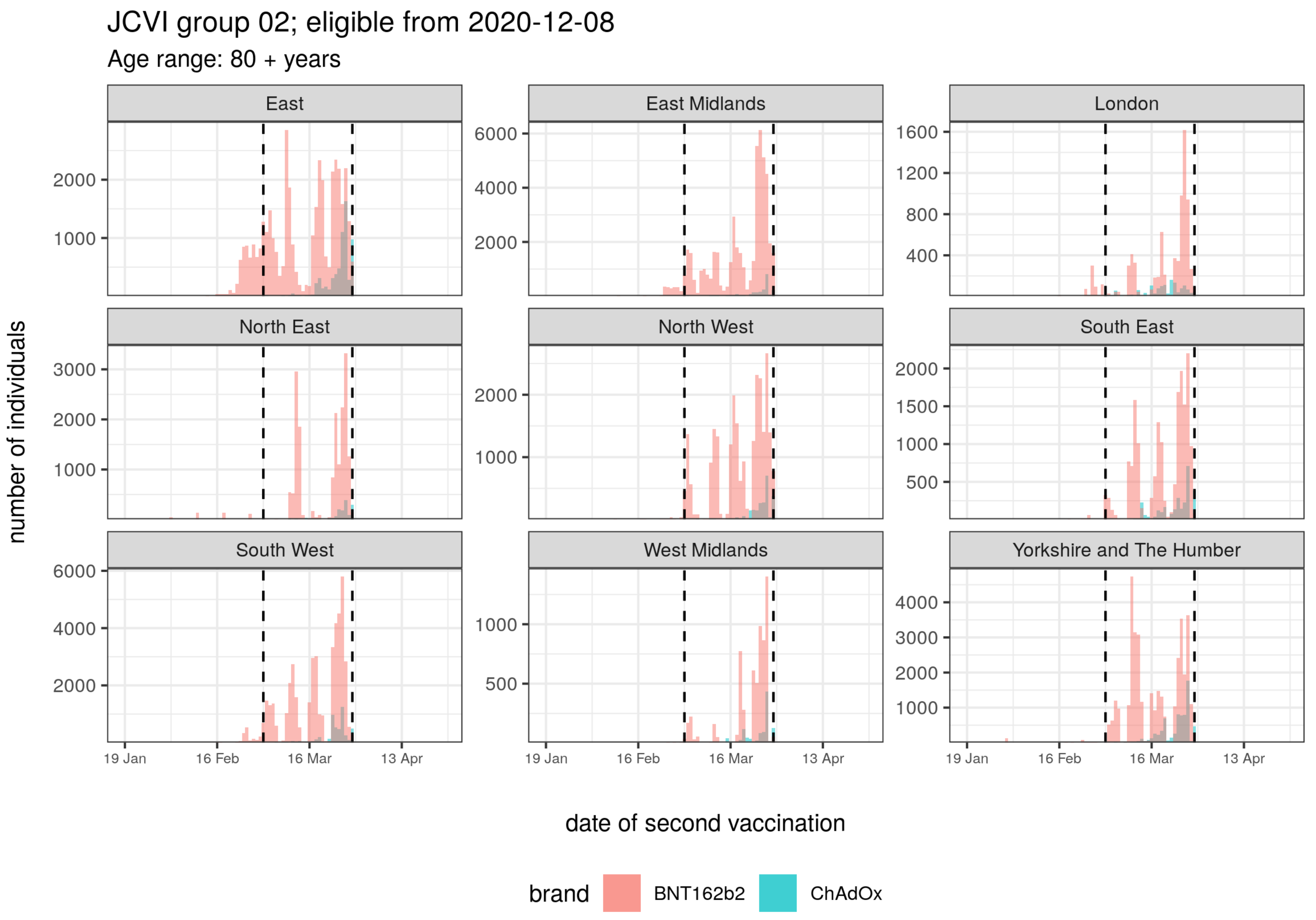

**Supplementary Figure 3:** Second vaccination period for JCVI group 2

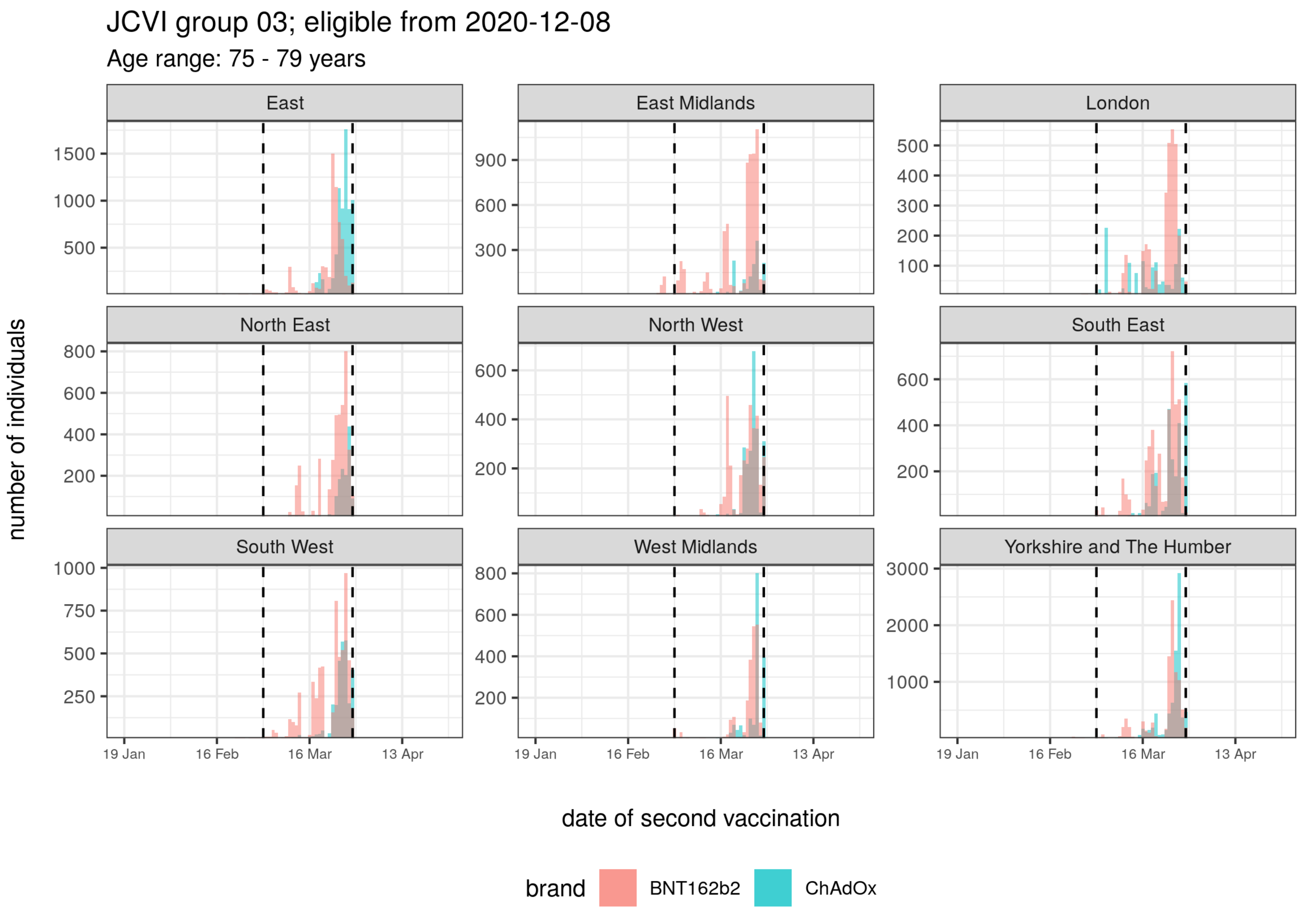

**Supplementary Figure 4:** Second vaccination period for JCVI group 3

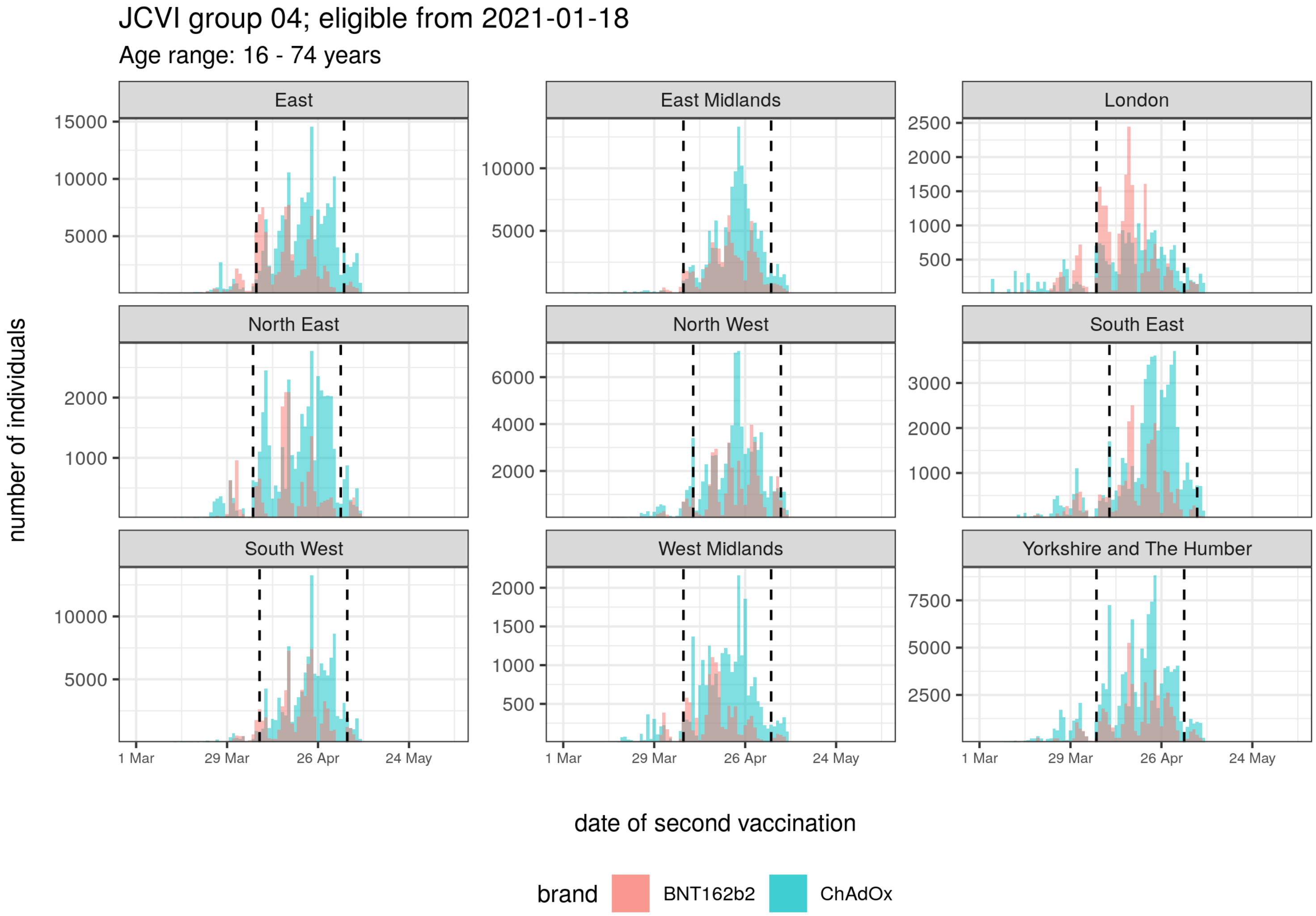

**Supplementary Figure 5:** Second vaccination period for JCVI group 4

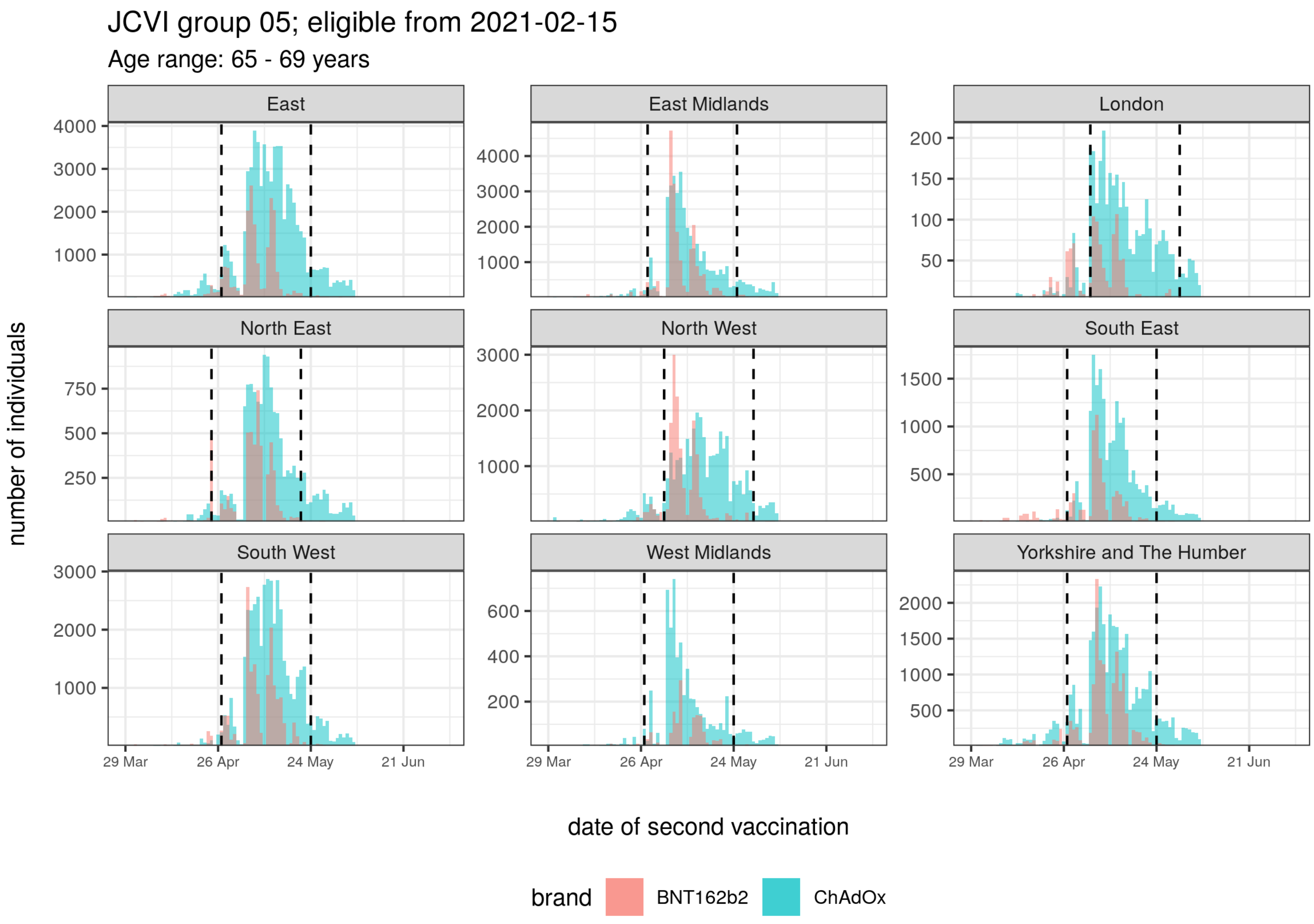

**Supplementary Figure 6:** Second vaccination period for JCVI group 5

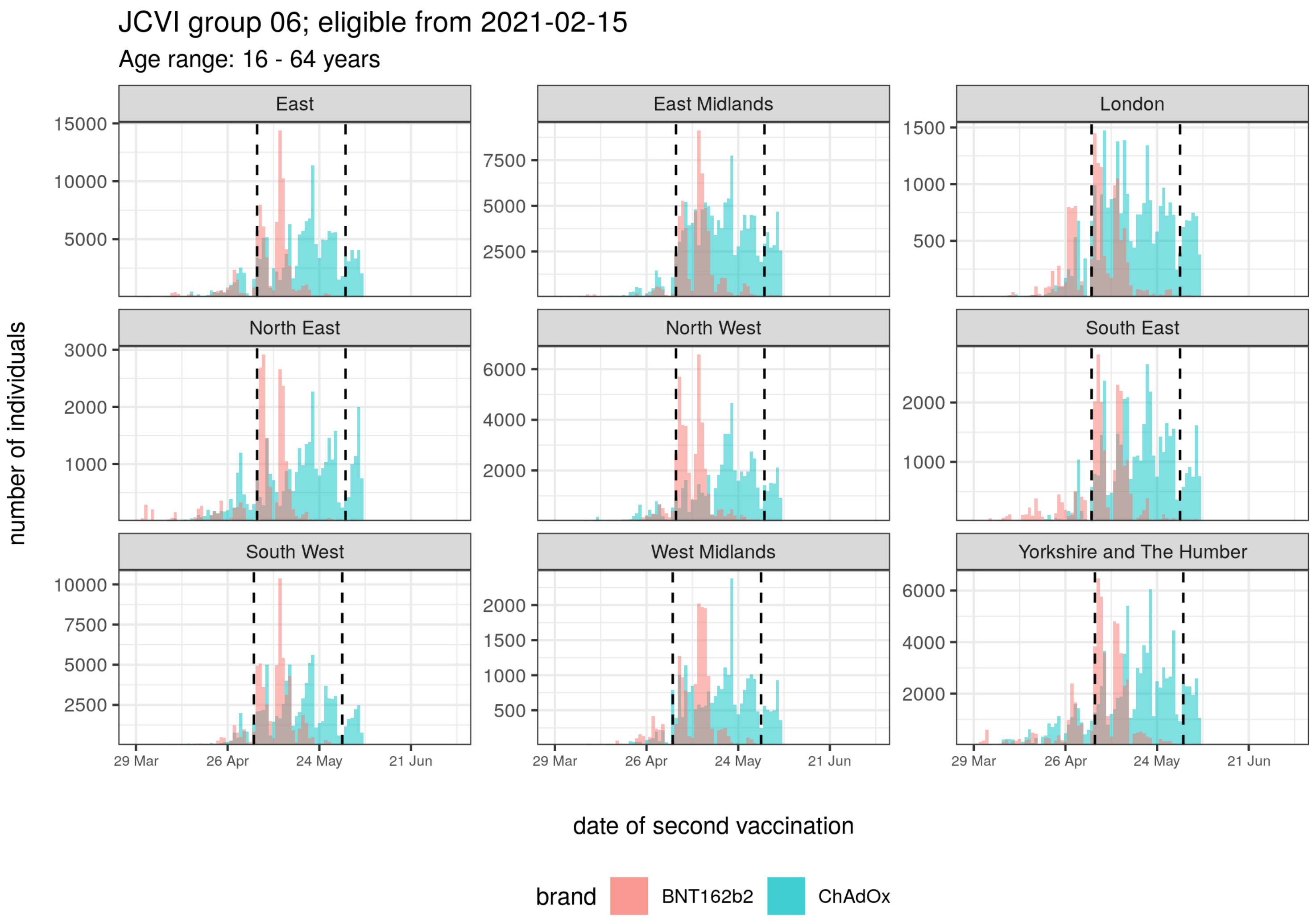

**Supplementary Figure 7:** Second vaccination period for JCVI group 6

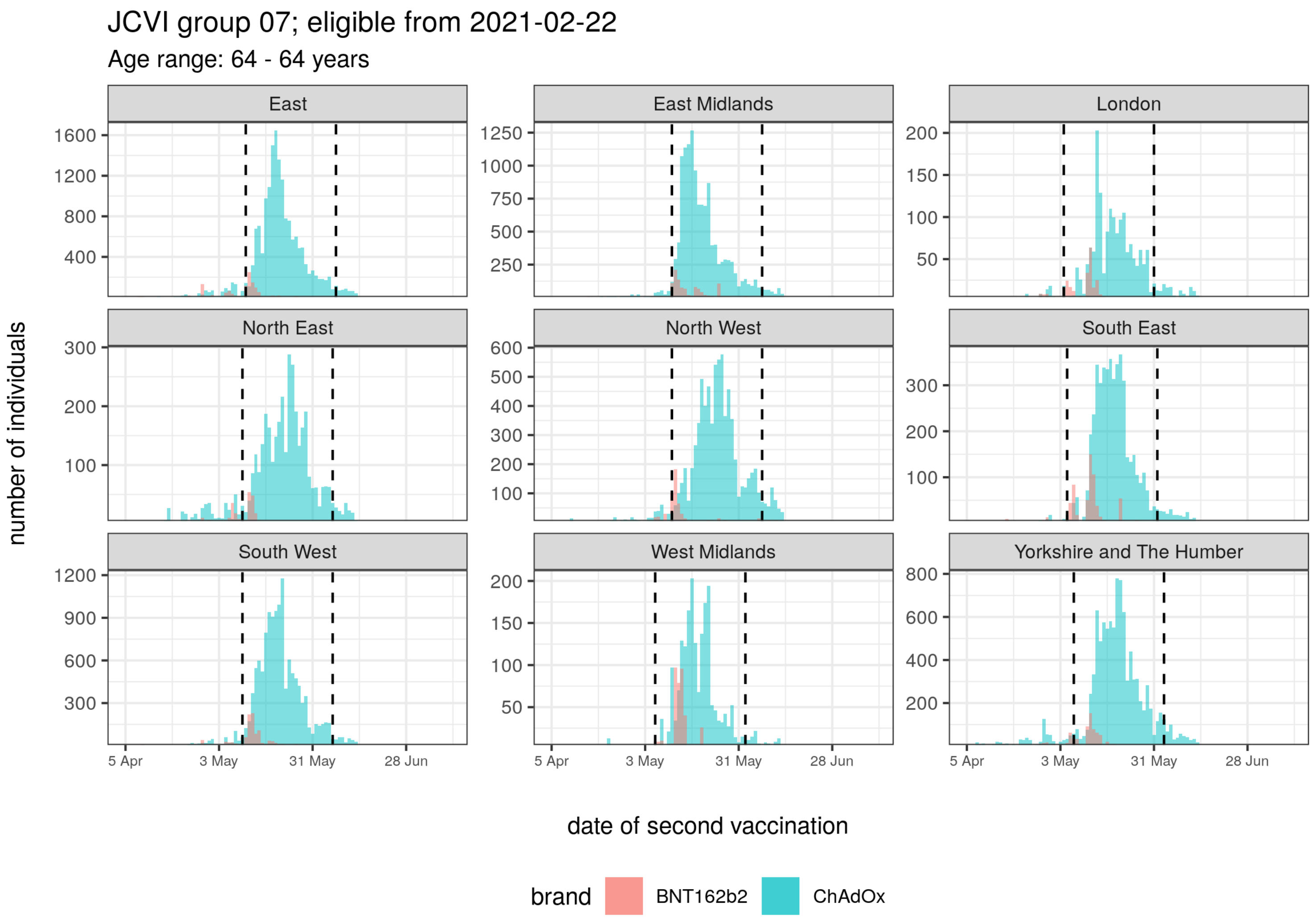

**Supplementary Figure 8:** Second vaccination period for JCVI group 7 and aged 64 years

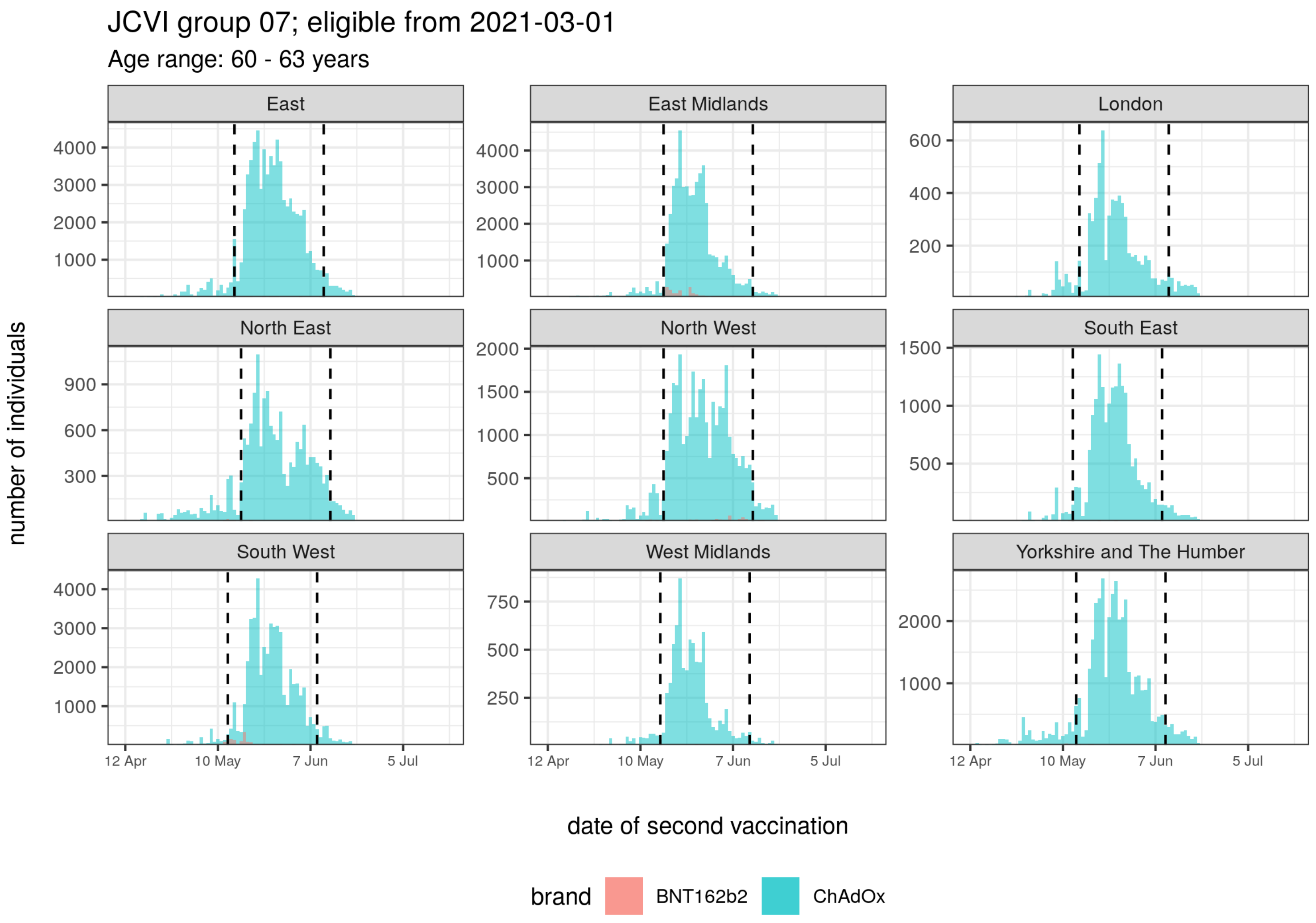

**Supplementary Figure 9:** Second vaccination period for JCVI group 7 and aged 60-63 years

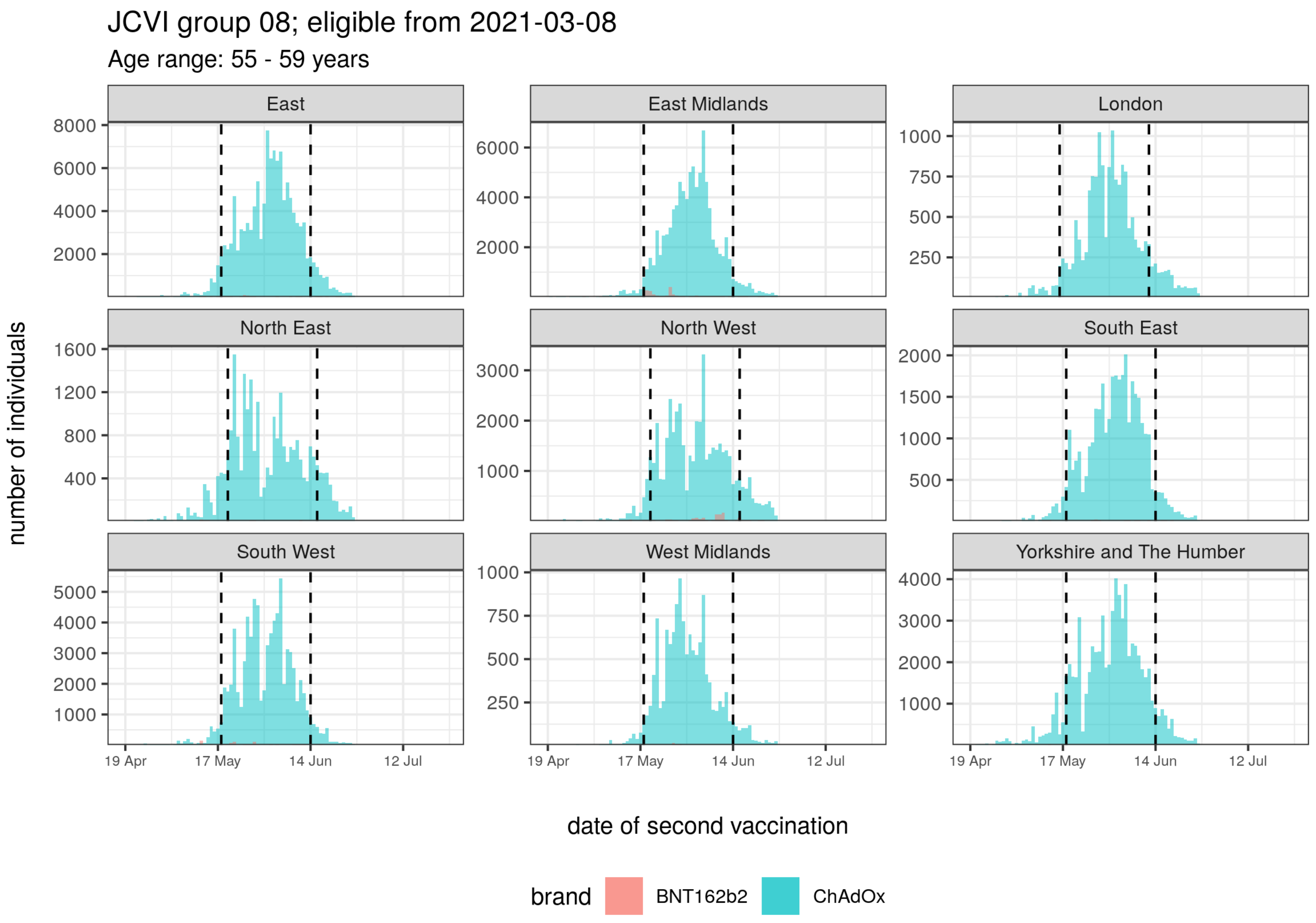

**Supplementary Figure 10:** Second vaccination period for JCVI group 8

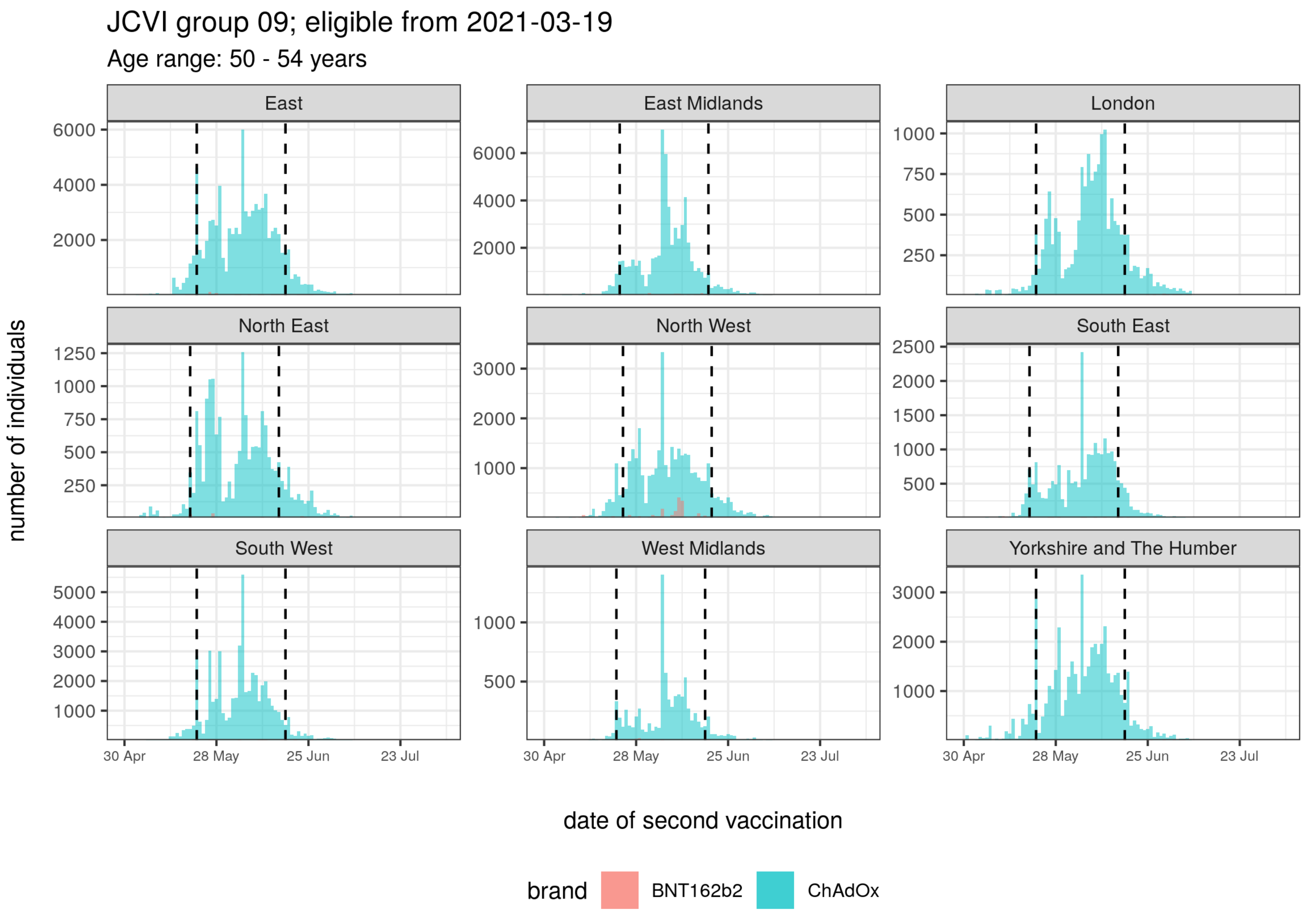

Supplementary Figure 11: Second vaccination period for JCVI group 9

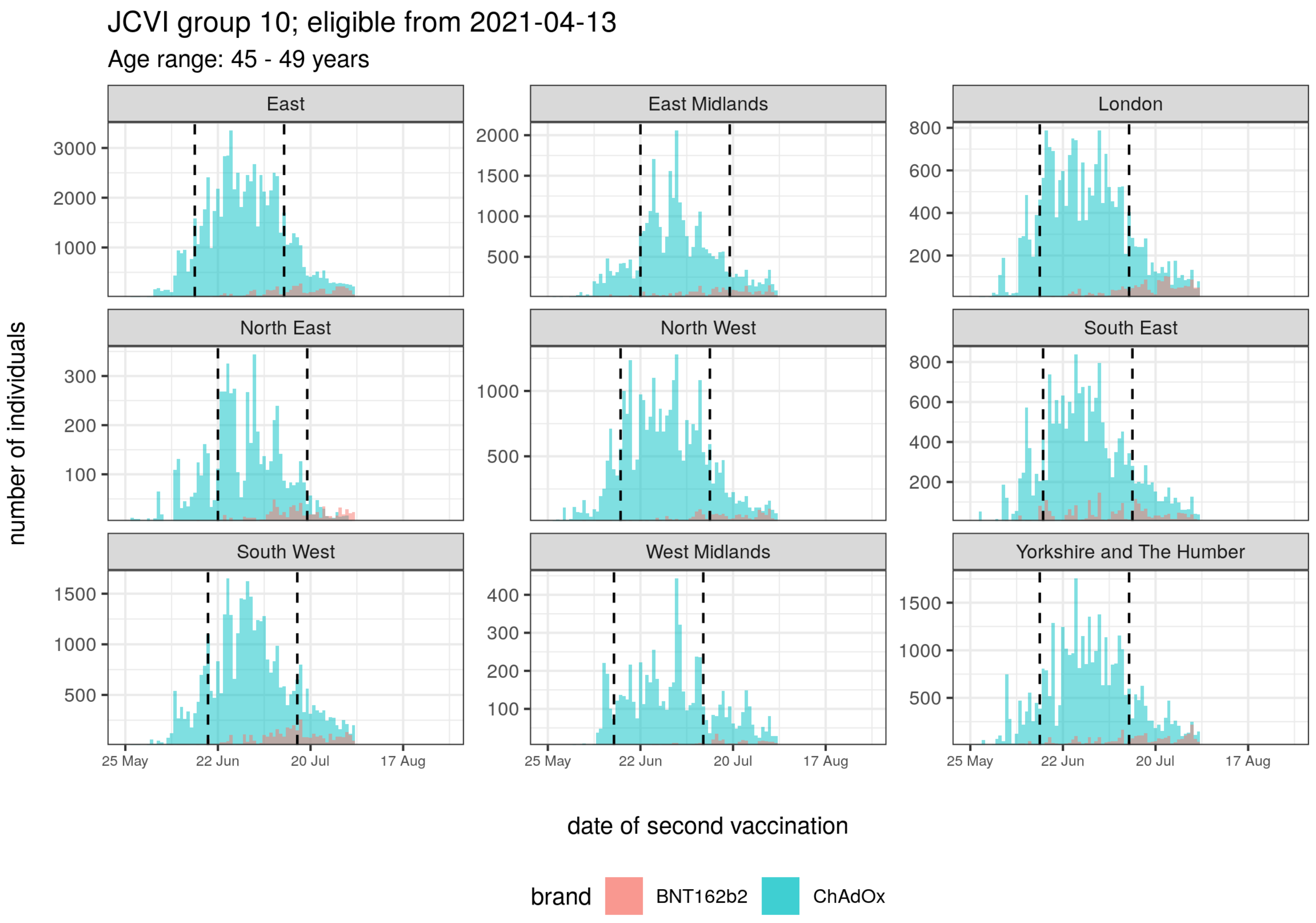

**Supplementary Figure 12:** Second vaccination period for JCVI group 10 and aged 45-49 years

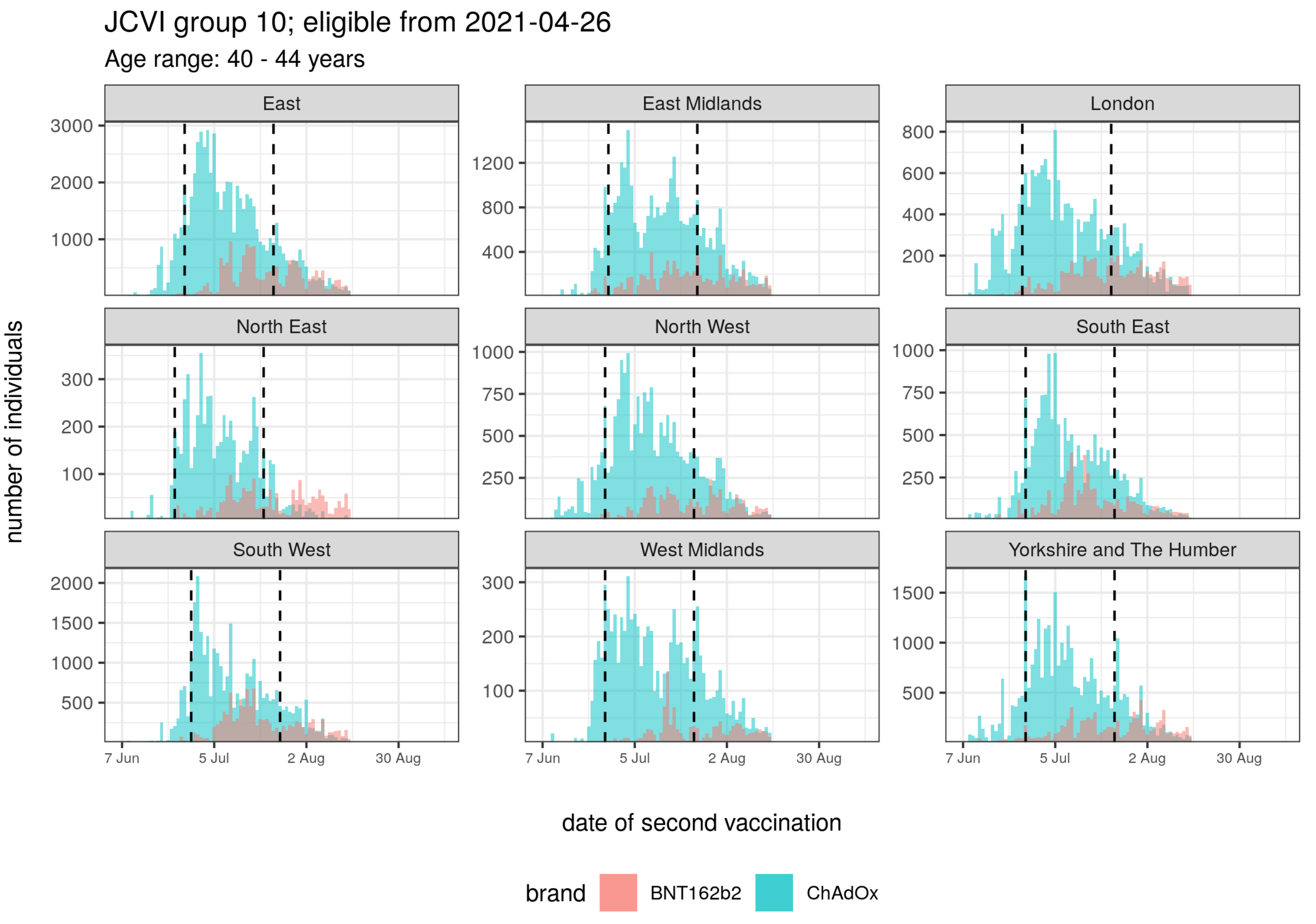

**Supplementary Figure 13:** Second vaccination period for JCVI group 10 and aged 40-44 years

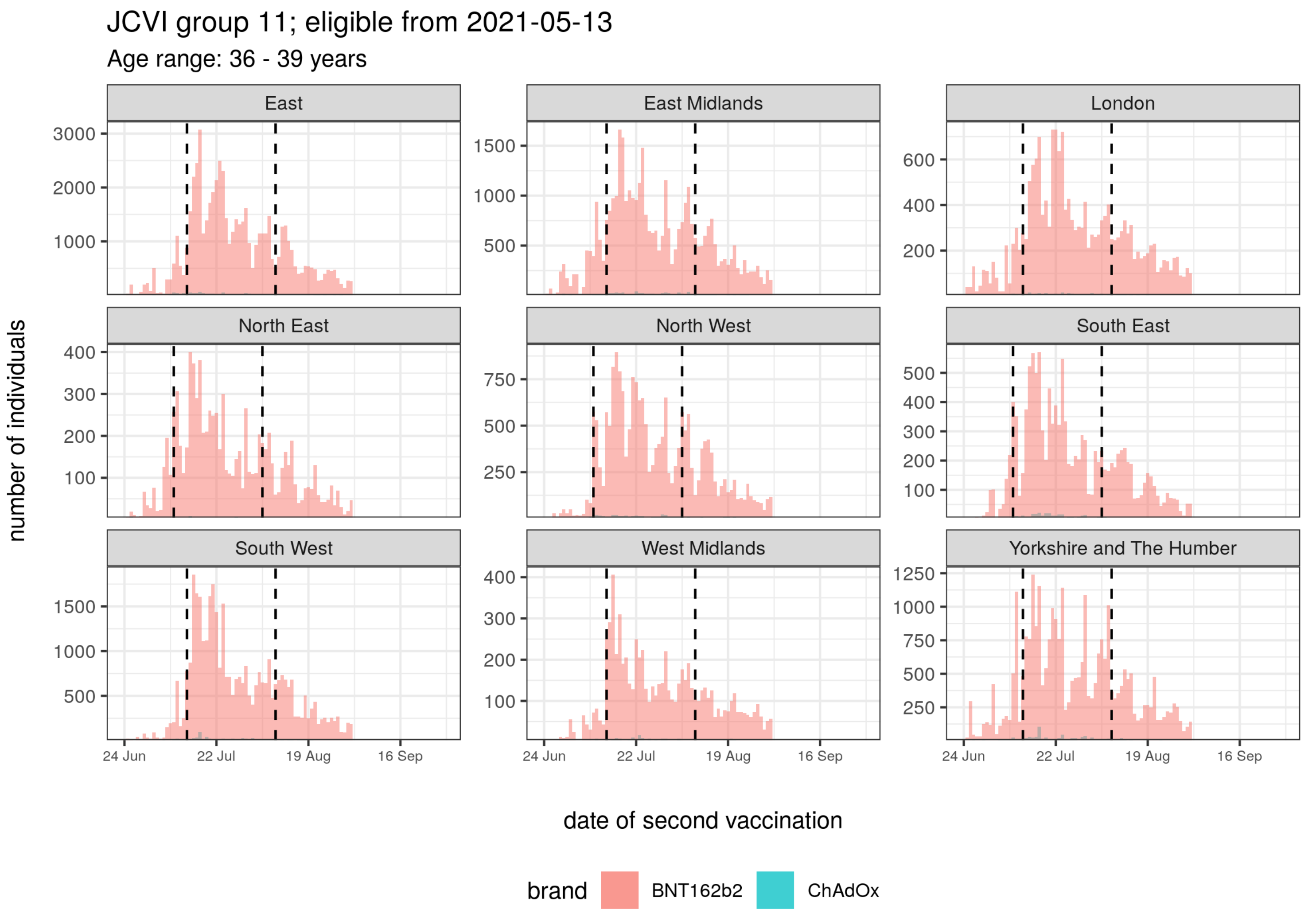

**Supplementary Figure 14:** Second vaccination period for JCVI group 11 and aged 36-39 years

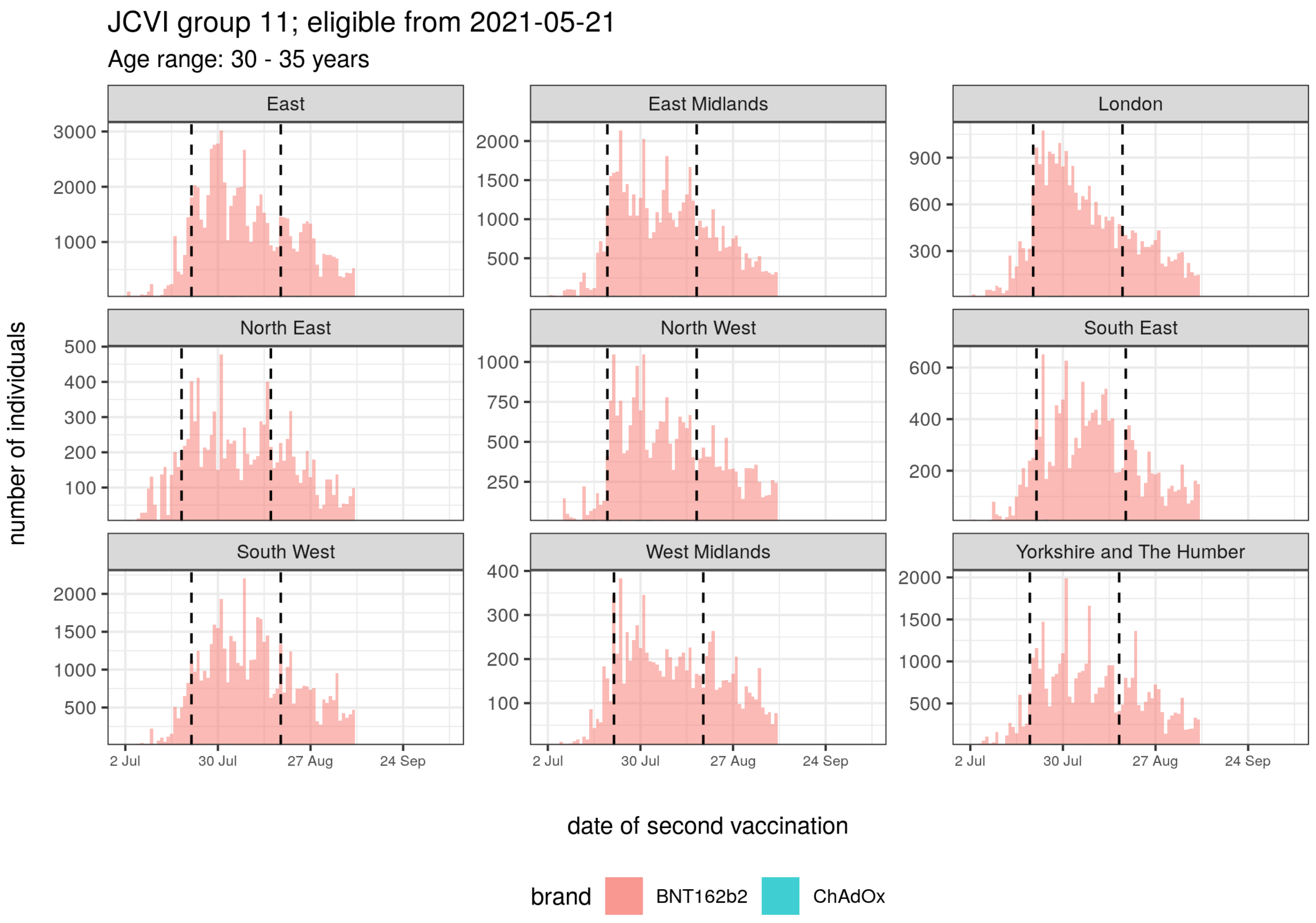

**Supplementary Figure 15:** Second vaccination period for JCVI group 11 and aged 30-35 years

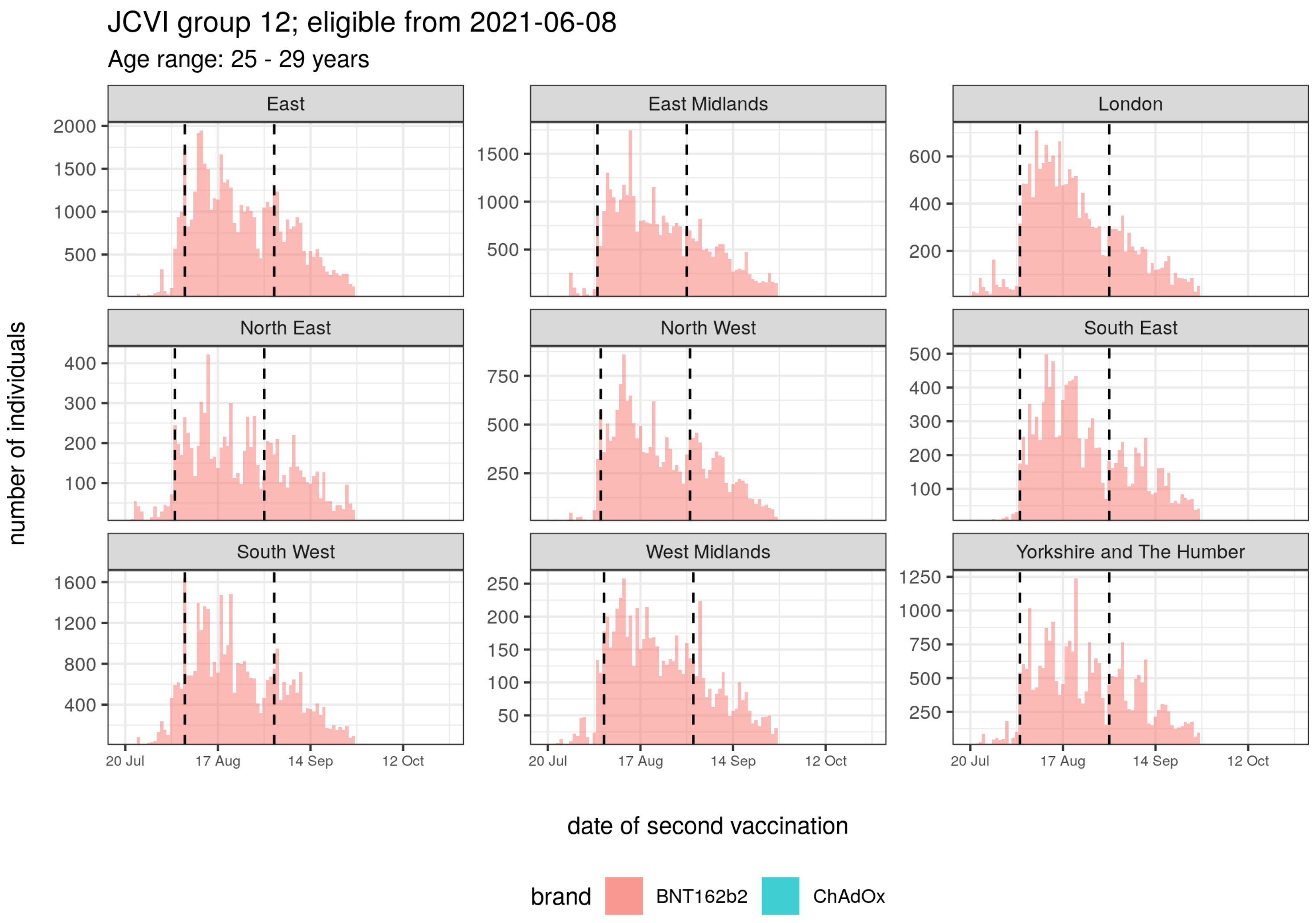

**Supplementary Figure 16:** Second vaccination period for JCVI group 12 and aged 25-29 years

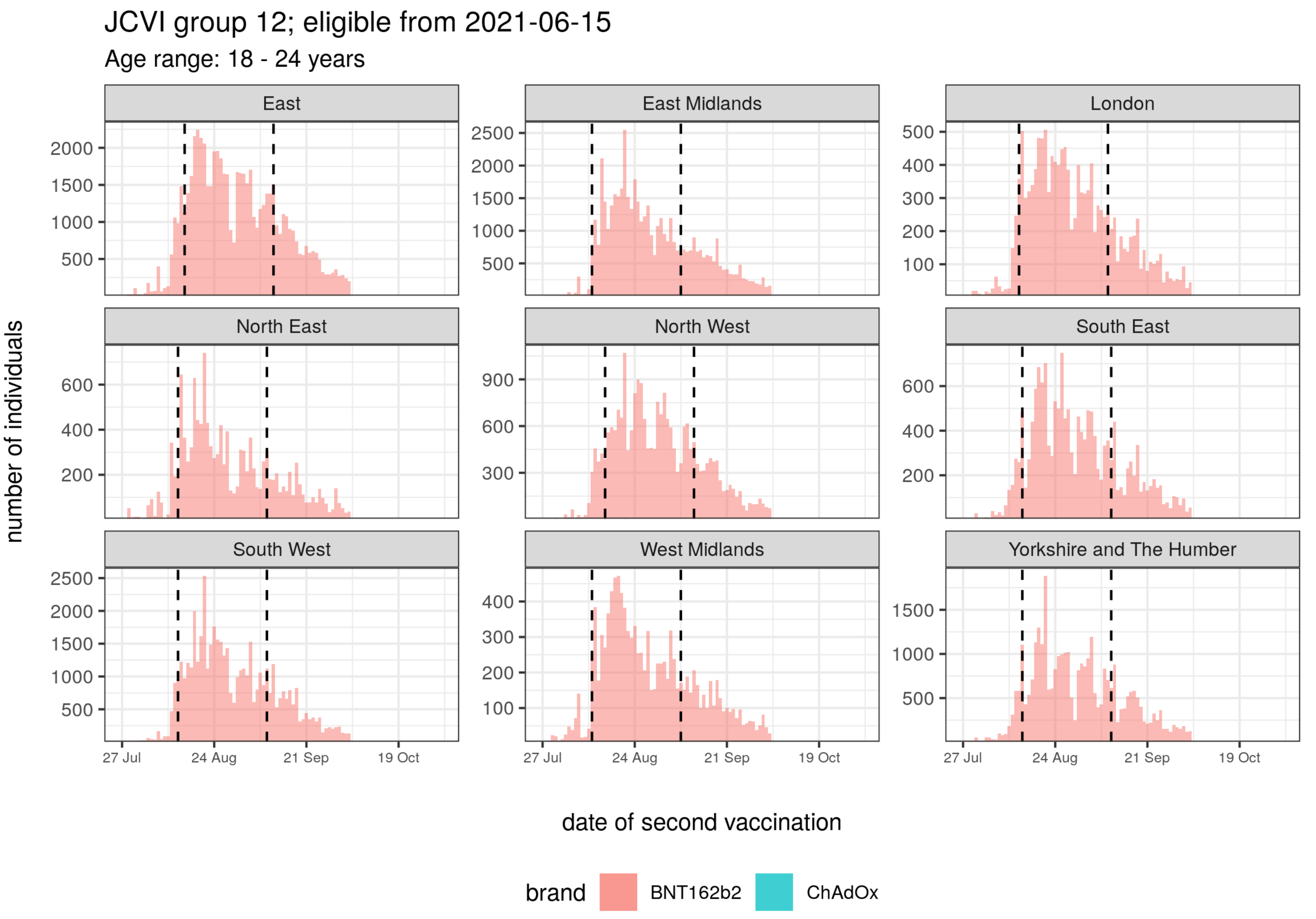

**Supplementary Figure 17:** Second vaccination period for JCVI group 12 and aged 18-24 years

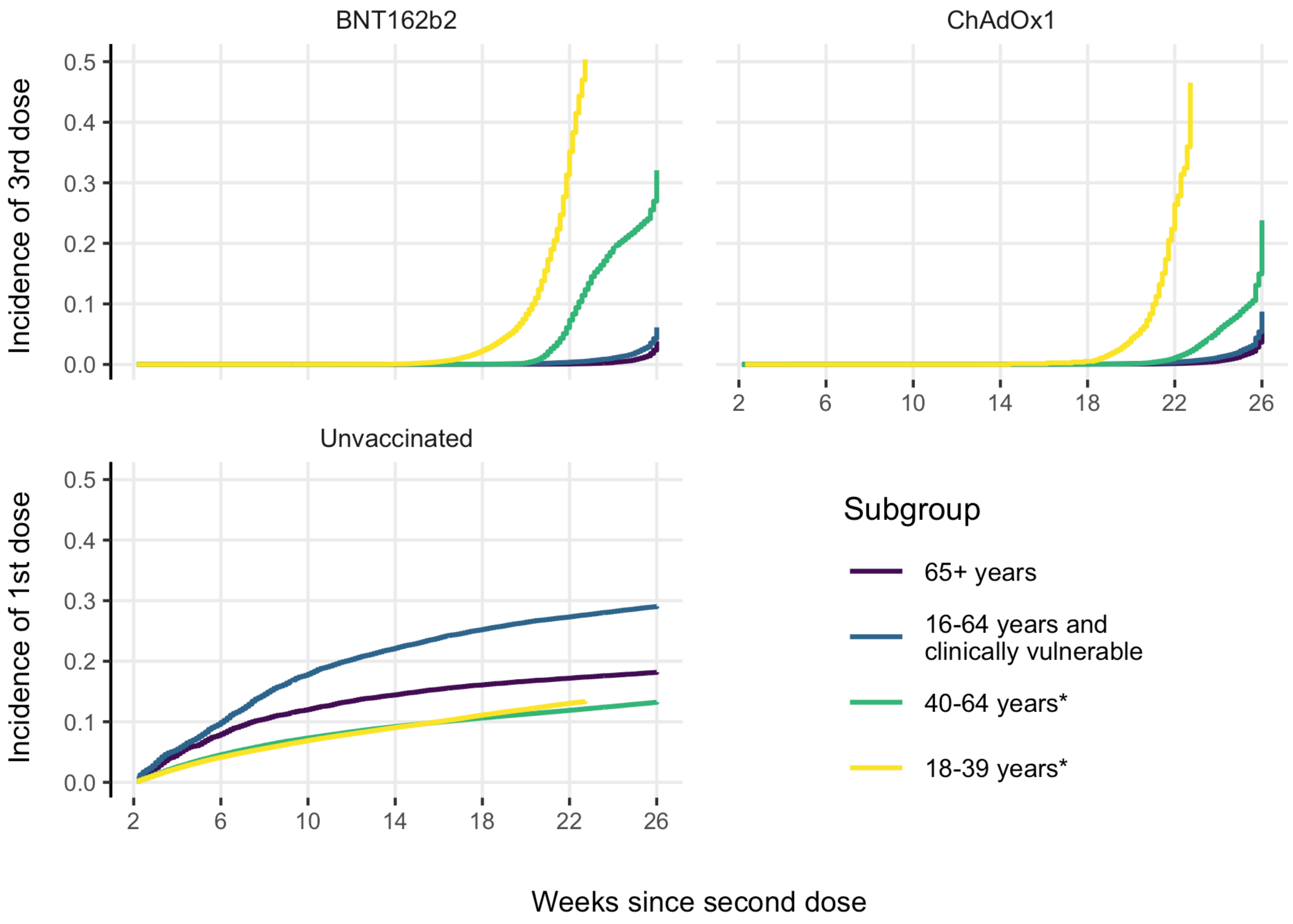

**Supplementary Figure 18:** Cumulative incidence of subsequent vaccination. *And not clinically vulnerable.

**Supplementary Table 4:** Additional characteristics summarised across groups.

| | | 65+ years | | | 16-64 years and clinically vulnerable | | | 40-64 years[a] | | | 18-39 years[a] | |
|---|---|---|---|---|---|---|---|---|---|---|---|---|
| **Variable** | **Characteristic** | **BNT162b2** | **ChAdOx1** | **Unvaccinated** | **BNT162b2** | **ChAdOx1** | **Unvaccinated** | **BNT162b2** | **ChAdOx1** | **Unvaccinated** | **BNT162b2** | **Unvaccinated** |
| | N | 653,435 | 841,239 | 152,294 | 374,137 | 651,574 | 310,445 | 62,652 | 1,463,607 | 623,998 | 683,746 | 1,347,251 |
| Region | East | 126,284 (19%) | 197,728 (24%) | 33,748 (22%) | 81,599 (22%) | 151,537 (23%) | 63,686 (21%) | 13,490 (22%) | 375,750 (26%) | 139,558 (22%) | 164,199 (24%) | 299,336 (22%) |
| | East Midlands | 129,644 (20%) | 141,026 (17%) | 23,292 (15%) | 64,627 (17%) | 133,576 (21%) | 53,088 (17%) | 11,747 (19%) | 247,775 (17%) | 95,693 (15%) | 118,456 (17%) | 220,567 (16%) |
| | London | 23,473 (4%) | 16,323 (2%) | 21,195 (14%) | 14,685 (4%) | 28,210 (4%) | 35,338 (11%) | 3,612 (6%) | 63,552 (4%) | 105,658 (17%) | 52,990 (8%) | 194,521 (14%) |
| | North East | 36,361 (6%) | 42,745 (5%) | 6,271 (4%) | 19,075 (5%) | 32,567 (5%) | 16,697 (5%) | 1,742 (3%) | 63,202 (4%) | 25,191 (4%) | 26,886 (4%) | 61,902 (5%) |
| | North West | 70,496 (11%) | 90,452 (11%) | 10,092 (7%) | 45,305 (12%) | 59,300 (9%) | 24,342 (8%) | 6,420 (10%) | 147,555 (10%) | 39,823 (6%) | 59,547 (9%) | 88,943 (7%) |
| | South East | 43,996 (7%) | 62,863 (7%) | 10,265 (7%) | 20,694 (6%) | 43,310 (7%) | 16,254 (5%) | 5,793 (9%) | 101,695 (7%) | 37,424 (6%) | 38,262 (6%) | 78,179 (6%) |
| | South West | 115,730 (18%) | 142,787 (17%) | 18,816 (12%) | 63,056 (17%) | 89,574 (14%) | 29,691 (10%) | 12,724 (20%) | 231,862 (16%) | 61,391 (10%) | 118,253 (17%) | 124,764 (9%) |
| | West Midlands | 17,664 (3%) | 25,822 (3%) | 8,348 (5%) | 14,083 (4%) | 26,342 (4%) | 19,612 (6%) | 1,334 (2%) | 39,455 (3%) | 31,342 (5%) | 23,183 (3%) | 76,399 (6%) |
| | Yorkshire and The Humber | 89,787 (14%) | 121,493 (14%) | 20,267 (13%) | 51,013 (14%) | 87,158 (13%) | 51,737 (17%) | 5,790 (9%) | 192,761 (13%) | 87,918 (14%) | 81,970 (12%) | 202,640 (15%) |
| JCVI group | 02 | 228,980 (35%) | 26,901 (3%) | 32,978 (22%) | - | - | - | - | - | - | - | - |
| | 03 | 42,926 (7%) | 27,812 (3%) | 26,790 (18%) | - | - | - | - | - | - | - | - |
| | 04 | 289,508 (44%) | 559,548 (67%) | 42,337 (28%) | 63,465 (17%) | 132,777 (20%) | 31,316 (10%) | - | - | - | - | - |
| | 05 | 92,021 (14%) | 226,978 (27%) | 50,189 (33%) | - | - | - | - | - | - | - | - |
| | 06 | - | - | - | 310,672 (83%) | 518,797 (80%) | 279,129 (90%) | - | - | - | - | - |
| | 07 | - | - | - | - | - | - | 8,656 (14%) | 344,277 (24%) | 52,791 (8%) | - | - |
| | 08 | - | - | - | - | - | - | 4,158 (7%) | 452,092 (31%) | 84,346 (14%) | - | - |
| | 09 | - | - | - | - | - | - | 3,319 (5%) | 301,770 (21%) | 113,437 (18%) | - | - |
| | 10 | - | - | - | - | - | - | 46,519 (74%) | 365,468 (25%) | 373,424 (60%) | - | - |
| | 11 | - | - | - | - | - | - | - | - | - | 355,857 (52%) | 672,588 (50%) |
| | 12 | - | - | - | - | - | - | - | - | - | 327,889 (48%) | 674,663 (50%) |
| Resident in long-term residential home | yes | 213 (0%) | 351 (0%) | 84 (0%) | 669 (0%) | 2,494 (0%) | 1,005 (0%) | - (-%) | 85 (0%) | 146 (0%) | 17 (0%) | 238 (0%) |
| | Chronic heart disease | 202,268 (31%) | 197,000 (23%) | 32,936 (22%) | 88,479 (24%) | 146,997 (23%) | 54,064 (17%) | 107 (0%) | 2,819 (0%) | 766 (0%) | 263 (0%) | 673 (0%) |
| | Chronic kidney disease | 114,885 (18%) | 81,160 (10%) | 16,899 (11%) | 21,853 (6%) | 33,314 (5%) | 9,522 (3%) | 18 (0%) | 483 (0%) | 97 (0%) | 25 (0%) | 57 (0%) |
| | Chronic liver disease | 19,282 (3%) | 28,202 (3%) | 4,264 (3%) | 28,863 (8%) | 62,366 (10%) | 34,884 (11%) | 77 (0%) | 1,384 (0%) | 481 (0%) | 365 (0%) | 522 (0%) |
| | Chronic neurological disease | 72,106 (11%) | 70,779 (8%) | 13,421 (9%) | 59,813 (16%) | 103,887 (16%) | 50,922 (16%) | 30 (0%) | 765 (0%) | 239 (0%) | 191 (0%) | 404 (0%) |
| History of | Chronic respiratory disease | 69,405 (11%) | 79,746 (9%) | 11,678 (8%) | 31,628 (8%) | 56,703 (9%) | 18,697 (6%) | 21 (0%) | 712 (0%) | 227 (0%) | 78 (0%) | 140 (0%) |
| | Diabetes | 119,012 (18%) | 138,583 (16%) | 26,295 (17%) | 126,618 (34%) | 186,286 (29%) | 70,472 (23%) | 143 (0%) | 3,763 (0%) | 1,076 (0%) | 762 (0%) | 2,593 (0%) |
| | Immunosupp-ression | 35,830 (5%) | 43,211 (5%) | 4,728 (3%) | 43,745 (12%) | 75,818 (12%) | 26,228 (8%) | 27 (0%) | 797 (0%) | 215 (0%) | 243 (0%) | 421 (0%) |
| | Learning disability | 731 (0%) | 1,382 (0%) | 360 (0%) | 12,770 (3%) | 23,376 (4%) | 11,499 (4%) | - (-%) | 29 (0%) | 7 (0%) | 61 (0%) | 113 (0%) |
| | Serious mental illness | 5,543 (1%) | 8,917 (1%) | 3,588 (2%) | 17,147 (5%) | 32,165 (5%) | 32,580 (10%) | 81 (0%) | 2,362 (0%) | 2,517 (0%) | 279 (0%) | 2,135 (0%) |
| Pregnancy | yes | - (-%) | - (-%) | - (-%) | 751 (0%) | 1,844 (0%) | 10,509 (3%) | 305 (0%) | 298 (0%) | 1,776 (0%) | 8,463 (1%) | 43,354 (3%) |
| Housebound | yes | 8,430 (1%) | 14,878 (2%) | 5,797 (4%) | 1,617 (0%) | 6,270 (1%) | 1,744 (1%) | 6 (0%) | 319 (0%) | 334 (0%) | 37 (0%) | 178 (0%) |

[a] And not clinically vulnerable

**Supplementary Table 5:** Event counts during each comparison period.

| Outcome | Weeks since 2nd dose | 65+ years | | | 16-64 years and clinically vulnerable | | | 40-64 years[a] | | | 18-39 years[a] | |
|---|---|---|---|---|---|---|---|---|---|---|---|---|
| | | BNT162b2 | ChAdOx1 | Unvaccinated | BNT162b2 | ChAdOx1 | Unvaccinated | BNT162b2 | ChAdOx1 | Unvaccinated | BNT162b2 | Unvaccinated |
| COVID-19 hospitalisation | 3-6 | 27 / 50,048 | 34 / 64,444 | 76 / 11,499 | - / 28,664 | 27 / 49,918 | 166 / 23,652 | - / 4,800 | 59 / 112,139 | 534 / 47,422 | 13 / 52,239 | 727 / 102,580 |
| | 7-10 | 24 / 50,151 | 77 / 64,331 | 96 / 10,608 | 27 / 28,621 | 157 / 49,830 | 437 / 21,235 | - / 4,794 | 110 / 111,997 | 569 / 45,219 | 26 / 51,904 | 554 / 97,782 |
| | 11-14 | 93 / 50,034 | 236 / 64,198 | 182 / 9,955 | 61 / 28,577 | 304 / 49,730 | 633 / 19,171 | - / 4,787 | 114 / 111,860 | 539 / 43,406 | 32 / 51,624 | 514 / 93,406 |
| | 15-18 | 230 / 49,897 | 431 / 64,048 | 275 / 9,655 | 108 / 28,525 | 395 / 49,614 | 505 / 18,087 | - / 4,779 | 171 / 111,714 | 536 / 42,578 | 29 / 41,176 | 384 / 66,786 |
| | 19-22 | 325 / 49,748 | 558 / 63,883 | 339 / 9,282 | 97 / 28,461 | 400 / 49,480 | 433 / 17,207 | - / 4,567 | 196 / 110,950 | 439 / 38,587 | 19 / 12,745 | 129 / 21,855 |
| | 23-26 | 488 / 49,554 | 612 / 63,639 | 360 / 9,149 | 132 / 28,307 | 495 / 49,180 | 416 / 16,670 | - / 1,783 | 235 / 94,138 | 311 / 25,541 | - / 50 | - / 552 |
| | Total | 1,187 / 299,431 | 1,948 / 384,543 | 1,328 / 60,148 | 425 / 171,155 | 1,778 / 297,753 | 2,590 / 116,023 | - / 25,509 | 885 / 652,799 | 2,928 / 242,754 | 119 / 209,736 | 2,308 / 382,960 |
| COVID-19 death | 3-6 | 6 / 50,051 | - / 64,446 | 21 / 11,506 | - / 28,665 | - / 49,920 | 9 / 23,661 | - / 4,800 | - / 112,144 | 14 / 47,459 | - / 52,241 | 6 / 102,680 |
| | 7-10 | - / 50,155 | - / 64,336 | 21 / 10,619 | - / 28,623 | - / 49,838 | 29 / 21,268 | - / 4,794 | - / 112,009 | 28 / 45,294 | - / 51,907 | 11 / 97,921 |
| | 11-14 | 9 / 50,041 | 28 / 64,214 | 41 / 9,975 | - / 28,582 | 21 / 49,754 | 66 / 19,239 | - / 4,787 | - / 111,880 | 28 / 43,524 | - / 51,629 | - / 93,579 |
| | 15-18 | 26 / 49,915 | 70 / 64,085 | 72 / 9,686 | 8 / 28,536 | 31 / 49,664 | 56 / 18,190 | - / 4,780 | 6 / 111,744 | 38 / 42,721 | - / 41,182 | 8 / 66,936 |
| | 19-22 | 84 / 49,782 | 105 / 63,953 | 96 / 9,333 | 15 / 28,479 | 34 / 49,558 | 60 / 17,334 | - / 4,567 | 9 / 110,993 | 30 / 38,746 | - / 12,747 | - / 21,912 |
| | 23-26 | 103 / 49,611 | 90 / 63,743 | 115 / 9,212 | 14 / 28,333 | 41 / 49,288 | 54 / 16,820 | - / 1,783 | 14 / 94,188 | 28 / 25,669 | - / 50 | - / 554 |
| | Total | 228 / 299,555 | 293 / 384,777 | 366 / 60,331 | 37 / 171,219 | 127 / 298,021 | 274 / 116,511 | - / 25,511 | 29 / 652,958 | 166 / 243,413 | - / 209,756 | 25 / 383,583 |
| Positive SARS-CoV-2 test | 3-6 | 90 / 50,040 | 203 / 64,430 | 130 / 11,492 | 150 / 28,653 | 1,391 / 49,863 | 1,668 / 23,578 | 224 / 4,774 | 8,831 / 111,589 | 5,821 / 46,951 | 2,659 / 51,802 | 16,168 / 100,318 |
| | 7-10 | 275 / 50,134 | 1,111 / 64,281 | 230 / 10,596 | 1,420 / 28,567 | 5,455 / 49,527 | 3,502 / 21,002 | 384 / 4,744 | 13,301 / 110,568 | 5,892 / 44,382 | 4,016 / 51,222 | 13,908 / 94,552 |
| | 11-14 | 865 / 49,979 | 2,784 / 64,011 | 382 / 9,935 | 2,395 / 28,371 | 7,680 / 48,938 | 4,107 / 18,708 | 689 / 4,700 | 17,268 / 109,314 | 5,567 / 42,210 | 7,139 / 50,540 | 15,838 / 89,372 |
| | 15-18 | 1,860 / 49,755 | 4,560 / 63,609 | 519 / 9,616 | 3,136 / 28,112 | 8,295 / 48,220 | 3,219 / 17,424 | 991 / 4,625 | 20,204 / 107,723 | 5,843 / 41,029 | 11,547 / 39,747 | 13,600 / 63,229 |
| | 19-22 | 2,368 / 49,462 | 5,081 / 63,107 | 632 / 9,228 | 3,385 / 27,815 | 9,801 / 47,461 | 2,702 / 16,398 | 1,350 / 4,342 | 28,033 / 105,257 | 5,795 / 36,894 | 6,159 / 12,006 | 4,975 / 20,530 |
| | 23-26 | 3,474 / 49,084 | 7,337 / 62,463 | 649 / 9,075 | 5,162 / 27,344 | 12,691 / 46,330 | 3,059 / 15,713 | 400 / 1,705 | 26,822 / 88,321 | 4,196 / 24,387 | 33 / 45 | 157 / 514 |
| | Total | 8,932 / 298,454 | 21,076 / 381,899 | 2,542 / 59,943 | 15,648 / 168,863 | 45,313 / 290,339 | 18,257 / 112,823 | 4,038 / 24,891 | 114,459 / 632,772 | 33,114 / 235,852 | 31,553 / 205,362 | 64,646 / 368,515 |
| Non-COVID-19 death | 3-6 | 745 / 50,051 | 681 / 64,446 | 441 / 11,506 | 127 / 28,665 | 198 / 49,920 | 145 / 23,661 | - / 4,800 | 82 / 112,144 | 80 / 47,459 | - / 52,241 | 19 / 102,680 |
| | 7-10 | 982 / 50,155 | 782 / 64,336 | 390 / 10,619 | 128 / 28,623 | 247 / 49,838 | 149 / 21,268 | - / 4,794 | 104 / 112,009 | 67 / 45,294 | - / 51,907 | 8 / 97,921 |
| | 11-14 | 1,065 / 50,041 | 855 / 64,214 | 331 / 9,975 | 144 / 28,582 | 288 / 49,754 | 130 / 19,239 | - / 4,787 | 114 / 111,880 | 63 / 43,524 | - / 51,629 | 15 / 93,579 |
| | 15-18 | 1,143 / 49,915 | 947 / 64,085 | 328 / 9,686 | 153 / 28,536 | 297 / 49,664 | 127 / 18,190 | - / 4,780 | 143 / 111,744 | 50 / 42,721 | - / 41,182 | 8 / 66,936 |
| | 19-22 | 1,175 / 49,782 | 962 / 63,953 | 287 / 9,333 | 137 / 28,479 | 289 / 49,558 | 87 / 17,334 | 9 / 4,567 | 146 / 110,993 | 66 / 38,746 | - / 12,747 | 6 / 21,912 |
| | 23-26 | 1,336 / 49,611 | 1,053 / 63,743 | 302 / 9,212 | 174 / 28,333 | 313 / 49,288 | 96 / 16,820 | - / 1,783 | 145 / 94,188 | 42 / 25,669 | - / 50 | - / 554 |
| | Total | 6,446 / 299,555 | 5,280 / 384,777 | 2,079 / 60,331 | 863 / 171,219 | 1,632 / 298,021 | 734 / 116,511 | 9 / 25,511 | 734 / 652,958 | 368 / 243,413 | - / 209,756 | 56 / 383,583 |
| Any SARS-CoV-2 test | 3-6 | 52,539 / 47,779 | 81,127 / 60,725 | 6,549 / 10,936 | 48,931 / 26,275 | 91,551 / 45,529 | 20,732 / 21,914 | 9,077 / 4,363 | 239,878 / 100,297 | 34,921 / 44,356 | 115,954 / 47,006 | 99,519 / 93,091 |
| | 7-10 | 59,197 / 47,528 | 90,700 / 60,225 | 6,159 / 10,077 | 60,373 / 25,822 | 108,393 / 44,579 | 21,682 / 19,382 | 10,528 / 4,290 | 247,732 / 99,182 | 34,385 / 41,854 | 116,457 / 46,231 | 84,765 / 88,159 |
| | 11-14 | 64,240 / 47,115 | 96,581 / 59,616 | 5,876 / 9,458 | 57,568 / 25,691 | 102,043 / 44,464 | 20,554 / 17,267 | 11,036 / 4,220 | 248,568 / 98,644 | 32,726 / 39,764 | 113,612 / 45,858 | 77,169 / 84,177 |
| | 15-18 | 64,891 / 46,911 | 96,184 / 59,417 | 5,987 / 9,142 | 59,454 / 25,598 | 107,656 / 43,692 | 19,349 / 16,014 | 11,396 / 4,139 | 272,107 / 95,703 | 31,167 / 38,708 | 112,953 / 36,010 | 60,931 / 60,425 |
| | 19-22 | 69,277 / 46,506 | 106,642 / 58,540 | 5,927 / 8,758 | 64,945 / 24,950 | 114,891 / 42,572 | 17,094 / 15,096 | 12,180 / 3,873 | 279,888 / 93,366 | 28,563 / 35,009 | 53,658 / 10,806 | 23,726 / 19,987 |
| | 23-26 | 76,071 / 45,885 | 116,065 / 57,584 | 5,934 / 8,609 | 65,995 / 24,474 | 113,673 / 41,560 | 15,949 / 14,542 | 5,479 / 1,541 | 248,893 / 78,716 | 21,113 / 23,346 | 412 / 44 | 897 / 513 |
| | Total | 386,215 / 281,723 | 587,299 / 356,106 | 36,432 / 56,981 | 357,266 / 152,810 | 638,207 / 262,397 | 115,360 / 104,215 | 59,696 / 22,428 | 1,537,066 / 565,909 | 182,875 / 223,036 | 513,046 / 185,955 | 347,007 / 346,351 |

[a] And not clinically vulnerable

**Supplementary Table 6:** Unadjusted hazard ratios for each comparison period.

| | | 65+ years | | | 16-64 years and clinically vulnerable | | | 40-64 years[a] | | | 18-39 years[a] |
|---|---|---|---|---|---|---|---|---|---|---|---|
| **Outcome** | **Weeks since 2nd dose** | **BNT162b2 vs unvaccinated** | **ChAdOx1 vs unvaccinated** | **BNT162b2 vs ChAdOx1** | **BNT162b2 vs unvaccinated** | **ChAdOx1 vs unvaccinated** | **BNT162b2 vs ChAdOx1** | **BNT162b2 vs unvaccinated** | **ChAdOx1 vs unvaccinated** | **BNT162b2 vs ChAdOx1** | **BNT162b2 vs unvaccinated** |
| COVID-19 hospitalisation | 3-6 | 0.08 (0.05-0.14) | 0.13 (0.08-0.20) | 0.42 (0.23-0.75) | - | 0.10 (0.07-0.15) | - | - | 0.06 (0.04-0.08) | - | 0.03 (0.02-0.06) |
| | 7-10 | 0.06 (0.04-0.10) | 0.12 (0.09-0.16) | 0.45 (0.29-0.71) | 0.06 (0.04-0.08) | 0.14 (0.12-0.17) | 0.37 (0.25-0.56) | - | 0.06 (0.05-0.07) | - | 0.09 (0.06-0.13) |
| | 11-14 | 0.11 (0.08-0.14) | 0.15 (0.12-0.18) | 0.58 (0.45-0.74) | 0.06 (0.05-0.08) | 0.18 (0.16-0.21) | 0.37 (0.28-0.48) | - | 0.06 (0.05-0.07) | - | 0.10 (0.07-0.14) |
| | 15-18 | 0.13 (0.11-0.16) | 0.18 (0.16-0.21) | 0.58 (0.49-0.68) | 0.12 (0.10-0.14) | 0.26 (0.23-0.29) | 0.50 (0.40-0.61) | - | 0.11 (0.09-0.13) | - | 0.11 (0.08-0.16) |
| | 19-22 | 0.15 (0.13-0.17) | 0.23 (0.21-0.27) | 0.55 (0.48-0.63) | 0.13 (0.11-0.17) | 0.30 (0.26-0.33) | 0.49 (0.40-0.61) | - | 0.13 (0.11-0.15) | - | 0.22 (0.14-0.36) |
| | 23-26 | 0.21 (0.18-0.24) | 0.28 (0.24-0.32) | 0.73 (0.64-0.82) | 0.17 (0.14-0.20) | 0.35 (0.31-0.39) | 0.48 (0.40-0.58) | - | 0.17 (0.14-0.20) | - | - |
| COVID-19 death | 3-6 | 0.04 (0.02-0.12) | - | - | - | - | - | - | - | - | - |
| | 7-10 | - | - | - | - | - | - | - | - | - | - |
| | 11-14 | 0.06 (0.03-0.12) | 0.10 (0.06-0.15) | 0.50 (0.22-1.12) | - | 0.13 (0.08-0.20) | - | - | - | - | - |
| | 15-18 | 0.06 (0.04-0.10) | 0.14 (0.10-0.19) | 0.34 (0.20-0.57) | 0.07 (0.04-0.16) | 0.16 (0.10-0.23) | 0.45 (0.19-1.04) | - | 0.04 (0.02-0.09) | - | - |
| | 19-22 | 0.11 (0.08-0.15) | 0.14 (0.11-0.19) | 0.65 (0.48-0.87) | 0.15 (0.08-0.27) | 0.17 (0.11-0.25) | 0.92 (0.49-1.71) | - | 0.06 (0.03-0.13) | - | - |
| | 23-26 | 0.12 (0.09-0.16) | 0.17 (0.13-0.23) | 0.68 (0.50-0.91) | 0.13 (0.07-0.24) | 0.21 (0.14-0.31) | 0.62 (0.34-1.12) | - | 0.09 (0.05-0.18) | - | - |
| Positive SARS-CoV-2 test | 3-6 | 0.22 (0.16-0.29) | 0.55 (0.44-0.69) | 0.62 (0.47-0.81) | 0.17 (0.14-0.20) | 0.60 (0.56-0.65) | 0.34 (0.28-0.40) | 0.34 (0.29-0.39) | 0.93 (0.90-0.96) | 0.36 (0.32-0.41) | 0.31 (0.30-0.33) |
| | 7-10 | 0.41 (0.34-0.48) | 0.78 (0.70-0.86) | 0.54 (0.48-0.62) | 0.30 (0.29-0.32) | 0.63 (0.61-0.65) | 0.49 (0.46-0.51) | 0.58 (0.52-0.64) | 1.10 (1.07-1.13) | 0.48 (0.43-0.53) | 0.53 (0.51-0.55) |
| | 11-14 | 0.50 (0.45-0.55) | 0.83 (0.77-0.89) | 0.61 (0.57-0.66) | 0.38 (0.36-0.40) | 0.74 (0.72-0.77) | 0.57 (0.55-0.60) | 0.94 (0.86-1.01) | 1.44 (1.40-1.48) | 0.54 (0.50-0.59) | 0.77 (0.75-0.79) |
| | 15-18 | 0.67 (0.62-0.73) | 1.02 (0.96-1.08) | 0.70 (0.66-0.73) | 0.54 (0.51-0.56) | 0.93 (0.90-0.97) | 0.63 (0.60-0.65) | 1.22 (1.14-1.31) | 1.80 (1.76-1.84) | 0.62 (0.58-0.66) | 1.25 (1.22-1.28) |
| | 19-22 | 0.75 (0.70-0.81) | 1.21 (1.14-1.28) | 0.70 (0.67-0.73) | 0.77 (0.74-0.81) | 1.18 (1.14-1.22) | 0.67 (0.64-0.70) | 1.54 (1.45-1.64) | 2.12 (2.08-2.17) | 0.66 (0.63-0.70) | 1.87 (1.80-1.93) |
| | 23-26 | 0.99 (0.92-1.07) | 1.46 (1.36-1.56) | 0.76 (0.73-0.80) | 0.91 (0.87-0.95) | 1.35 (1.31-1.40) | 0.69 (0.67-0.71) | 1.35 (1.21-1.51) | 2.43 (2.36-2.49) | 0.60 (0.55-0.67) | 2.18 (1.53-3.11) |
| Non-COVID-19 death | 3-6 | 0.30 (0.27-0.34) | 0.57 (0.49-0.65) | 0.61 (0.54-0.70) | 0.62 (0.48-0.79) | 0.52 (0.43-0.64) | 1.21 (0.96-1.51) | - | 0.31 (0.22-0.43) | - | - |
| | 7-10 | 0.39 (0.35-0.43) | 0.63 (0.56-0.71) | 0.71 (0.64-0.78) | 0.58 (0.46-0.73) | 0.58 (0.49-0.70) | 1.00 (0.81-1.23) | - | 0.38 (0.29-0.51) | - | - |
| | 11-14 | 0.45 (0.41-0.50) | 0.67 (0.60-0.76) | 0.72 (0.66-0.79) | 0.63 (0.51-0.79) | 0.68 (0.57-0.80) | 0.91 (0.75-1.11) | - | 0.52 (0.39-0.68) | - | - |
| | 15-18 | 0.49 (0.44-0.54) | 0.74 (0.66-0.83) | 0.69 (0.63-0.75) | 0.73 (0.59-0.90) | 0.84 (0.70-0.99) | 1.02 (0.84-1.23) | - | 0.51 (0.39-0.66) | - | - |
| | 19-22 | 0.51 (0.47-0.57) | 0.79 (0.70-0.89) | 0.70 (0.64-0.76) | 0.67 (0.53-0.84) | 0.77 (0.64-0.93) | 0.84 (0.69-1.02) | 1.18 (0.56-2.51) | 0.53 (0.41-0.69) | 2.70 (1.30-5.60) | - |
| | 23-26 | 0.58 (0.52-0.65) | 0.86 (0.75-0.99) | 0.77 (0.71-0.85) | 0.97 (0.75-1.24) | 0.99 (0.80-1.22) | 1.04 (0.87-1.25) | - | 0.53 (0.40-0.71) | - | - |
| Any SARS-CoV-2 test | 3-6 | 1.17 (1.14-1.19) | 1.16 (1.14-1.18) | 0.72 (0.72-0.73) | 1.48 (1.46-1.50) | 1.38 (1.37-1.40) | 0.70 (0.69-0.71) | 2.36 (2.30-2.42) | 1.77 (1.75-1.78) | 0.77 (0.75-0.79) | 2.00 (1.99-2.02) |
| | 7-10 | 1.72 (1.70-1.76) | 1.90 (1.87-1.93) | 0.88 (0.87-0.89) | 1.89 (1.87-1.92) | 1.89 (1.87-1.91) | 0.95 (0.94-0.96) | 2.70 (2.64-2.77) | 2.84 (2.82-2.86) | 0.84 (0.82-0.86) | 2.55 (2.53-2.57) |
| | 11-14 | 2.14 (2.10-2.18) | 2.64 (2.59-2.69) | 0.92 (0.91-0.93) | 2.00 (1.98-2.03) | 2.08 (2.06-2.11) | 1.02 (1.01-1.03) | 3.02 (2.95-3.08) | 3.43 (3.40-3.46) | 0.85 (0.83-0.86) | 2.82 (2.80-2.84) |
| | 15-18 | 2.62 (2.56-2.67) | 3.66 (3.58-3.75) | 0.93 (0.92-0.94) | 1.93 (1.90-1.95) | 2.22 (2.19-2.25) | 0.97 (0.96-0.98) | 3.30 (3.22-3.37) | 4.49 (4.45-4.54) | 0.86 (0.85-0.88) | 3.46 (3.43-3.49) |
| | 19-22 | 3.46 (3.37-3.55) | 5.60 (5.45-5.76) | 0.94 (0.93-0.94) | 2.38 (2.35-2.42) | 2.90 (2.85-2.94) | 0.99 (0.98-1.00) | 3.63 (3.55-3.71) | 6.84 (6.75-6.93) | 0.86 (0.84-0.88) | 5.47 (5.39-5.55) |
| | 23-26 | 5.53 (5.36-5.70) | 10.09 (9.75-10.45) | 1.07 (1.06-1.08) | 3.43 (3.37-3.50) | 4.50 (4.42-4.59) | 1.08 (1.07-1.09) | 5.03 (4.86-5.21) | 13.46 (13.24-13.69) | 1.10 (1.07-1.14) | 10.85 (9.75-12.07) |

[a] And not clinically vulnerable

**Supplementary Table 7:** Adjusted hazard ratios for each comparison period.

| | | 65+ years | | | 16-64 years and clinically vulnerable | | | 40-64 years[a] | | | 18-39 years[a] |
|---|---|---|---|---|---|---|---|---|---|---|---|
| **Outcome** | **Weeks since 2nd dose** | **BNT162b2 vs unvaccinated** | **ChAdOx1 vs unvaccinated** | **BNT162b2 vs ChAdOx1** | **BNT162b2 vs unvaccinated** | **ChAdOx1 vs unvaccinated** | **BNT162b2 vs ChAdOx1** | **BNT162b2 vs unvaccinated** | **ChAdOx1 vs unvaccinated** | **BNT162b2 vs ChAdOx1** | **BNT162b2 vs unvaccinated** |
| COVID-19 hospitalisation | 3-6 | 0.09 (0.05-0.14) | 0.13 (0.08-0.20) | 0.44 (0.25-0.78) | - | 0.08 (0.05-0.12) | - | - | 0.05 (0.04-0.07) | - | 0.04 (0.02-0.06) |
| | 7-10 | 0.06 (0.04-0.10) | 0.12 (0.09-0.16) | 0.45 (0.29-0.71) | 0.04 (0.03-0.06) | 0.11 (0.09-0.13) | 0.37 (0.24-0.55) | - | 0.05 (0.04-0.06) | - | 0.09 (0.06-0.14) |
| | 11-14 | 0.11 (0.08-0.13) | 0.15 (0.12-0.18) | 0.59 (0.46-0.75) | 0.05 (0.04-0.06) | 0.13 (0.12-0.15) | 0.36 (0.28-0.48) | - | 0.05 (0.04-0.06) | - | 0.10 (0.07-0.14) |
| | 15-18 | 0.13 (0.11-0.16) | 0.18 (0.15-0.20) | 0.59 (0.50-0.70) | 0.09 (0.07-0.11) | 0.19 (0.17-0.21) | 0.49 (0.40-0.61) | - | 0.10 (0.08-0.11) | - | 0.11 (0.07-0.15) |
| | 19-22 | 0.14 (0.12-0.17) | 0.22 (0.19-0.25) | 0.56 (0.49-0.65) | 0.10 (0.08-0.13) | 0.22 (0.19-0.25) | 0.49 (0.39-0.60) | - | 0.11 (0.09-0.13) | - | 0.20 (0.12-0.32) |
| | 23-26 | 0.20 (0.18-0.24) | 0.26 (0.23-0.30) | 0.73 (0.65-0.83) | 0.13 (0.10-0.15) | 0.25 (0.22-0.29) | 0.47 (0.39-0.57) | - | 0.14 (0.12-0.17) | - | - |
| COVID-19 death | 3-6 | 0.05 (0.02-0.13) | - | - | - | - | - | - | - | - | - |
| | 7-10 | - | - | - | - | - | - | - | - | - | - |
| | 11-14 | 0.06 (0.03-0.12) | 0.10 (0.06-0.16) | 0.51 (0.23-1.12) | - | 0.08 (0.05-0.12) | - | - | - | - | - |
| | 15-18 | 0.07 (0.04-0.10) | 0.14 (0.10-0.20) | 0.35 (0.21-0.58) | 0.04 (0.02-0.09) | 0.09 (0.06-0.14) | 0.45 (0.19-1.05) | - | 0.03 (0.01-0.08) | - | - |
| | 19-22 | 0.11 (0.09-0.15) | 0.14 (0.11-0.19) | 0.67 (0.50-0.90) | 0.09 (0.05-0.16) | 0.10 (0.07-0.15) | 0.91 (0.49-1.71) | - | 0.06 (0.03-0.11) | - | - |
| | 23-26 | 0.12 (0.09-0.16) | 0.17 (0.12-0.23) | 0.70 (0.53-0.94) | 0.08 (0.04-0.14) | 0.13 (0.08-0.19) | 0.59 (0.32-1.07) | - | 0.08 (0.04-0.16) | - | - |
| Positive SARS-CoV-2 test | 3-6 | 0.19 (0.14-0.26) | 0.47 (0.38-0.59) | 0.62 (0.47-0.81) | 0.19 (0.15-0.22) | 0.66 (0.61-0.70) | 0.35 (0.29-0.41) | 0.27 (0.23-0.31) | 0.79 (0.76-0.82) | 0.37 (0.33-0.43) | 0.24 (0.23-0.25) |
| | 7-10 | 0.35 (0.30-0.41) | 0.66 (0.59-0.73) | 0.54 (0.47-0.62) | 0.34 (0.32-0.36) | 0.68 (0.66-0.71) | 0.50 (0.47-0.53) | 0.46 (0.41-0.51) | 0.93 (0.90-0.95) | 0.49 (0.44-0.54) | 0.41 (0.40-0.42) |
| | 11-14 | 0.43 (0.39-0.48) | 0.70 (0.65-0.76) | 0.61 (0.57-0.66) | 0.42 (0.39-0.44) | 0.81 (0.78-0.83) | 0.59 (0.56-0.62) | 0.73 (0.68-0.79) | 1.21 (1.18-1.24) | 0.56 (0.51-0.60) | 0.59 (0.58-0.61) |
| | 15-18 | 0.58 (0.53-0.63) | 0.86 (0.80-0.91) | 0.69 (0.66-0.73) | 0.58 (0.55-0.61) | 1.01 (0.97-1.04) | 0.65 (0.62-0.67) | 0.95 (0.89-1.02) | 1.51 (1.47-1.54) | 0.63 (0.59-0.67) | 0.95 (0.92-0.97) |
| | 19-22 | 0.64 (0.60-0.70) | 1.01 (0.95-1.08) | 0.70 (0.66-0.73) | 0.82 (0.78-0.86) | 1.26 (1.22-1.30) | 0.69 (0.66-0.72) | 1.19 (1.12-1.27) | 1.77 (1.73-1.81) | 0.67 (0.64-0.71) | 1.35 (1.31-1.40) |
| | 23-26 | 0.85 (0.78-0.92) | 1.21 (1.13-1.30) | 0.76 (0.73-0.79) | 0.96 (0.92-1.01) | 1.45 (1.40-1.50) | 0.72 (0.69-0.74) | 1.03 (0.93-1.15) | 1.99 (1.94-2.05) | 0.60 (0.55-0.67) | 1.53 (1.07-2.18) |
| Non-COVID-19 death | 3-6 | 0.32 (0.28-0.36) | 0.48 (0.42-0.55) | 0.79 (0.70-0.89) | 0.39 (0.30-0.50) | 0.35 (0.28-0.43) | 1.21 (0.96-1.53) | - | 0.34 (0.24-0.47) | - | - |
| | 7-10 | 0.41 (0.37-0.45) | 0.52 (0.46-0.59) | 0.89 (0.81-0.98) | 0.36 (0.29-0.46) | 0.38 (0.32-0.46) | 0.98 (0.79-1.20) | - | 0.42 (0.32-0.56) | - | - |
| | 11-14 | 0.46 (0.42-0.51) | 0.55 (0.49-0.62) | 0.91 (0.83-0.99) | 0.40 (0.31-0.50) | 0.44 (0.37-0.53) | 0.89 (0.73-1.08) | - | 0.57 (0.43-0.75) | - | - |
| | 15-18 | 0.50 (0.45-0.55) | 0.59 (0.53-0.67) | 0.87 (0.80-0.95) | 0.46 (0.37-0.58) | 0.55 (0.46-0.66) | 0.99 (0.82-1.19) | - | 0.56 (0.43-0.73) | - | - |
| | 19-22 | 0.51 (0.46-0.57) | 0.63 (0.56-0.71) | 0.88 (0.81-0.95) | 0.42 (0.33-0.53) | 0.51 (0.43-0.62) | 0.81 (0.67-0.99) | 1.10 (0.52-2.35) | 0.58 (0.44-0.76) | 2.67 (1.30-5.50) | - |
| | 23-26 | 0.57 (0.51-0.64) | 0.66 (0.58-0.76) | 0.97 (0.89-1.05) | 0.61 (0.47-0.79) | 0.65 (0.52-0.80) | 1.01 (0.84-1.20) | - | 0.59 (0.44-0.79) | - | - |
| Any SARS-CoV-2 test | 3-6 | 0.88 (0.87-0.90) | 0.84 (0.82-0.85) | 0.74 (0.73-0.75) | 1.40 (1.38-1.42) | 1.26 (1.25-1.28) | 0.71 (0.71-0.72) | 1.73 (1.69-1.78) | 1.40 (1.39-1.41) | 0.79 (0.77-0.81) | 1.56 (1.55-1.58) |
| | 7-10 | 1.29 (1.27-1.31) | 1.34 (1.32-1.37) | 0.90 (0.89-0.91) | 1.76 (1.73-1.78) | 1.70 (1.68-1.72) | 0.96 (0.95-0.97) | 1.96 (1.91-2.01) | 2.18 (2.16-2.19) | 0.86 (0.84-0.87) | 1.94 (1.93-1.96) |
| | 11-14 | 1.59 (1.55-1.62) | 1.84 (1.80-1.88) | 0.94 (0.93-0.95) | 1.82 (1.79-1.85) | 1.84 (1.82-1.86) | 1.03 (1.02-1.04) | 2.17 (2.12-2.23) | 2.59 (2.57-2.61) | 0.86 (0.84-0.88) | 2.12 (2.10-2.14) |
| | 15-18 | 1.92 (1.88-1.96) | 2.51 (2.45-2.57) | 0.94 (0.93-0.95) | 1.73 (1.70-1.75) | 1.94 (1.91-1.96) | 0.98 (0.97-0.99) | 2.35 (2.30-2.41) | 3.31 (3.28-3.34) | 0.87 (0.86-0.89) | 2.54 (2.52-2.56) |
| | 19-22 | 2.51 (2.44-2.57) | 3.77 (3.66-3.87) | 0.95 (0.94-0.96) | 2.11 (2.08-2.14) | 2.49 (2.46-2.53) | 1.00 (0.99-1.01) | 2.59 (2.53-2.65) | 4.85 (4.79-4.91) | 0.87 (0.85-0.89) | 3.78 (3.73-3.84) |
| | 23-26 | 3.94 (3.82-4.07) | 6.59 (6.37-6.82) | 1.08 (1.07-1.09) | 2.99 (2.94-3.05) | 3.80 (3.73-3.87) | 1.09 (1.08-1.10) | 3.43 (3.31-3.56) | 8.95 (8.80-9.09) | 1.08 (1.05-1.11) | 7.05 (6.33-7.84) |

[a] And not clinically vulnerable

**Supplementary Table 8:** Ratio of adjusted hazard ratios for each comparison period.

| | 65+ years | | | 16-64 years and clinically vulnerable | | | 40-64 years[a] | | | 18-39 years[a] | | |
|---|---|---|---|---|---|---|---|---|---|---|---|---|
| **Outcome** | **BNT162b2 vs unvaccinated** | **ChAdOx1 vs unvaccinated** | **BNT162b2 vs ChAdOx1** | **BNT162b2 vs unvaccinated** | **ChAdOx1 vs unvaccinated** | **BNT162b2 vs ChAdOx1** | **BNT162b2 vs unvaccinated** | **ChAdOx1 vs unvaccinated** | **BNT162b2 vs ChAdOx1** | **BNT162b2 vs unvaccinated** | **ChAdOx1 vs unvaccinated** | **BNT162b2 vs ChAdOx1** |
| COVID-19 hospitalisation | 1.23 (1.15-1.32) | 1.20 (1.14-1.27) | 1.09 (1.01-1.18) | 1.34 (1.20-1.48) | 1.25 (1.19-1.30) | 1.07 (0.98-1.16) | - | 1.27 (1.16-1.38) | - | 1.39 (1.15-1.68) | - | - |
| COVID-19 death | 1.26 (1.08-1.48) | 1.16 (0.98-1.37) | 1.23 (0.96-1.58) | 1.30 (0.77-2.18) | 1.18 (0.97-1.43) | 1.10 (0.55-2.19) | - | 1.53 (0.89-2.64) | - | - | - | - |
| Positive SARS-CoV-2 test | 1.27 (1.20-1.34) | 1.18 (1.15-1.21) | 1.07 (1.04-1.10) | 1.36 (1.29-1.43) | 1.19 (1.16-1.22) | 1.13 (1.07-1.19) | 1.33 (1.18-1.49) | 1.21 (1.19-1.24) | 1.10 (1.04-1.17) | 1.52 (1.47-1.58) | - | - |
| Non-COVID-19 death | 1.11 (1.07-1.15) | 1.07 (1.03-1.10) | 1.02 (1.00-1.05) | 1.08 (1.02-1.15) | 1.13 (1.07-1.18) | 0.97 (0.91-1.03) | - | 1.11 (1.03-1.19) | - | - | - | - |
| Any SARS-CoV-2 test | 1.32 (1.27-1.37) | 1.48 (1.42-1.54) | 1.06 (1.03-1.09) | 1.13 (1.07-1.20) | 1.21 (1.14-1.28) | 1.06 (1.01-1.12) | 1.13 (1.10-1.16) | 1.41 (1.32-1.50) | 1.05 (1.02-1.08) | 1.32 (1.20-1.44) | - | - |

[a] And not clinically vulnerable

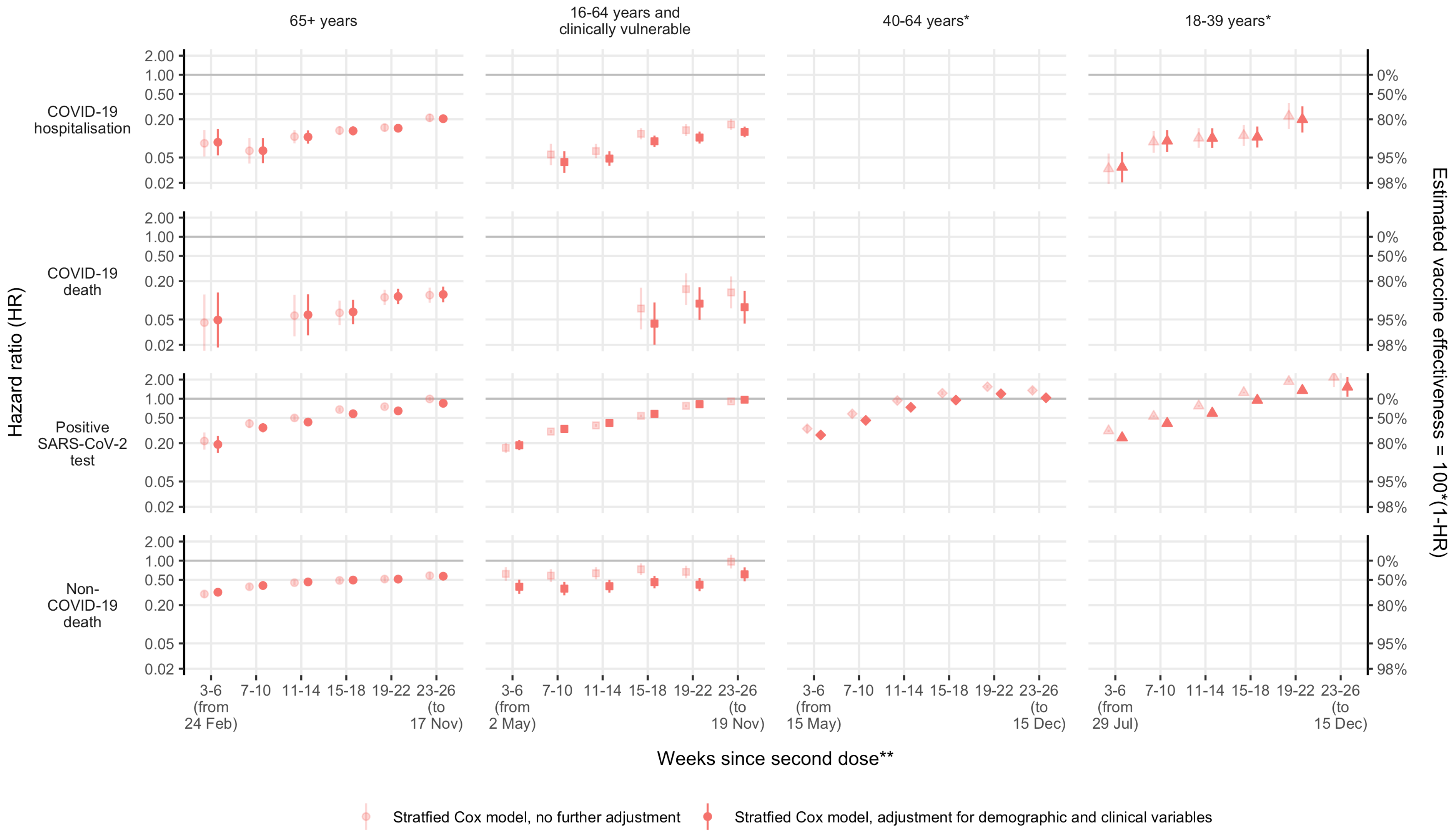

**Supplementary Figure 19:** Unadjusted and adjusted hazard ratios for BNT162b2 vs unvaccinated

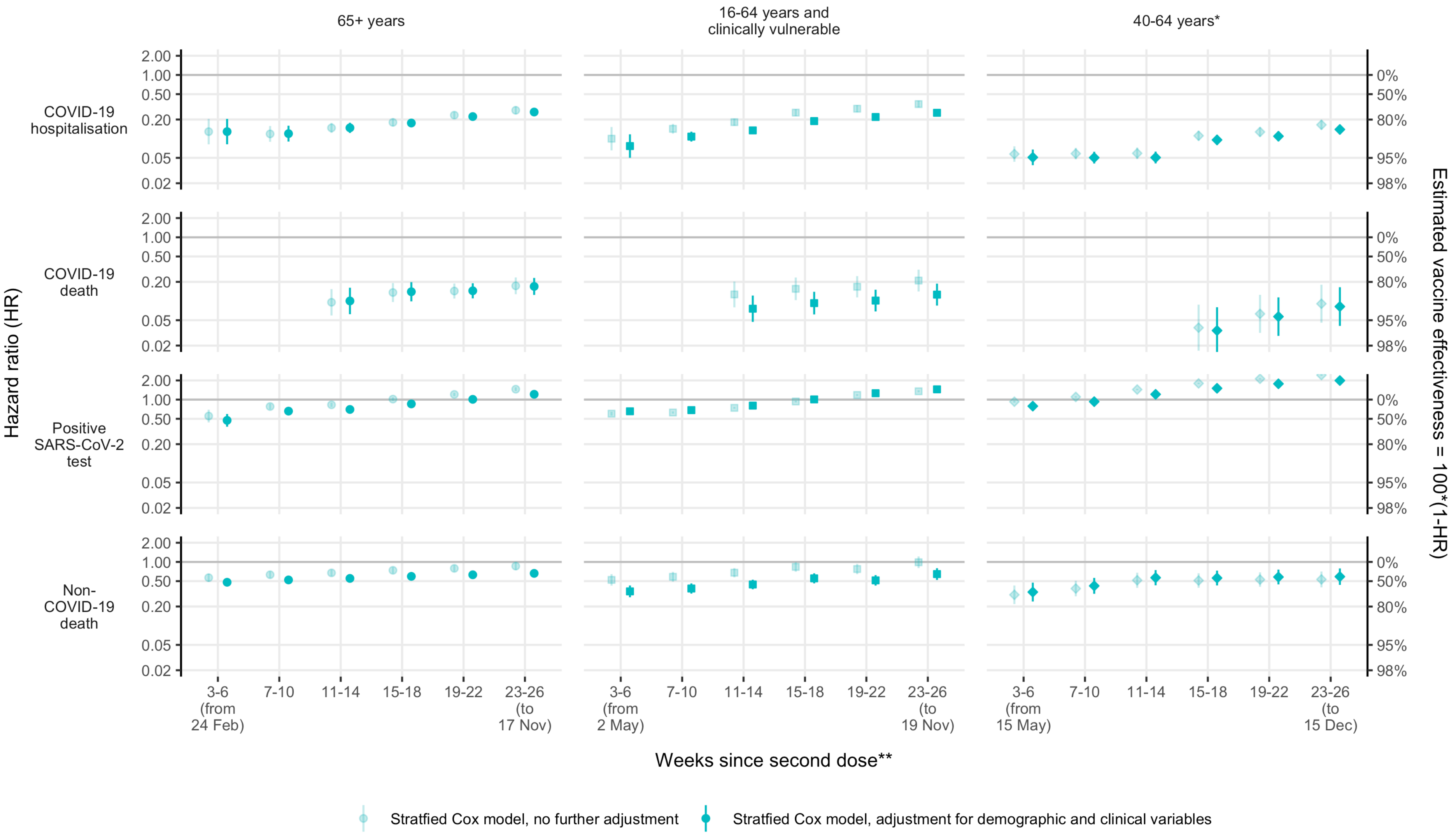

**Supplementary Figure 20:** Unadjusted and adjusted hazard ratios for ChAdOx1 vs unvaccinated

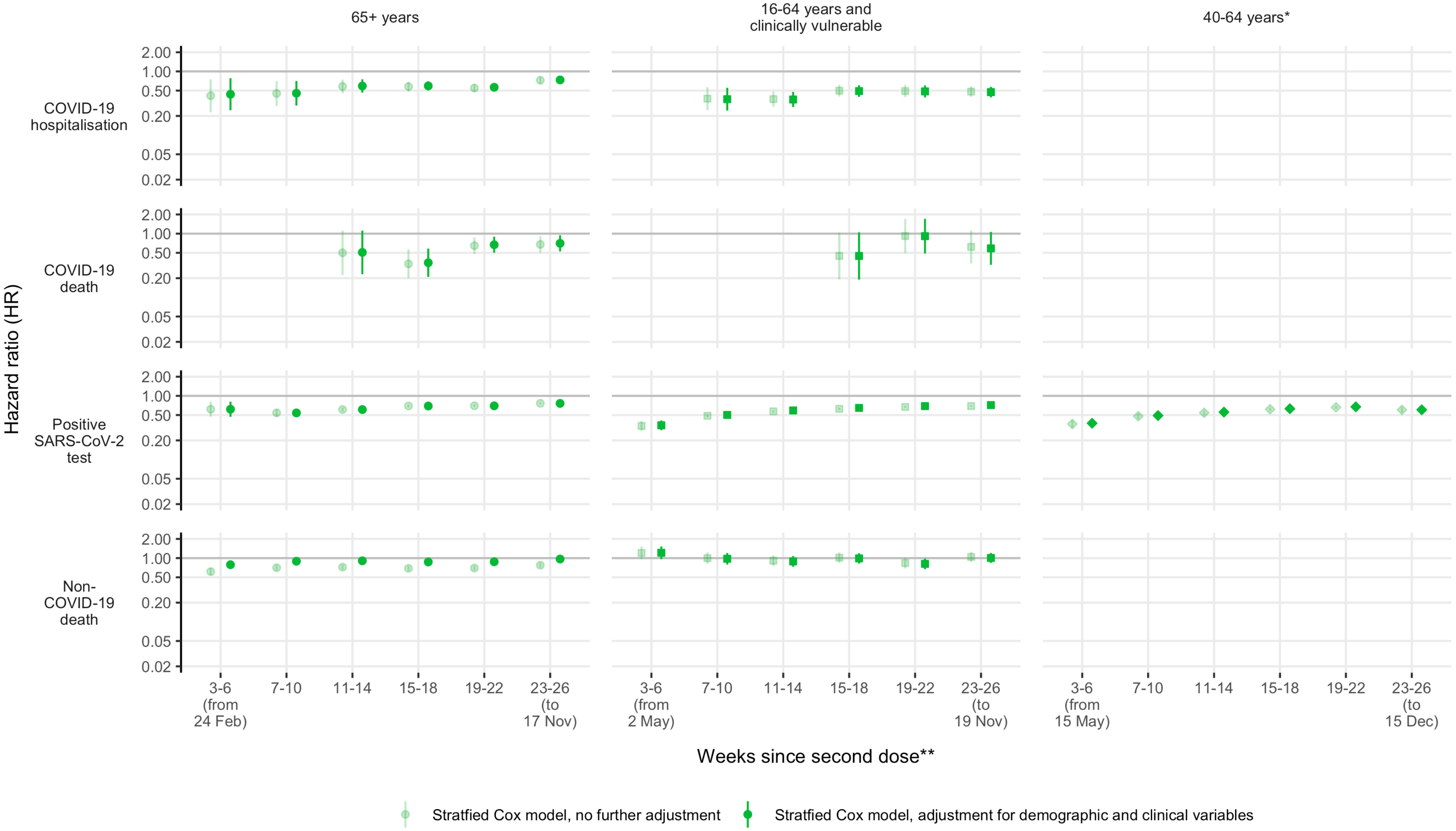

**Supplementary Figure 21:** Unadjusted and adjusted hazard ratios for BNT162b2 vs ChAdOx1

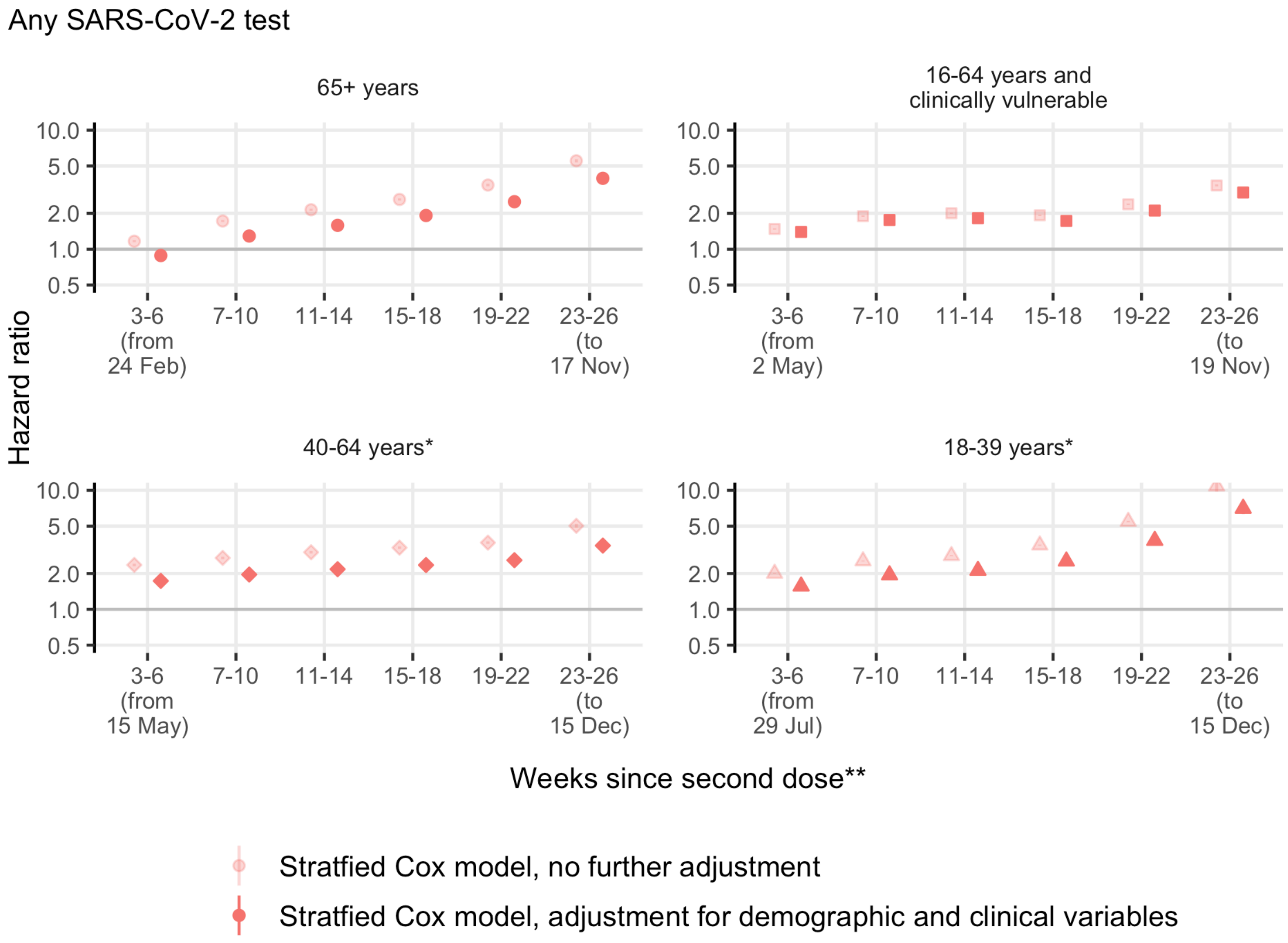

**Supplementary Figure 22:** Unadjusted and adjusted hazard ratios for any SARS-CoV-2 test for BNT162b2 vs unvaccinated

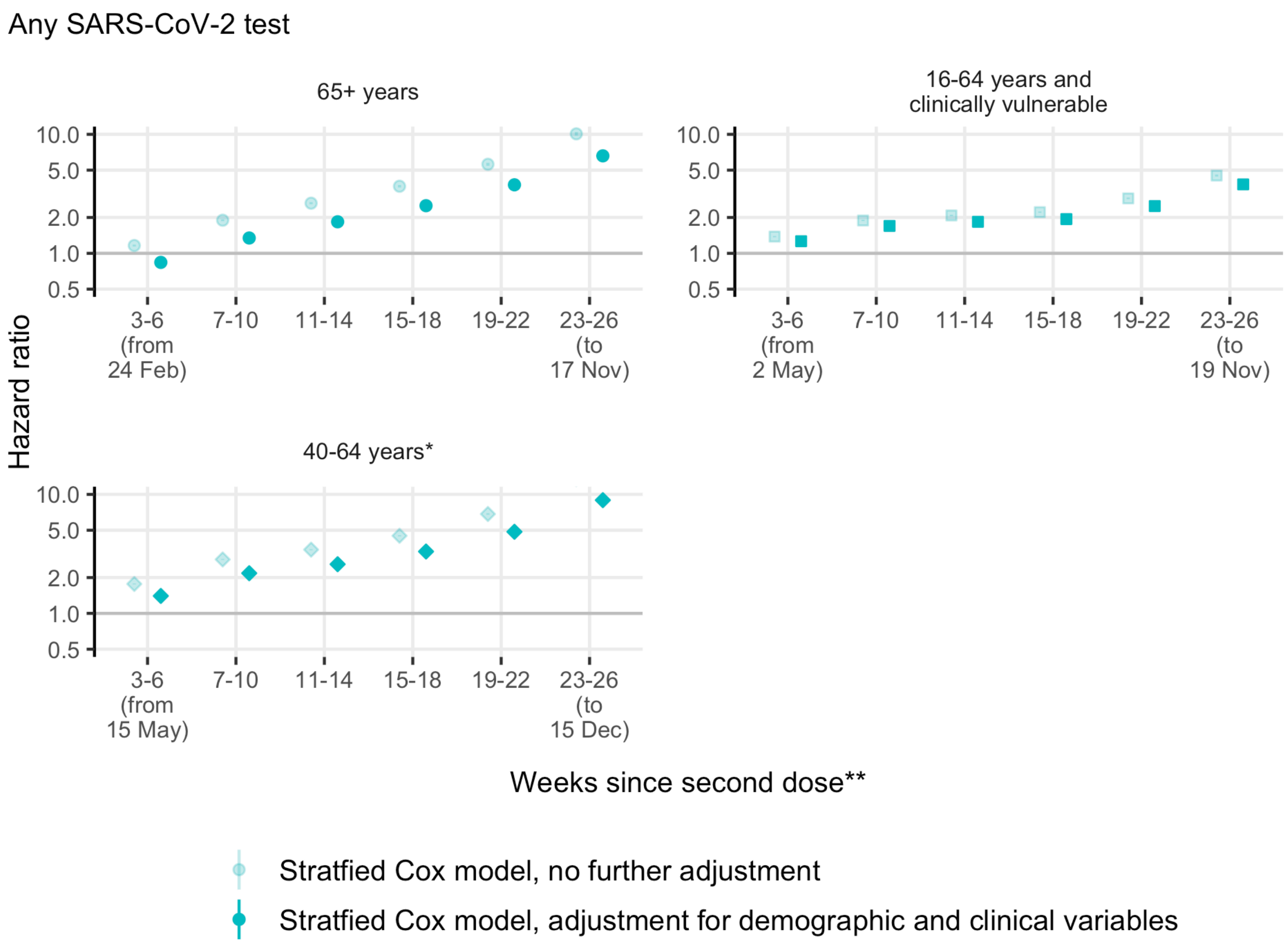

**Supplementary Figure 23:** Unadjusted and adjusted hazard ratios for any SARS-CoV-2 test for ChAdOx1 vs unvaccinated

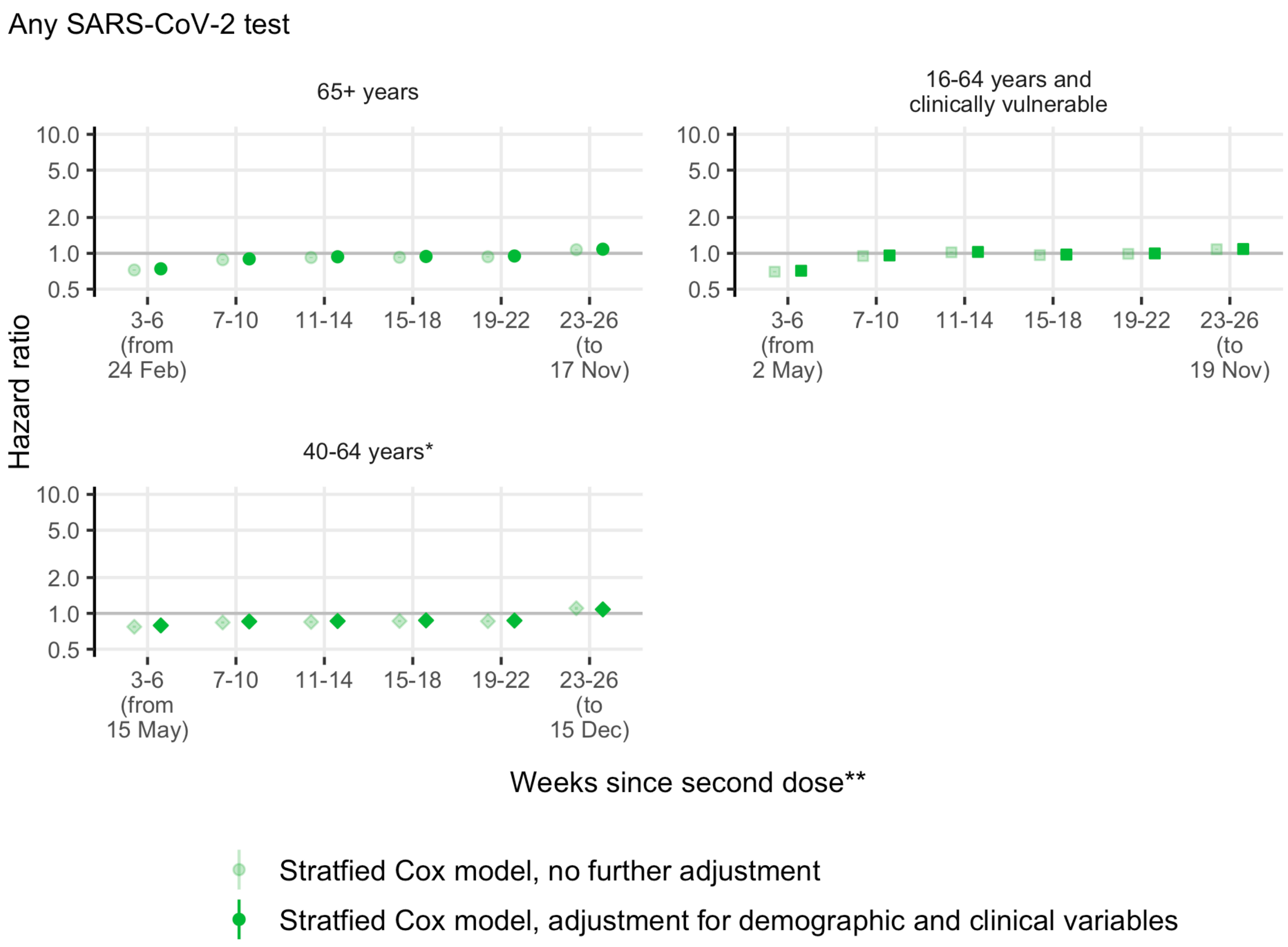

**Supplementary Figure 24:** Unadjusted and adjusted hazard ratios for any SARS-CoV-2 test for BNT162b2 vs ChAdOx1

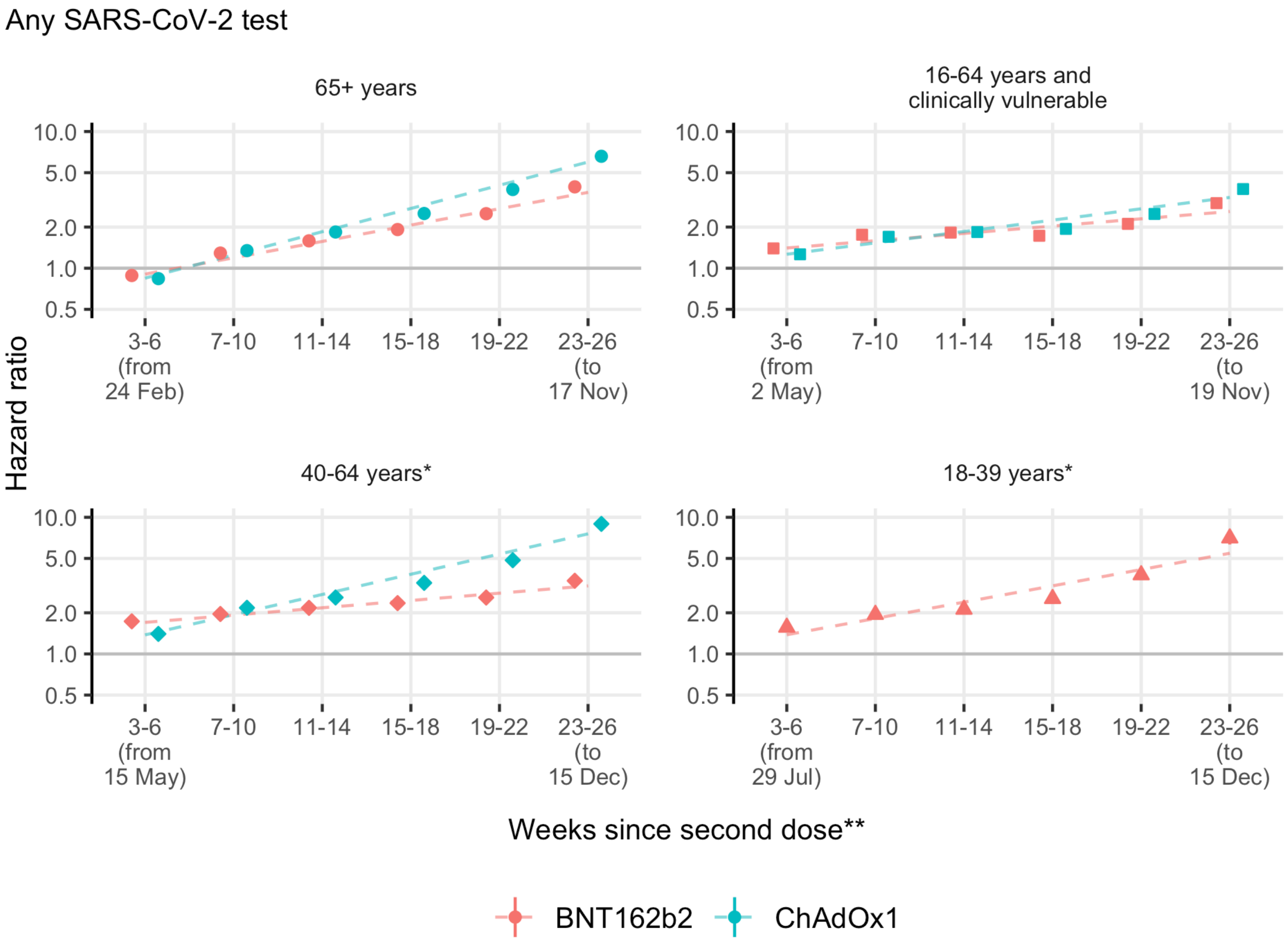

**Supplementary Figure 25:** Adjusted hazard ratios for any SARS-CoV-2 test for BNT162b2 and ChAdOx1 vs unvaccinated

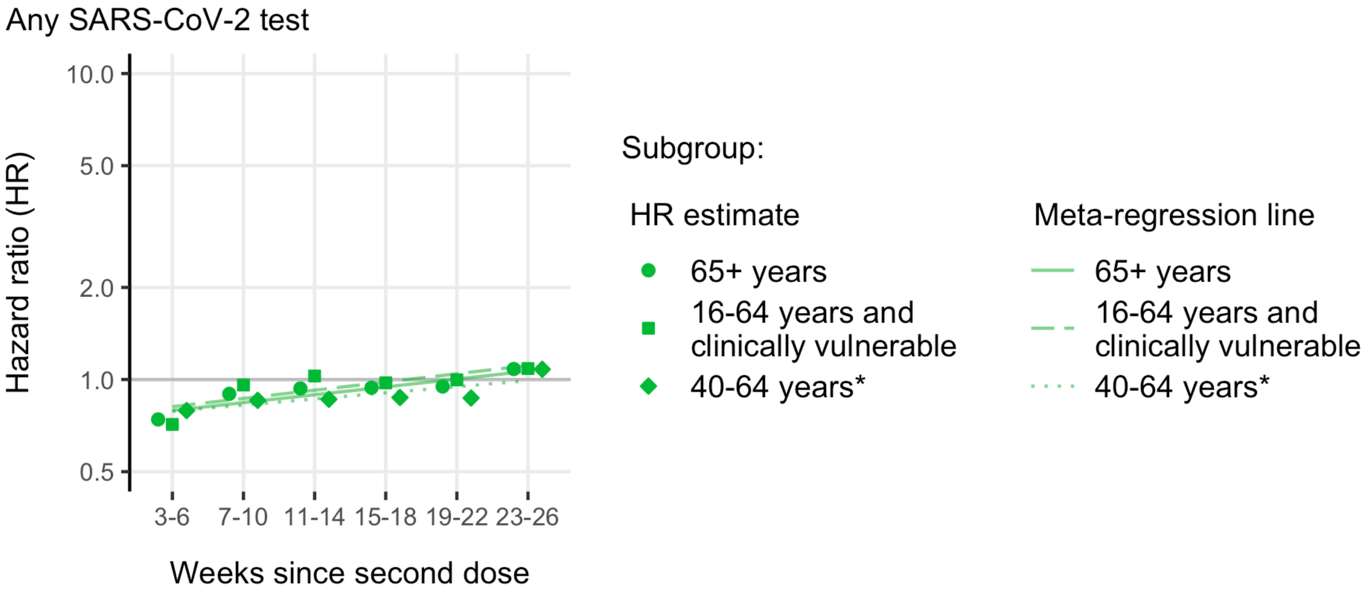

**Supplementary Figure 26:** Adjusted hazard ratios for any SARS-CoV-2 test for BNT162b2 vs ChAdOx1

**Supplementary Table 9:** Hazard ratios corresponding to covariates in the adjusted BNT162b2 vs unvaccinated model in the 65+ years subgroup.

| Variable | Category[b] | COVID-19 hospitalisation | COVID-19 death | Positive SARS-CoV-2 test | Non-COVID-19 death | Any SARS-CoV-2 test |
|---|---|---|---|---|---|---|
| Age[a] | age_65to70 | 1.06 (1.00-1.12) | 1.01 (0.88-1.16) | 0.96 (0.94-0.99) | 1.08 (1.01-1.15) | 0.95 (0.94-0.96) |
| | age_65to75 | 0.98 (0.95-1.01) | 0.96 (0.90-1.03) | 0.97 (0.96-0.98) | 1.01 (0.99-1.03) | 0.97 (0.97-0.98) |
| | age_75to80 | 0.99 (0.91-1.08) | 1.00 (0.83-1.19) | 0.95 (0.91-1.00) | 1.07 (1.02-1.13) | 0.96 (0.95-0.97) |
| | age_80plus | 1.41 (0.87-2.31) | 1.11 (0.53-2.35) | 0.84 (0.61-1.17) | 0.98 (0.83-1.17) | 0.66 (0.62-0.71) |
| | age_80plus_squared | 1.00 (1.00-1.00) | 1.00 (1.00-1.00) | 1.00 (1.00-1.00) | 1.00 (1.00-1.00) | 1.00 (1.00-1.00) |
| Sex | Female | 1.00 (ref) | - | 1.00 (ref) | 1.00 (ref) | 1.00 (ref) |
| | Male | 1.31 (1.21-1.42) | - | 1.14 (1.10-1.18) | 1.26 (1.20-1.31) | 1.00 (0.99-1.01) |
| IMD | 1 most deprived | 1.00 (ref) | 1.00 (ref) | 1.00 (ref) | 1.00 (ref) | 1.00 (ref) |
| | 2 | 0.85 (0.76-0.95) | - | 0.97 (0.92-1.03) | 0.94 (0.87-1.01) | 1.11 (1.09-1.13) |
| | 3 | 0.76 (0.68-0.86) | - | 0.89 (0.84-0.94) | 0.84 (0.79-0.91) | 1.19 (1.17-1.21) |
| | 4 | - | - | 0.90 (0.85-0.96) | 0.86 (0.80-0.93) | 1.25 (1.23-1.28) |
| | 5 least deprived | - | - | 0.89 (0.84-0.95) | 0.80 (0.75-0.87) | 1.34 (1.32-1.36) |
| | 4 / 5 least deprived | 0.67 (0.60-0.75) | - | - | - | - |
| | 2 / 3 / 4 / 5 least deprived | - | 0.70 (0.58-0.85) | - | - | - |
| Ethnicity | White | 1.00 (ref) | 1.00 (ref) | 1.00 (ref) | 1.00 (ref) | 1.00 (ref) |
| | Black | - | - | - | 0.58 (0.48-0.72) | 0.81 (0.77-0.85) |
| | South Asian | - | - | - | 0.49 (0.42-0.57) | 0.62 (0.60-0.64) |
| | Mixed | - | - | - | - | 0.80 (0.74-0.86) |
| | Other | - | - | - | - | 0.63 (0.60-0.67) |
| | Black / South Asian / Mixed / Other | 1.12 (0.99-1.27) | 1.65 (1.31-2.08) | 1.07 (0.99-1.15) | - | - |
| | Mixed / Other | - | - | - | 0.61 (0.49-0.76) | - |
| BMI | Not obese | 1.00 (ref) | 1.00 (ref) | 1.00 (ref) | 1.00 (ref) | 1.00 (ref) |
| | Obese I (30-34.9) | - | - | 1.20 (1.15-1.26) | 0.76 (0.72-0.81) | 0.94 (0.93-0.95) |
| | Obese II (35-39.9) | - | - | 1.20 (1.12-1.29) | 0.78 (0.70-0.86) | 0.90 (0.88-0.92) |
| | Obese III (40+) | - | - | 1.23 (1.11-1.37) | 0.83 (0.72-0.97) | 0.85 (0.82-0.88) |
| | Obese I (30-34.9) / Obese II (35-39.9) / Obese III (40+) | 1.31 (1.20-1.43) | 1.63 (1.37-1.94) | - | - | - |
| History of | Chronic respiratory disease | 1.17 (1.03-1.32) | 1.68 (1.36-2.08) | 1.08 (1.00-1.16) | 1.55 (1.45-1.65) | 1.00 (0.98-1.02) |
| | Chronic heart disease | 1.03 (0.93-1.15) | - | 1.05 (0.98-1.11) | 1.28 (1.21-1.36) | 1.15 (1.13-1.17) |
| | Chronic liver disease | - | - | 0.96 (0.87-1.07) | 1.34 (1.20-1.49) | 1.10 (1.07-1.13) |
| | Chronic kidney disease | 1.06 (0.94-1.18) | - | 1.02 (0.96-1.09) | 1.08 (1.02-1.14) | 1.09 (1.07-1.11) |
| | Chronic neurological disease | 1.13 (1.01-1.27) | - | 1.02 (0.95-1.10) | 1.19 (1.12-1.26) | 1.13 (1.11-1.15) |
| | Diabetes | 1.11 (1.00-1.23) | - | 1.04 (0.98-1.11) | 1.12 (1.06-1.18) | 0.99 (0.98-1.01) |
| | Immunosuppression | - | - | 1.08 (0.99-1.18) | 1.25 (1.15-1.35) | 1.12 (1.10-1.15) |
| | Learning disability | - | - | - | - | 1.21 (1.06-1.39) |
| | Serious mental illness | - | - | - | 1.37 (1.16-1.62) | 1.07 (1.02-1.12) |
| Shielding criteria met | yes | 1.82 (1.64-2.02) | - | 1.30 (1.23-1.37) | 1.82 (1.72-1.93) | 1.28 (1.26-1.30) |
| Morbidity count | 0 | 1.00 (ref) | 1.00 (ref) | 1.00 (ref) | 1.00 (ref) | 1.00 (ref) |
| | 1 | 1.74 (1.53-1.98) | - | 1.07 (1.00-1.14) | 1.49 (1.38-1.61) | 0.90 (0.89-0.92) |
| | 2+ | 2.51 (2.08-3.03) | - | 1.13 (1.00-1.27) | 1.70 (1.51-1.90) | 0.85 (0.82-0.87) |
| | 1 / 2+ | - | 3.26 (2.57-4.15) | - | - | - |
| Flu vaccine in previous 5 years | yes | - | - | 1.31 (1.23-1.38) | 0.93 (0.86-1.00) | 1.49 (1.46-1.51) |
| Resident in long-term residential home | yes | - | - | - | 5.68 (4.82-6.69) | 7.36 (6.92-7.83) |
| Housebound | yes | - | - | 1.20 (1.05-1.37) | 2.37 (2.20-2.55) | 1.28 (1.23-1.32) |
| Number of SARS-CoV-2 tests | 0 | 1.00 (ref) | 1.00 (ref) | 1.00 (ref) | 1.00 (ref) | 1.00 (ref) |
| | 1 | 1.23 (1.08-1.40) | - | 1.42 (1.34-1.50) | 1.23 (1.15-1.32) | 1.51 (1.49-1.53) |
| | 2 | - | - | - | 1.57 (1.42-1.73) | 1.70 (1.66-1.74) |
| | 3+ | - | - | - | 2.54 (2.35-2.75) | 2.79 (2.72-2.85) |
| | 2 / 3+ | 1.74 (1.52-1.99) | - | 1.39 (1.29-1.50) | - | - |
| | 1 / 2 / 3+ | - | 1.63 (1.33-2.00) | - | - | - |

[a] Categories separated by " / " were merged due to low event counts. [b] Age terms were fit separately within strata. The age range in the strata is indicated by the suffix following the underscore on the age category label. Strata in which the age range was < 5 years included only a linear age term, otherwise a quadratic age term was included.

**Supplementary Table 10:** Hazard ratios corresponding to covariates in the adjusted ChAdOx1 vs unvaccinated model in the 65+ years subgroup.

| Variable | Category[b] | COVID-19 hospitalisation | COVID-19 death | Positive SARS-CoV-2 test | Non-COVID-19 death | Any SARS-CoV-2 test |
|---|---|---|---|---|---|---|
| Age[a] | age_65to70 | 1.05 (1.00-1.10) | 1.04 (0.92-1.17) | 0.95 (0.93-0.96) | 1.11 (1.05-1.16) | 0.93 (0.93-0.94) |
| | age_65to75 | 1.02 (1.00-1.04) | 0.94 (0.90-0.99) | 0.96 (0.96-0.97) | 1.04 (1.03-1.06) | 0.97 (0.97-0.97) |
| | age_75to80 | 0.93 (0.85-1.01) | 0.92 (0.77-1.10) | 0.94 (0.89-0.99) | 1.02 (0.96-1.07) | 0.97 (0.96-0.99) |
| | age_80plus | 1.38 (0.68-2.79) | 1.28 (0.46-3.55) | 1.00 (0.62-1.62) | 0.94 (0.72-1.22) | 0.99 (0.88-1.11) |
| | age_80plus_squared | 1.00 (0.99-1.00) | 1.00 (0.99-1.00) | 1.00 (1.00-1.00) | 1.00 (1.00-1.00) | 1.00 (1.00-1.00) |
| Sex | Female | 1.00 (ref) | - | 1.00 (ref) | 1.00 (ref) | 1.00 (ref) |
| | Male | 1.31 (1.22-1.40) | - | 1.08 (1.05-1.11) | 1.29 (1.23-1.35) | 0.98 (0.98-0.99) |
| IMD | 1 most deprived | 1.00 (ref) | 1.00 (ref) | 1.00 (ref) | 1.00 (ref) | 1.00 (ref) |
| | 2 | 0.88 (0.79-0.97) | 0.72 (0.57-0.90) | 0.95 (0.91-0.99) | 0.94 (0.87-1.01) | 1.13 (1.11-1.14) |
| | 3 | 0.77 (0.69-0.85) | - | 0.93 (0.89-0.97) | 0.82 (0.77-0.89) | 1.21 (1.19-1.22) |
| | 4 | 0.72 (0.65-0.81) | - | 0.92 (0.88-0.96) | 0.84 (0.78-0.90) | 1.32 (1.30-1.34) |
| | 5 least deprived | 0.61 (0.53-0.69) | - | 0.91 (0.88-0.96) | 0.79 (0.73-0.85) | 1.43 (1.41-1.45) |
| | 3 / 4 / 5 least deprived | - | 0.57 (0.47-0.69) | - | - | - |
| Ethnicity | White | 1.00 (ref) | 1.00 (ref) | 1.00 (ref) | 1.00 (ref) | 1.00 (ref) |
| | Black | - | - | - | - | 0.84 (0.80-0.88) |
| | South Asian | - | - | - | - | 0.60 (0.58-0.62) |
| | Mixed | - | - | - | - | 0.81 (0.76-0.86) |
| | Other | - | - | - | - | 0.67 (0.64-0.70) |
| | Black / South Asian / Mixed / Other | 1.19 (1.06-1.34) | 1.26 (1.00-1.57) | 1.13 (1.06-1.19) | 0.55 (0.49-0.62) | - |
| BMI | Not obese | 1.00 (ref) | 1.00 (ref) | 1.00 (ref) | 1.00 (ref) | 1.00 (ref) |
| | Obese I (30-34.9) | 1.51 (1.39-1.65) | - | 1.24 (1.20-1.28) | 0.75 (0.70-0.80) | 0.94 (0.93-0.95) |
| | Obese II (35-39.9) | - | - | 1.28 (1.22-1.34) | 0.80 (0.73-0.88) | 0.90 (0.88-0.91) |
| | Obese III (40+) | - | - | 1.29 (1.20-1.38) | 0.97 (0.87-1.09) | 0.86 (0.84-0.88) |
| | Obese II (35-39.9) / Obese III (40+) | 1.74 (1.58-1.92) | - | - | - | - |
| | Obese I (30-34.9) / Obese II (35-39.9) / Obese III (40+) | - | 1.92 (1.63-2.25) | - | - | - |
| History of | Chronic respiratory disease | 1.35 (1.22-1.50) | - | 1.10 (1.04-1.16) | 1.67 (1.56-1.79) | 0.96 (0.94-0.98) |
| | Chronic heart disease | 1.16 (1.06-1.26) | - | 1.12 (1.07-1.17) | 1.29 (1.21-1.37) | 1.12 (1.10-1.14) |
| | Chronic liver disease | - | - | 1.13 (1.05-1.21) | 1.40 (1.27-1.55) | 1.11 (1.09-1.14) |
| | Chronic kidney disease | 1.11 (1.00-1.23) | - | 1.06 (1.01-1.12) | 1.03 (0.97-1.10) | 1.08 (1.06-1.10) |
| | Chronic neurological disease | 1.22 (1.10-1.36) | - | 1.05 (0.99-1.11) | 1.29 (1.21-1.38) | 1.10 (1.08-1.11) |
| | Diabetes | 1.26 (1.15-1.38) | 1.70 (1.43-2.02) | 1.08 (1.03-1.14) | 1.13 (1.06-1.20) | 0.98 (0.96-0.99) |
| | Immunosuppression | - | - | 1.06 (0.99-1.13) | 1.07 (0.98-1.17) | 1.11 (1.08-1.13) |
| | Learning disability | - | - | - | - | 1.47 (1.34-1.61) |
| | Serious mental illness | - | - | - | 1.30 (1.11-1.50) | 1.00 (0.96-1.04) |
| Shielding criteria met | yes | 1.90 (1.72-2.09) | - | 1.13 (1.08-1.19) | 2.04 (1.90-2.18) | 1.21 (1.20-1.23) |
| Morbidity count | 0 | 1.00 (ref) | 1.00 (ref) | 1.00 (ref) | 1.00 (ref) | 1.00 (ref) |
| | 1 | 1.66 (1.49-1.86) | - | 0.95 (0.91-1.00) | 1.74 (1.61-1.89) | 0.89 (0.88-0.91) |
| | 2+ | 2.18 (1.85-2.56) | - | 0.91 (0.83-1.00) | 2.17 (1.92-2.45) | 0.84 (0.82-0.87) |
| | 1 / 2+ | - | 3.51 (2.76-4.47) | - | - | - |
| Flu vaccine in previous 5 years | yes | 1.00 (0.92-1.09) | - | 1.38 (1.33-1.44) | 0.81 (0.76-0.87) | 1.55 (1.53-1.57) |
| Resident in long-term residential home | yes | - | - | - | 4.41 (3.65-5.33) | 4.87 (4.57-5.19) |
| Housebound | yes | 1.51 (1.30-1.75) | - | 0.89 (0.81-0.99) | 2.76 (2.56-2.97) | 1.03 (1.01-1.06) |
| Number of SARS-CoV-2 tests | 0 | 1.00 (ref) | 1.00 (ref) | 1.00 (ref) | 1.00 (ref) | 1.00 (ref) |
| | 1 | 1.31 (1.17-1.46) | - | 1.34 (1.28-1.39) | 1.34 (1.25-1.44) | 1.48 (1.46-1.50) |
| | 2 | - | - | 1.34 (1.24-1.44) | 1.61 (1.45-1.79) | 1.68 (1.64-1.71) |
| | 3+ | - | - | 1.29 (1.18-1.40) | 2.61 (2.41-2.83) | 2.53 (2.48-2.59) |
| | 2 / 3+ | 1.64 (1.45-1.85) | - | - | - | - |
| | 1 / 2 / 3+ | - | 1.64 (1.35-1.98) | - | - | - |

[a] Categories separated by " / " were merged due to low event counts. [b] Age terms were fit separately within strata. The age range in the strata is indicated by the suffix following the underscore on the age category label. Strata in which the age range was < 5 years included only a linear age term, otherwise a quadratic age term was included.

**Supplementary Table 11:** Hazard ratios corresponding to covariates in the adjusted BNT162b2 vs ChAdOx1 model in the 65+ years subgroup.

| Variable | Category[b] | COVID-19 hospitalisation | COVID-19 death | Positive SARS-CoV-2 test | Non-COVID-19 death | Any SARS-CoV-2 test |
|---|---|---|---|---|---|---|
| Age[a] | age_65to70 | 1.06 (0.99-1.13) | 1.03 (0.80-1.31) | 0.94 (0.93-0.96) | 1.11 (1.06-1.17) | 0.94 (0.93-0.94) |
| | age_65to75 | 1.02 (1.00-1.04) | 0.94 (0.89-0.99) | 0.97 (0.96-0.98) | 1.04 (1.03-1.05) | 0.97 (0.97-0.97) |
| | age_75to80 | 0.90 (0.80-1.01) | 0.83 (0.65-1.06) | 0.92 (0.88-0.96) | 1.07 (1.01-1.14) | 0.96 (0.95-0.97) |
| | age_80plus | 1.36 (0.77-2.41) | 0.70 (0.28-1.77) | 0.80 (0.57-1.13) | 1.00 (0.83-1.19) | 0.65 (0.61-0.69) |
| | age_80plus_squared | 1.00 (1.00-1.00) | 1.00 (1.00-1.01) | 1.00 (1.00-1.00) | 1.00 (1.00-1.00) | 1.00 (1.00-1.00) |
| Sex | Female | 1.00 (ref) | - | 1.00 (ref) | 1.00 (ref) | 1.00 (ref) |
| | Male | 1.46 (1.36-1.57) | - | 1.13 (1.10-1.15) | 1.29 (1.24-1.34) | 1.00 (0.99-1.01) |
| IMD | 1 most deprived | 1.00 (ref) | 1.00 (ref) | 1.00 (ref) | 1.00 (ref) | 1.00 (ref) |
| | 2 | 0.81 (0.73-0.90) | - | 0.95 (0.91-0.99) | 0.92 (0.86-0.98) | 1.13 (1.12-1.15) |
| | 3 | 0.66 (0.59-0.73) | - | 0.91 (0.87-0.94) | 0.82 (0.77-0.87) | 1.21 (1.20-1.23) |
| | 4 | - | - | 0.91 (0.88-0.95) | 0.84 (0.79-0.89) | 1.32 (1.30-1.33) |
| | 5 least deprived | - | - | 0.90 (0.87-0.94) | 0.78 (0.73-0.83) | 1.42 (1.40-1.43) |
| | 4 / 5 least deprived | 0.60 (0.55-0.66) | - | - | - | - |
| | 2 / 3 / 4 / 5 least deprived | - | 0.51 (0.42-0.62) | - | - | - |
| Ethnicity | White | 1.00 (ref) | 1.00 (ref) | 1.00 (ref) | 1.00 (ref) | 1.00 (ref) |
| | Black | - | - | - | - | 0.85 (0.81-0.89) |
| | South Asian | - | - | - | - | 0.62 (0.61-0.64) |
| | Mixed | - | - | - | - | 0.82 (0.78-0.87) |
| | Other | - | - | - | - | 0.70 (0.67-0.73) |
| | Black / South Asian / Mixed / Other | 1.44 (1.23-1.68) | 1.53 (1.07-2.17) | 1.15 (1.09-1.22) | 0.78 (0.70-0.87) | - |
| BMI | Not obese | 1.00 (ref) | 1.00 (ref) | 1.00 (ref) | 1.00 (ref) | 1.00 (ref) |
| | Obese I (30-34.9) | - | - | 1.21 (1.18-1.24) | 0.75 (0.71-0.79) | 0.94 (0.93-0.95) |
| | Obese II (35-39.9) | - | - | 1.24 (1.19-1.30) | 0.81 (0.75-0.88) | 0.89 (0.88-0.91) |
| | Obese III (40+) | - | - | 1.29 (1.21-1.37) | 0.99 (0.89-1.09) | 0.85 (0.83-0.87) |
| | Obese I (30-34.9) / Obese II (35-39.9) / Obese III (40+) | 1.48 (1.37-1.59) | 1.77 (1.48-2.11) | - | - | - |
| History of | Chronic respiratory disease | 1.27 (1.15-1.40) | 2.27 (1.86-2.77) | 1.11 (1.06-1.16) | 1.57 (1.49-1.65) | 0.97 (0.96-0.98) |
| | Chronic heart disease | 1.07 (0.98-1.17) | - | 1.12 (1.07-1.16) | 1.28 (1.22-1.35) | 1.13 (1.12-1.15) |
| | Chronic liver disease | - | - | 1.10 (1.03-1.17) | 1.36 (1.25-1.48) | 1.11 (1.09-1.13) |
| | Chronic kidney disease | 1.14 (1.04-1.25) | 1.70 (1.39-2.08) | 1.07 (1.02-1.12) | 1.05 (1.00-1.10) | 1.09 (1.07-1.10) |
| | Chronic neurological disease | 1.11 (1.01-1.23) | - | 1.01 (0.96-1.06) | 1.23 (1.17-1.29) | 1.10 (1.09-1.12) |
| | Diabetes | 1.18 (1.08-1.29) | - | 1.05 (1.01-1.10) | 1.11 (1.06-1.17) | 0.97 (0.96-0.99) |
| | Immunosuppression | - | - | 1.11 (1.05-1.18) | 1.13 (1.05-1.21) | 1.11 (1.10-1.13) |
| | Learning disability | - | - | - | - | 1.35 (1.24-1.47) |
| | Serious mental illness | - | - | - | 1.27 (1.09-1.47) | 0.96 (0.93-0.99) |
| Shielding criteria met | yes | 2.35 (2.14-2.58) | - | 1.18 (1.13-1.22) | 2.10 (1.99-2.21) | 1.26 (1.25-1.28) |
| Morbidity count | 0 | 1.00 (ref) | 1.00 (ref) | 1.00 (ref) | 1.00 (ref) | 1.00 (ref) |
| | 1 | 1.81 (1.59-2.05) | - | 0.92 (0.89-0.96) | 1.57 (1.47-1.68) | 0.88 (0.87-0.89) |
| | 2+ | 2.85 (2.41-3.37) | - | 0.89 (0.83-0.97) | 1.93 (1.75-2.12) | 0.83 (0.81-0.85) |
| | 1 / 2+ | - | 3.95 (2.84-5.50) | - | - | - |
| Flu vaccine in previous 5 years | yes | - | - | 1.42 (1.36-1.48) | 0.71 (0.67-0.76) | 1.55 (1.53-1.57) |
| Resident in long-term residential home | yes | - | - | - | 5.11 (4.43-5.88) | 6.57 (6.27-6.89) |
| Housebound | yes | - | - | 0.83 (0.75-0.92) | 2.60 (2.44-2.78) | 1.10 (1.07-1.12) |
| Number of SARS-CoV-2 tests | 0 | 1.00 (ref) | 1.00 (ref) | 1.00 (ref) | 1.00 (ref) | 1.00 (ref) |
| | 1 | 1.41 (1.27-1.56) | - | 1.33 (1.29-1.38) | 1.24 (1.18-1.32) | 1.46 (1.45-1.48) |
| | 2 | - | - | - | 1.63 (1.51-1.77) | 1.64 (1.62-1.67) |
| | 3+ | - | - | - | 2.64 (2.47-2.81) | 2.56 (2.51-2.60) |
| | 2 / 3+ | 1.89 (1.70-2.11) | - | 1.29 (1.23-1.36) | - | - |
| | 1 / 2 / 3+ | - | 1.71 (1.41-2.07) | - | - | - |

[a] Categories separated by " / " were merged due to low event counts. [b] Age terms were fit separately within strata. The age range in the strata is indicated by the suffix following the underscore on the age category label. Strata in which the age range was < 5 years included only a linear age term, otherwise a quadratic age term was included.

**Supplementary Table 12:** Hazard ratios corresponding to covariates in the adjusted BNT162b2 vs unvaccinated model in the 16-64 years and clinically vulnerable subgroup.

| Variable | Category[b] | COVID-19 hospitalisation | COVID-19 death | Positive SARS-CoV-2 test | Non-COVID-19 death | Any SARS-CoV-2 test |
|---|---|---|---|---|---|---|
| Age[a] | age_16to64 | 1.00 (0.98-1.02) | 1.14 (1.00-1.29) | 1.01 (1.01-1.02) | 1.10 (1.05-1.16) | 1.00 (1.00-1.00) |
| | age_16to64_squared | 1.00 (1.00-1.00) | 1.00 (1.00-1.00) | 1.00 (1.00-1.00) | 1.00 (1.00-1.00) | 1.00 (1.00-1.00) |
| Sex | Female | 1.00 (ref) | - | 1.00 (ref) | 1.00 (ref) | 1.00 (ref) |
| | Male | 0.96 (0.89-1.03) | - | 0.86 (0.84-0.88) | 1.52 (1.37-1.69) | 0.77 (0.76-0.78) |
| IMD | 1 most deprived | 1.00 (ref) | 1.00 (ref) | 1.00 (ref) | 1.00 (ref) | 1.00 (ref) |
| | 2 | - | - | 1.07 (1.03-1.10) | 0.83 (0.72-0.95) | 1.11 (1.10-1.13) |
| | 3 | - | - | 1.09 (1.05-1.12) | 0.79 (0.68-0.93) | 1.22 (1.20-1.24) |
| | 4 | - | - | 1.12 (1.08-1.16) | 0.85 (0.73-1.00) | 1.31 (1.29-1.33) |
| | 5 least deprived | - | - | 1.12 (1.08-1.16) | 0.73 (0.61-0.88) | 1.44 (1.41-1.46) |
| | 2 / 3 / 4 / 5 least deprived | 0.99 (0.92-1.07) | 0.98 (0.73-1.31) | - | - | - |
| Ethnicity | White | 1.00 (ref) | 1.00 (ref) | 1.00 (ref) | 1.00 (ref) | 1.00 (ref) |
| | Black | - | - | - | - | 0.94 (0.92-0.97) |
| | South Asian | - | - | - | - | 0.69 (0.67-0.70) |
| | Mixed | - | - | - | - | 0.92 (0.89-0.95) |
| | Other | - | - | - | - | 0.77 (0.75-0.80) |
| | Black / South Asian / Mixed / Other | 1.27 (1.17-1.39) | 1.40 (1.00-1.96) | 0.89 (0.87-0.92) | 0.56 (0.47-0.67) | - |
| BMI | Not obese | 1.00 (ref) | 1.00 (ref) | 1.00 (ref) | 1.00 (ref) | 1.00 (ref) |
| | Obese I (30-34.9) | - | - | 1.13 (1.10-1.16) | 0.64 (0.55-0.73) | 1.01 (1.00-1.02) |
| | Obese II (35-39.9) | - | - | 1.20 (1.15-1.25) | 0.66 (0.55-0.80) | 0.98 (0.97-1.00) |
| | Obese III (40+) | - | - | 1.19 (1.13-1.25) | 0.73 (0.61-0.87) | 0.99 (0.97-1.01) |
| | Obese I (30-34.9) / Obese II (35-39.9) / Obese III (40+) | 1.92 (1.78-2.07) | 3.11 (2.31-4.20) | - | - | - |
| History of | Chronic respiratory disease | - | - | 0.91 (0.86-0.97) | 1.56 (1.34-1.82) | 0.87 (0.85-0.89) |
| | Chronic heart disease | - | 1.31 (0.96-1.79) | 1.06 (1.00-1.11) | 1.42 (1.25-1.61) | 1.02 (1.00-1.04) |
| | Chronic liver disease | - | - | 0.92 (0.87-0.97) | 1.93 (1.67-2.22) | 1.02 (1.00-1.04) |
| | Chronic kidney disease | - | - | 1.08 (1.01-1.16) | 1.38 (1.15-1.65) | 1.10 (1.07-1.13) |
| | Chronic neurological disease | - | - | 0.96 (0.91-1.01) | 1.14 (0.98-1.32) | 0.99 (0.97-1.01) |
| | Diabetes | - | - | 1.08 (1.03-1.14) | 0.97 (0.84-1.12) | 0.96 (0.94-0.98) |
| | Immunosuppression | - | 1.43 (0.98-2.11) | 1.13 (1.07-1.19) | 0.90 (0.75-1.07) | 1.10 (1.07-1.12) |
| | Learning disability | - | - | 0.58 (0.53-0.63) | - | 0.91 (0.88-0.94) |
| | Serious mental illness | - | - | 0.57 (0.53-0.61) | 1.10 (0.89-1.36) | 0.78 (0.76-0.80) |
| Shielding criteria met | yes | 1.46 (1.32-1.62) | - | 0.92 (0.88-0.95) | 1.98 (1.64-2.37) | 0.90 (0.89-0.92) |
| Morbidity count | 0 | 1.00 (ref) | 1.00 (ref) | 1.00 (ref) | 1.00 (ref) | 1.00 (ref) |
| | 1 | - | - | 0.90 (0.84-0.96) | 0.70 (0.54-0.91) | 0.98 (0.95-1.00) |
| | 2+ | - | - | 0.87 (0.78-0.98) | 1.19 (0.87-1.63) | 0.95 (0.91-0.99) |
| | 1 / 2+ | 1.89 (1.53-2.33) | 2.75 (1.07-7.11) | - | - | - |
| Flu vaccine in previous 5 years | yes | - | - | 1.05 (1.02-1.07) | 0.96 (0.85-1.09) | 1.24 (1.23-1.26) |
| Resident in long-term residential home | yes | - | - | - | - | 2.53 (2.36-2.72) |
| Housebound | yes | - | - | - | 3.36 (2.61-4.34) | 0.98 (0.92-1.05) |
| Number of SARS-CoV-2 tests | 0 | 1.00 (ref) | 1.00 (ref) | 1.00 (ref) | 1.00 (ref) | 1.00 (ref) |
| | 1 | - | - | 1.49 (1.45-1.54) | 1.31 (1.13-1.51) | 1.47 (1.45-1.48) |
| | 2 | - | - | 1.61 (1.55-1.68) | 1.67 (1.37-2.04) | 1.78 (1.75-1.81) |
| | 3+ | - | - | 1.69 (1.62-1.76) | 4.77 (4.19-5.42) | 2.68 (2.64-2.73) |
| | 1 / 2 / 3+ | 1.69 (1.57-1.82) | 1.30 (0.96-1.75) | - | - | - |
| Pregnancy | yes | - | - | - | - | 1.97 (1.92-2.02) |

[a] Categories separated by ” / ” were merged due to low event counts. [b] Age terms were fit separately within strata. The age range in the strata is indicated by the suffix following the underscore on the age category label. Strata in which the age range was < 5 years included only a linear age term, otherwise a quadratic age term was included.

**Supplementary Table 13:** Hazard ratios corresponding to covariates in the adjusted ChAdOx1 vs unvaccinated model in the 16-64 years and clinically vulnerable subgroup.

| Variable | Category[b] | COVID-19 hospitalisation | COVID-19 death | Positive SARS-CoV-2 test | Non-COVID-19 death | Any SARS-CoV-2 test |
|---|---|---|---|---|---|---|
| Age[a] | age_16to64 | 1.02 (1.00-1.04) | 1.10 (1.01-1.20) | 1.02 (1.02-1.03) | 1.13 (1.08-1.19) | 1.02 (1.02-1.02) |
| | age_16to64_squared | 1.00 (1.00-1.00) | 1.00 (1.00-1.00) | 1.00 (1.00-1.00) | 1.00 (1.00-1.00) | 1.00 (1.00-1.00) |
| Sex | Female | 1.00 (ref) | 1.00 (ref) | 1.00 (ref) | 1.00 (ref) | 1.00 (ref) |
| | Male | 1.07 (1.00-1.13) | 1.56 (1.27-1.92) | 0.88 (0.87-0.90) | 1.42 (1.30-1.54) | 0.77 (0.76-0.78) |
| IMD | 1 most deprived | 1.00 (ref) | 1.00 (ref) | 1.00 (ref) | 1.00 (ref) | 1.00 (ref) |
| | 2 | - | - | 1.08 (1.05-1.10) | 0.93 (0.83-1.04) | 1.13 (1.12-1.14) |
| | 3 | - | - | 1.10 (1.07-1.13) | 0.84 (0.74-0.95) | 1.22 (1.21-1.24) |
| | 4 | - | - | 1.12 (1.09-1.15) | 0.76 (0.67-0.88) | 1.31 (1.30-1.33) |
| | 5 least deprived | - | - | 1.15 (1.12-1.18) | 0.80 (0.70-0.93) | 1.44 (1.43-1.46) |
| | 2 / 3 / 4 / 5 least deprived | 0.96 (0.90-1.03) | 0.94 (0.75-1.16) | - | - | - |
| Ethnicity | White | 1.00 (ref) | 1.00 (ref) | 1.00 (ref) | 1.00 (ref) | 1.00 (ref) |
| | Black | 1.32 (1.17-1.50) | - | 0.89 (0.85-0.93) | - | 0.92 (0.90-0.94) |
| | South Asian | 1.25 (1.14-1.39) | - | 0.85 (0.83-0.88) | - | 0.66 (0.65-0.67) |
| | Mixed | - | - | 0.97 (0.91-1.03) | - | 0.92 (0.89-0.94) |
| | Other | - | - | 0.70 (0.66-0.75) | - | 0.75 (0.73-0.77) |
| | Mixed / Other | 1.22 (1.06-1.40) | - | - | - | - |
| | Black / South Asian / Mixed / Other | - | 1.15 (0.88-1.50) | - | 0.55 (0.47-0.64) | - |
| BMI | Not obese | 1.00 (ref) | 1.00 (ref) | 1.00 (ref) | 1.00 (ref) | 1.00 (ref) |
| | Obese I (30-34.9) | 1.53 (1.41-1.66) | - | 1.14 (1.12-1.17) | 0.66 (0.59-0.74) | 1.00 (0.99-1.01) |
| | Obese II (35-39.9) | 1.88 (1.70-2.07) | - | 1.18 (1.15-1.22) | 0.62 (0.53-0.73) | 1.00 (0.99-1.01) |
| | Obese III (40+) | 2.58 (2.38-2.79) | - | 1.16 (1.12-1.20) | 0.69 (0.59-0.80) | 1.01 (0.99-1.02) |
| | Obese I (30-34.9) / Obese II (35-39.9) / Obese III (40+) | - | 2.75 (2.21-3.43) | - | - | - |
| History of | Chronic respiratory disease | - | 1.08 (0.78-1.50) | 0.89 (0.85-0.94) | 1.58 (1.39-1.80) | 0.89 (0.87-0.90) |
| | Chronic heart disease | 1.23 (1.14-1.32) | - | 1.04 (1.00-1.08) | 1.42 (1.28-1.58) | 1.06 (1.04-1.07) |
| | Chronic liver disease | 1.23 (1.12-1.35) | - | 0.96 (0.92-1.00) | 1.77 (1.57-1.99) | 1.06 (1.04-1.07) |
| | Chronic kidney disease | - | - | 1.06 (1.01-1.12) | 1.39 (1.20-1.61) | 1.11 (1.09-1.13) |
| | Chronic neurological disease | 1.32 (1.21-1.44) | - | 0.93 (0.90-0.97) | 1.22 (1.08-1.37) | 1.00 (0.98-1.01) |
| | Diabetes | 1.62 (1.50-1.75) | 1.68 (1.36-2.08) | 1.08 (1.04-1.12) | 1.05 (0.94-1.18) | 0.94 (0.93-0.96) |
| | Immunosuppression | - | - | 1.05 (1.00-1.09) | 0.84 (0.73-0.98) | 1.10 (1.08-1.12) |
| | Learning disability | - | - | 0.55 (0.52-0.59) | - | 0.92 (0.89-0.94) |
| | Serious mental illness | - | - | 0.56 (0.53-0.59) | 1.17 (0.98-1.39) | 0.78 (0.76-0.79) |
| Shielding criteria met | yes | 1.09 (0.98-1.20) | - | 0.89 (0.86-0.92) | 2.12 (1.81-2.50) | 0.91 (0.90-0.93) |
| Morbidity count | 0 | 1.00 (ref) | 1.00 (ref) | 1.00 (ref) | 1.00 (ref) | 1.00 (ref) |
| | 1 | - | - | 0.94 (0.89-0.99) | 0.72 (0.58-0.89) | 0.97 (0.95-0.99) |
| | 2+ | - | - | 0.89 (0.82-0.97) | 1.11 (0.86-1.43) | 0.91 (0.88-0.94) |
| | 1 / 2+ | 1.30 (1.10-1.55) | 2.31 (1.18-4.51) | - | - | - |
| Flu vaccine in previous 5 years | yes | 1.21 (1.13-1.30) | - | 1.02 (1.01-1.04) | 0.92 (0.83-1.02) | 1.23 (1.22-1.24) |
| Resident in long-term residential home | yes | - | - | - | - | 2.42 (2.31-2.53) |
| Housebound | yes | - | - | 0.75 (0.67-0.84) | 3.71 (3.16-4.36) | 0.87 (0.83-0.90) |
| Number of SARS-CoV-2 tests | 0 | 1.00 (ref) | 1.00 (ref) | 1.00 (ref) | 1.00 (ref) | 1.00 (ref) |
| | 1 | 1.39 (1.28-1.50) | - | 1.40 (1.38-1.43) | 1.15 (1.02-1.30) | 1.44 (1.42-1.45) |
| | 2 | - | - | 1.49 (1.44-1.53) | 1.72 (1.47-2.00) | 1.73 (1.71-1.75) |
| | 3+ | - | - | 1.59 (1.55-1.64) | 4.35 (3.91-4.84) | 2.64 (2.60-2.67) |
| | 2 / 3+ | 1.86 (1.72-2.02) | - | - | - | - |
| | 1 / 2 / 3+ | - | 1.27 (1.02-1.58) | - | - | - |
| Pregnancy | yes | - | - | 1.10 (1.03-1.17) | - | 1.76 (1.72-1.81) |

[a] Categories separated by " / " were merged due to low event counts. [b] Age terms were fit separately within strata. The age range in the strata is indicated by the suffix following the underscore on the age category label. Strata in which the age range was < 5 years included only a linear age term, otherwise a quadratic age term was included.

**Supplementary Table 14:** Hazard ratios corresponding to covariates in the adjusted BNT162b2 vs ChAdOx1 model in the 16-64 years and clinically vulnerable subgroup.

| Variable | Category[b] | COVID-19 hospitalisation | COVID-19 death | Positive SARS-CoV-2 test | Non-COVID-19 death | Any SARS-CoV-2 test |
|---|---|---|---|---|---|---|
| Age[a] | age_16to64 | 1.03 (1.00-1.06) | 1.07 (0.91-1.25) | 1.06 (1.05-1.06) | 1.09 (1.04-1.14) | 1.03 (1.03-1.03) |
| | age_16to64_squared | 1.00 (1.00-1.00) | 1.00 (1.00-1.00) | 1.00 (1.00-1.00) | 1.00 (1.00-1.00) | 1.00 (1.00-1.00) |
| Sex | Female | 1.00 (ref) | - | 1.00 (ref) | 1.00 (ref) | 1.00 (ref) |
| | Male | 1.24 (1.14-1.35) | - | 0.93 (0.92-0.95) | 1.43 (1.32-1.55) | 0.77 (0.76-0.77) |
| IMD | 1 most deprived | 1.00 (ref) | 1.00 (ref) | 1.00 (ref) | 1.00 (ref) | 1.00 (ref) |
| | 2 | - | - | 1.06 (1.04-1.09) | 0.95 (0.85-1.07) | 1.15 (1.13-1.16) |
| | 3 | - | - | 1.08 (1.05-1.10) | 0.86 (0.76-0.97) | 1.26 (1.24-1.27) |
| | 4 | - | - | 1.09 (1.07-1.12) | 0.89 (0.78-1.01) | 1.35 (1.34-1.37) |
| | 5 least deprived | - | - | 1.13 (1.10-1.16) | 0.85 (0.74-0.97) | 1.49 (1.47-1.50) |
| | 2 / 3 / 4 / 5 least deprived | 0.85 (0.77-0.94) | 1.12 (0.75-1.68) | - | - | - |
| Ethnicity | White | 1.00 (ref) | 1.00 (ref) | 1.00 (ref) | 1.00 (ref) | 1.00 (ref) |
| | Black | - | - | - | - | 0.90 (0.87-0.92) |
| | South Asian | - | - | - | - | 0.61 (0.60-0.62) |
| | Mixed | - | - | - | - | 0.88 (0.86-0.91) |
| | Other | - | - | - | - | 0.74 (0.72-0.76) |
| | Black / South Asian / Mixed / Other | 1.15 (1.00-1.33) | 2.07 (1.21-3.54) | 0.84 (0.81-0.86) | 0.60 (0.50-0.72) | - |
| BMI | Not obese | 1.00 (ref) | 1.00 (ref) | 1.00 (ref) | 1.00 (ref) | 1.00 (ref) |
| | Obese I (30-34.9) | - | - | 1.11 (1.09-1.14) | 0.65 (0.59-0.73) | 0.99 (0.98-1.00) |
| | Obese II (35-39.9) | - | - | 1.14 (1.11-1.18) | 0.62 (0.54-0.72) | 0.98 (0.97-0.99) |
| | Obese III (40+) | - | - | 1.13 (1.09-1.17) | 0.66 (0.57-0.77) | 1.00 (0.98-1.01) |
| | Obese I (30-34.9) / Obese II (35-39.9) / Obese III (40+) | 1.65 (1.51-1.79) | 2.24 (1.59-3.16) | - | - | - |
| History of | Chronic respiratory disease | - | - | 0.89 (0.85-0.93) | 1.71 (1.51-1.94) | 0.87 (0.86-0.89) |
| | Chronic heart disease | - | 1.67 (1.17-2.38) | 1.03 (0.99-1.07) | 1.40 (1.27-1.54) | 1.04 (1.03-1.05) |
| | Chronic liver disease | - | - | 0.97 (0.93-1.00) | 1.70 (1.51-1.91) | 1.04 (1.03-1.06) |
| | Chronic kidney disease | - | - | 1.03 (0.99-1.08) | 1.40 (1.22-1.60) | 1.10 (1.08-1.12) |
| | Chronic neurological disease | - | - | 0.92 (0.89-0.95) | 1.19 (1.06-1.34) | 0.98 (0.97-1.00) |
| | Diabetes | - | - | 1.05 (1.01-1.09) | 1.08 (0.97-1.20) | 0.95 (0.93-0.96) |
| | Immunosuppression | - | 1.51 (1.01-2.28) | 1.05 (1.01-1.09) | 0.82 (0.71-0.94) | 1.09 (1.08-1.11) |
| | Learning disability | - | - | 0.53 (0.50-0.57) | - | 0.96 (0.94-0.98) |
| | Serious mental illness | - | - | 0.57 (0.54-0.60) | 1.15 (0.94-1.39) | 0.74 (0.73-0.76) |
| Shielding criteria met | yes | 1.71 (1.49-1.98) | - | 0.90 (0.87-0.93) | 2.47 (2.12-2.88) | 0.91 (0.89-0.92) |
| Morbidity count | 0 | 1.00 (ref) | 1.00 (ref) | 1.00 (ref) | 1.00 (ref) | 1.00 (ref) |
| | 1 | - | - | 0.94 (0.89-0.99) | 0.62 (0.51-0.75) | 0.98 (0.96-1.00) |
| | 2+ | - | - | 0.91 (0.84-0.98) | 0.99 (0.78-1.26) | 0.92 (0.89-0.94) |
| | 1 / 2+ | 2.27 (1.82-2.83) | 4.22 (1.30-13.65) | - | - | - |
| Flu vaccine in previous 5 years | yes | - | - | 1.05 (1.03-1.07) | 0.87 (0.78-0.96) | 1.26 (1.25-1.27) |
| Resident in long-term residential home | yes | - | - | - | - | 2.38 (2.27-2.49) |
| Housebound | yes | - | - | - | 3.60 (3.05-4.26) | 0.87 (0.84-0.91) |
| Number of SARS-CoV-2 tests | 0 | 1.00 (ref) | 1.00 (ref) | 1.00 (ref) | 1.00 (ref) | 1.00 (ref) |
| | 1 | - | - | 1.32 (1.30-1.35) | 1.11 (0.99-1.24) | 1.39 (1.37-1.40) |
| | 2 | - | - | 1.40 (1.36-1.44) | 1.54 (1.32-1.78) | 1.68 (1.66-1.70) |
| | 3+ | - | - | 1.49 (1.44-1.54) | 4.03 (3.64-4.48) | 2.45 (2.42-2.49) |
| | 1 / 2 / 3+ | 1.77 (1.63-1.93) | 1.89 (1.36-2.63) | - | - | - |
| Pregnancy | yes | - | - | - | - | 1.03 (0.98-1.08) |

[a] Categories separated by " / " were merged due to low event counts. [b] Age terms were fit separately within strata. The age range in the strata is indicated by the suffix following the underscore on the age category label. Strata in which the age range was < 5 years included only a linear age term, otherwise a quadratic age term was included.

**Supplementary Table 15:** Hazard ratios corresponding to covariates in the adjusted BNT162b2 vs unvaccinated model in the 40-64 years and not clinically vulnerable subgroup.

| Variable | Category[b] | COVID-19 hospitalisation | COVID-19 death | Positive SARS-CoV-2 test | Non-COVID-19 death | Any SARS-CoV-2 test |
|---|---|---|---|---|---|---|
| Age[a] | age_40to45 | 1.08 (0.97-1.21) | - | 0.98 (0.97-0.99) | 1.22 (1.00-1.48) | 1.00 (0.99-1.00) |
| | age_45to50 | 0.89 (0.78-1.01) | - | 0.98 (0.97-1.00) | 1.18 (1.01-1.39) | 1.00 (0.99-1.01) |
| | age_50to55 | 1.01 (0.89-1.15) | - | 0.96 (0.94-0.98) | 1.04 (0.90-1.20) | 0.99 (0.98-1.00) |
| | age_55to60 | 0.97 (0.83-1.13) | - | 0.98 (0.96-1.00) | 1.04 (0.89-1.22) | 0.98 (0.97-0.99) |
| | age_60to64 | 0.87 (0.68-1.10) | - | 0.94 (0.90-0.98) | 1.12 (0.92-1.37) | 0.96 (0.95-0.98) |
| Sex | Female | - | - | 1.00 (ref) | - | 1.00 (ref) |
| | Male | - | - | 0.73 (0.72-0.75) | - | 0.71 (0.70-0.71) |
| IMD | 1 most deprived | 1.00 (ref) | - | 1.00 (ref) | 1.00 (ref) | 1.00 (ref) |
| | 2 | 1.04 (0.82-1.33) | - | 1.08 (1.05-1.12) | - | 1.13 (1.11-1.15) |
| | 3 | - | - | 1.12 (1.08-1.15) | - | 1.22 (1.20-1.24) |
| | 4 | - | - | 1.15 (1.12-1.19) | - | 1.32 (1.29-1.34) |
| | 5 least deprived | - | - | 1.18 (1.14-1.23) | - | 1.44 (1.41-1.47) |
| | 3 / 4 / 5 least deprived | 1.08 (0.87-1.33) | - | - | - | - |
| | 2 / 3 / 4 / 5 least deprived | - | - | - | 0.66 (0.53-0.83) | - |
| Ethnicity | White | 1.00 (ref) | - | 1.00 (ref) | 1.00 (ref) | 1.00 (ref) |
| | Black | 1.43 (1.06-1.94) | - | - | - | 0.91 (0.89-0.94) |
| | South Asian | 1.04 (0.77-1.40) | - | - | - | 0.57 (0.56-0.58) |
| | Mixed | 0.70 (0.38-1.30) | - | - | - | 0.85 (0.82-0.88) |
| | Other | 0.72 (0.44-1.17) | - | - | - | 0.63 (0.61-0.65) |
| | Black / South Asian / Mixed / Other | - | - | 0.67 (0.65-0.69) | 0.53 (0.39-0.72) | - |
| BMI | Not obese | 1.00 (ref) | - | 1.00 (ref) | 1.00 (ref) | 1.00 (ref) |
| | Obese I (30-34.9) | 2.53 (2.07-3.10) | - | 1.27 (1.23-1.30) | - | 1.09 (1.07-1.11) |
| | Obese II (35-39.9) | 3.89 (2.97-5.08) | - | - | - | 1.06 (1.03-1.09) |
| | Obese III (40+) | NA (NA-NA) | - | - | - | 0.83 (0.70-0.99) |
| | Obese II (35-39.9) / Obese III (40+) | - | - | 1.28 (1.23-1.34) | - | - |
| | Obese I (30-34.9) / Obese II (35-39.9) / Obese III (40+) | - | - | - | 0.70 (0.52-0.94) | - |
| History of | Chronic heart disease | - | - | - | - | 1.02 (0.86-1.22) |
| | Chronic liver disease | - | - | - | - | 0.94 (0.77-1.14) |
| | Chronic neurological disease | - | - | - | - | 0.93 (0.73-1.18) |
| | Diabetes | 1.89 (0.75-4.73) | - | - | - | 0.73 (0.61-0.87) |
| | Immunosuppression | - | - | - | - | 1.07 (0.86-1.33) |
| | Serious mental illness | - | - | - | - | 0.60 (0.50-0.71) |
| Shielding criteria met | yes | - | - | - | - | 1.32 (1.18-1.48) |
| Morbidity count | 0 | 1.00 (ref) | - | 1.00 (ref) | 1.00 (ref) | 1.00 (ref) |
| | 1 | - | - | - | - | 1.47 (1.26-1.71) |
| | 2+ | - | - | - | - | 2.30 (1.64-3.23) |
| | 1 / 2+ | 1.86 (1.02-3.36) | - | 1.12 (1.03-1.21) | 8.93 (6.41-12.44) | - |
| Flu vaccine in previous 5 years | yes | - | - | 1.04 (1.00-1.08) | - | 1.22 (1.20-1.25) |
| Number of SARS-CoV-2 tests | 0 | 1.00 (ref) | - | 1.00 (ref) | 1.00 (ref) | 1.00 (ref) |
| | 1 | - | - | 2.11 (2.05-2.17) | - | 2.06 (2.03-2.10) |
| | 2 | - | - | 2.38 (2.28-2.49) | - | 2.50 (2.45-2.56) |
| | 3+ | - | - | 2.65 (2.55-2.76) | - | 4.81 (4.71-4.91) |
| | 1 / 2 / 3+ | 1.41 (1.14-1.75) | - | - | 1.73 (1.35-2.23) | - |
| Pregnancy | yes | - | - | - | - | 1.89 (1.77-2.02) |

[a] Categories separated by ” / ” were merged due to low event counts. [b] Age terms were fit separately within strata. The age range in the strata is indicated by the suffix following the underscore on the age category label. Strata in which the age range was < 5 years included only a linear age term, otherwise a quadratic age term was included.

**Supplementary Table 16:** Hazard ratios corresponding to covariates in the adjusted ChAdOx1 vs unvaccinated model in the 40-64 years and not clinically vulnerable subgroup.

| Variable | Category[b] | COVID-19 hospitalisation | COVID-19 death | Positive SARS-CoV-2 test | Non-COVID-19 death | Any SARS-CoV-2 test |
|---|---|---|---|---|---|---|
| Age[a] | age_40to45 | 1.04 (1.00-1.09) | 1.34 (0.93-1.95) | 0.99 (0.98-0.99) | 1.19 (1.00-1.41) | 1.00 (0.99-1.00) |
| | age_45to50 | 1.03 (0.98-1.08) | 0.94 (0.75-1.16) | 0.96 (0.96-0.97) | 1.16 (1.03-1.32) | 0.99 (0.98-0.99) |
| | age_50to55 | 1.04 (1.00-1.09) | 0.94 (0.76-1.16) | 0.95 (0.94-0.95) | 1.09 (1.00-1.19) | 0.98 (0.97-0.98) |
| | age_55to60 | 1.02 (0.98-1.07) | 1.08 (0.88-1.32) | 0.96 (0.95-0.96) | 1.07 (0.99-1.15) | 0.99 (0.99-0.99) |
| | age_60to64 | 0.99 (0.92-1.07) | 1.21 (0.94-1.56) | 0.95 (0.93-0.96) | 1.09 (0.99-1.20) | 0.97 (0.96-0.97) |
| Sex | Female | 1.00 (ref) | - | 1.00 (ref) | 1.00 (ref) | 1.00 (ref) |
| | Male | 1.32 (1.23-1.41) | - | 0.91 (0.91-0.92) | 1.58 (1.39-1.79) | 0.80 (0.79-0.80) |
| IMD | 1 most deprived | 1.00 (ref) | 1.00 (ref) | 1.00 (ref) | 1.00 (ref) | 1.00 (ref) |
| | 2 | 0.97 (0.89-1.07) | - | 1.09 (1.07-1.11) | 0.73 (0.61-0.87) | 1.16 (1.15-1.17) |
| | 3 | 0.91 (0.82-1.01) | - | 1.12 (1.10-1.14) | 0.60 (0.50-0.72) | 1.29 (1.27-1.30) |
| | 4 | 0.90 (0.81-1.00) | - | 1.18 (1.16-1.20) | - | 1.40 (1.39-1.42) |
| | 5 least deprived | 0.92 (0.82-1.03) | - | 1.25 (1.23-1.27) | - | 1.53 (1.52-1.55) |
| | 2 / 3 / 4 / 5 least deprived | - | 0.77 (0.54-1.11) | - | - | - |
| | 4 / 5 least deprived | - | - | - | 0.47 (0.40-0.56) | - |
| Ethnicity | White | 1.00 (ref) | 1.00 (ref) | 1.00 (ref) | 1.00 (ref) | 1.00 (ref) |
| | Black | - | - | 0.77 (0.74-0.80) | - | 0.84 (0.82-0.85) |
| | South Asian | - | - | 0.66 (0.64-0.68) | - | 0.57 (0.56-0.57) |
| | Mixed | - | - | 0.82 (0.78-0.86) | - | 0.83 (0.81-0.85) |
| | Other | - | - | 0.59 (0.57-0.62) | - | 0.64 (0.63-0.66) |
| | Black / South Asian / Mixed / Other | 1.18 (1.09-1.28) | 1.00 (0.64-1.55) | - | 0.53 (0.42-0.68) | - |
| BMI | Not obese | 1.00 (ref) | 1.00 (ref) | 1.00 (ref) | 1.00 (ref) | 1.00 (ref) |
| | Obese I (30-34.9) | 2.22 (2.06-2.40) | - | 1.15 (1.14-1.17) | - | 0.98 (0.98-0.99) |
| | Obese II (35-39.9) | - | - | 1.19 (1.16-1.22) | - | 0.97 (0.96-0.98) |
| | Obese III (40+) | - | - | 1.34 (1.19-1.51) | - | 1.57 (1.13-2.18) |
| | Obese II (35-39.9) / Obese III (40+) | 3.28 (2.96-3.63) | - | - | - | - |
| | Obese I (30-34.9) / Obese II (35-39.9) / Obese III (40+) | - | 3.63 (2.65-4.98) | - | 0.74 (0.63-0.86) | - |
| History of | Chronic respiratory disease | - | - | - | - | 1.76 (1.26-2.46) |
| | Chronic heart disease | - | - | 1.14 (1.01-1.30) | - | 1.95 (1.41-2.71) |
| | Chronic liver disease | - | - | 1.17 (1.00-1.36) | - | 1.85 (1.33-2.57) |
| | Chronic kidney disease | - | - | - | - | 1.53 (1.08-2.17) |
| | Chronic neurological disease | - | - | - | - | 1.65 (1.17-2.31) |
| | Diabetes | - | - | 1.22 (1.08-1.37) | - | 1.60 (1.15-2.21) |
| | Immunosuppression | - | - | - | - | 1.94 (1.39-2.71) |
| | Serious mental illness | - | - | 0.81 (0.69-0.95) | - | 1.23 (0.89-1.72) |
| Shielding criteria met | yes | - | - | 0.94 (0.81-1.09) | - | 1.33 (1.25-1.41) |
| Morbidity count | 0 | 1.00 (ref) | 1.00 (ref) | 1.00 (ref) | 1.00 (ref) | 1.00 (ref) |
| | 1 | - | - | - | - | 0.59 (0.43-0.82) |
| | 2+ | - | - | - | - | 0.36 (0.18-0.70) |
| | 1 / 2+ | 2.09 (1.78-2.46) | 2.32 (1.18-4.57) | 0.92 (0.84-1.01) | 10.38 (8.73-12.34) | - |
| Flu vaccine in previous 5 years | yes | 1.20 (1.08-1.32) | - | 1.08 (1.06-1.09) | 0.89 (0.77-1.02) | 1.31 (1.30-1.32) |
| Resident in long-term residential home | yes | - | - | - | - | 1.47 (1.18-1.84) |
| Housebound | yes | - | - | - | - | 0.60 (0.51-0.71) |
| Number of SARS-CoV-2 tests | 0 | 1.00 (ref) | 1.00 (ref) | 1.00 (ref) | 1.00 (ref) | 1.00 (ref) |
| | 1 | 1.55 (1.41-1.70) | - | 1.42 (1.40-1.44) | - | 1.40 (1.39-1.40) |
| | 2 | 1.61 (1.38-1.88) | - | 1.57 (1.54-1.61) | - | 1.71 (1.69-1.73) |
| | 3+ | 1.82 (1.59-2.10) | - | 1.99 (1.96-2.03) | - | 3.51 (3.48-3.55) |
| | 1 / 2 / 3+ | - | 1.39 (0.94-2.05) | - | 1.22 (1.06-1.41) | - |
| Pregnancy | yes | - | - | 1.41 (1.23-1.61) | - | 1.79 (1.67-1.91) |

[a] Categories separated by ” / ” were merged due to low event counts. [b] Age terms were fit separately within strata. The age range in the strata is indicated by the suffix following the underscore on the age category label. Strata in which the age range was < 5 years included only a linear age term, otherwise a quadratic age term was included.

**Supplementary Table 17:** Hazard ratios corresponding to covariates in the adjusted BNT162b2 vs ChAdOx1 model in the 40-64 years and not clinically vulnerable subgroup.

| Variable | Category[b] | COVID-19 hospitalisation | COVID-19 death | Positive SARS-CoV-2 test | Non-COVID-19 death | Any SARS-CoV-2 test |
|---|---|---|---|---|---|---|
| Age[a] | age_40to45 | 1.10 (0.84-1.42) | - | 0.98 (0.98-0.99) | 1.18 (0.90-1.54) | 1.00 (0.99-1.00) |
| | age_45to50 | 0.90 (0.70-1.16) | - | 0.95 (0.94-0.96) | 1.16 (0.94-1.42) | 0.98 (0.98-0.99) |
| | age_50to55 | 1.10 (0.85-1.43) | - | 0.94 (0.94-0.95) | 1.11 (0.98-1.26) | 0.98 (0.97-0.98) |
| | age_55to60 | 0.87 (0.73-1.04) | - | 0.96 (0.95-0.96) | 1.07 (0.97-1.17) | 0.99 (0.99-0.99) |
| | age_60to64 | 0.96 (0.74-1.25) | - | 0.95 (0.94-0.96) | 1.08 (0.96-1.22) | 0.97 (0.96-0.97) |
| Sex | Female | - | - | 1.00 (ref) | - | 1.00 (ref) |
| | Male | - | - | 0.98 (0.97-0.99) | - | 0.81 (0.80-0.81) |
| IMD | 1 most deprived | 1.00 (ref) | - | 1.00 (ref) | 1.00 (ref) | 1.00 (ref) |
| | 2 | 0.60 (0.38-0.96) | - | 1.09 (1.07-1.12) | - | 1.18 (1.16-1.19) |
| | 3 | - | - | 1.13 (1.11-1.16) | - | 1.32 (1.30-1.33) |
| | 4 | - | - | 1.19 (1.17-1.22) | - | 1.44 (1.43-1.46) |
| | 5 least deprived | - | - | 1.28 (1.25-1.30) | - | 1.58 (1.56-1.60) |
| | 3 / 4 / 5 least deprived | 0.53 (0.36-0.78) | - | - | - | - |
| | 2 / 3 / 4 / 5 least deprived | - | - | - | 0.50 (0.41-0.61) | - |
| Ethnicity | White | 1.00 (ref) | - | 1.00 (ref) | 1.00 (ref) | 1.00 (ref) |
| | Black | - | - | - | - | 0.76 (0.74-0.78) |
| | South Asian | - | - | - | - | 0.57 (0.56-0.58) |
| | Mixed | - | - | - | - | 0.83 (0.80-0.85) |
| | Other | - | - | - | - | 0.67 (0.65-0.68) |
| | Black / South Asian / Mixed / Other | 1.95 (1.21-3.16) | - | 0.71 (0.69-0.73) | 0.56 (0.36-0.85) | - |
| BMI | Not obese | 1.00 (ref) | - | 1.00 (ref) | 1.00 (ref) | 1.00 (ref) |
| | Obese I (30-34.9) | 2.38 (1.70-3.33) | - | 1.11 (1.09-1.13) | - | 0.97 (0.97-0.98) |
| | Obese II (35-39.9) | - | - | - | - | 0.95 (0.94-0.96) |
| | Obese III (40+) | - | - | - | - | 0.95 (0.87-1.03) |
| | Obese II (35-39.9) / Obese III (40+) | 2.51 (1.52-4.14) | - | 1.16 (1.13-1.18) | - | - |
| | Obese I (30-34.9) / Obese II (35-39.9) / Obese III (40+) | - | - | - | 0.78 (0.64-0.95) | - |
| History of | Chronic heart disease | - | - | - | - | 1.15 (1.06-1.25) |
| | Chronic liver disease | - | - | - | - | 1.09 (1.00-1.20) |
| | Chronic neurological disease | - | - | - | - | 0.97 (0.87-1.08) |
| | Diabetes | NA (NA-NA) | - | - | - | 0.95 (0.88-1.03) |
| | Immunosuppression | - | - | - | - | 1.17 (1.05-1.30) |
| | Serious mental illness | - | - | - | - | 0.71 (0.64-0.78) |
| Shielding criteria met | yes | - | - | - | - | 1.28 (1.20-1.37) |
| Morbidity count | 0 | 1.00 (ref) | - | 1.00 (ref) | 1.00 (ref) | 1.00 (ref) |
| | 1 | - | - | - | - | 0.96 (0.90-1.03) |
| | 2+ | - | - | - | - | 0.98 (0.83-1.15) |
| | 1 / 2+ | 4.82 (2.60-8.93) | - | 0.96 (0.92-1.01) | 10.18 (8.04-12.89) | - |
| Flu vaccine in previous 5 years | yes | - | - | 1.11 (1.10-1.13) | - | 1.34 (1.33-1.34) |
| Number of SARS-CoV-2 tests | 0 | 1.00 (ref) | - | 1.00 (ref) | 1.00 (ref) | 1.00 (ref) |
| | 1 | - | - | 1.27 (1.26-1.29) | - | 1.33 (1.32-1.34) |
| | 2 | - | - | 1.39 (1.36-1.42) | - | 1.62 (1.61-1.64) |
| | 3+ | - | - | 1.85 (1.81-1.88) | - | 3.36 (3.33-3.39) |
| | 1 / 2 / 3+ | 1.97 (1.45-2.68) | - | - | 1.10 (0.92-1.31) | - |
| Pregnancy | yes | - | - | - | - | 1.02 (0.93-1.12) |

[a] Categories separated by ” / ” were merged due to low event counts. [b] Age terms were fit separately within strata. The age range in the strata is indicated by the suffix following the underscore on the age category label. Strata in which the age range was < 5 years included only a linear age term, otherwise a quadratic age term was included.

**Supplementary Table 18:** Hazard ratios corresponding to covariates in the adjusted BNT162b2 vs unvaccinated model in the 18-39 years and not clinically vulnerable subgroup.

| Variable | Category[b] | COVID-19 hospitalisation | COVID-19 death | Positive SARS-CoV-2 test | Non-COVID-19 death | Any SARS-CoV-2 test |
|---|---|---|---|---|---|---|
| Age[a] | age_18to25 | 1.09 (1.04-1.15) | - | 1.01 (1.00-1.01) | 0.92 (0.66-1.28) | 1.02 (1.01-1.02) |
| | age_25to30 | 1.09 (1.02-1.15) | - | 1.01 (1.00-1.02) | 0.63 (0.39-1.01) | 1.00 (1.00-1.00) |
| | age_30to36 | 1.04 (1.01-1.08) | - | 1.02 (1.01-1.02) | 1.27 (1.03-1.57) | 0.99 (0.99-1.00) |
| | age_36to40 | 1.08 (1.01-1.15) | - | 1.01 (1.00-1.02) | 1.04 (0.76-1.42) | 1.00 (0.99-1.00) |
| Sex | Female | 1.00 (ref) | - | 1.00 (ref) | - | 1.00 (ref) |
| | Male | 0.78 (0.71-0.86) | - | 0.74 (0.73-0.75) | - | 0.77 (0.77-0.78) |
| IMD | 1 most deprived | 1.00 (ref) | - | 1.00 (ref) | 1.00 (ref) | 1.00 (ref) |
| | 2 | - | - | 1.03 (1.01-1.05) | - | 1.07 (1.06-1.08) |
| | 3 | - | - | 1.04 (1.02-1.06) | - | 1.14 (1.13-1.15) |
| | 4 | - | - | 1.02 (1.00-1.04) | - | 1.19 (1.18-1.20) |
| | 5 least deprived | - | - | 1.03 (1.01-1.05) | - | 1.25 (1.24-1.27) |
| | 2 / 3 / 4 / 5 least deprived | 0.92 (0.84-1.00) | - | - | 0.76 (0.43-1.32) | - |
| Ethnicity | White | 1.00 (ref) | - | 1.00 (ref) | 1.00 (ref) | 1.00 (ref) |
| | Black / South Asian / Mixed / Other | 1.04 (0.95-1.15) | - | 0.61 (0.59-0.62) | 0.48 (0.23-1.01) | 0.73 (0.72-0.73) |
| BMI | Not obese | 1.00 (ref) | - | 1.00 (ref) | 1.00 (ref) | 1.00 (ref) |
| | Obese I (30-34.9) | - | - | 1.20 (1.17-1.22) | - | 1.05 (1.04-1.06) |
| | Obese II (35-39.9) / Obese III (40+) | - | - | 1.18 (1.14-1.22) | - | 1.04 (1.03-1.06) |
| | Obese I (30-34.9) / Obese II (35-39.9) / Obese III (40+) | 2.24 (2.04-2.47) | - | - | 1.40 (0.65-3.02) | - |
| Morbidity count | 0 | 1.00 (ref) | - | 1.00 (ref) | 1.00 (ref) | 1.00 (ref) |
| | 1 / 2+ | 1.15 (0.91-1.46) | - | 1.04 (0.98-1.10) | 13.86 (5.18-37.05) | 1.07 (1.04-1.10) |
| Flu vaccine in previous 5 years | yes | 1.00 (0.89-1.13) | - | 1.03 (1.01-1.05) | - | 0.98 (0.97-0.99) |
| Number of SARS-CoV-2 tests | 0 | 1.00 (ref) | - | 1.00 (ref) | 1.00 (ref) | 1.00 (ref) |
| | 1 | - | - | 1.85 (1.82-1.88) | - | 1.62 (1.61-1.64) |
| | 2 | - | - | 2.04 (2.00-2.09) | - | 1.94 (1.93-1.96) |
| | 3+ | - | - | 2.38 (2.33-2.42) | - | 3.18 (3.15-3.20) |
| | 1 / 2 / 3+ | 1.62 (1.49-1.77) | - | - | 1.17 (0.63-2.17) | - |
| Pregnancy | yes | 10.36 (9.29-11.56) | - | - | - | 1.97 (1.95-2.00) |

[a] Categories separated by " / " were merged due to low event counts. [b] Age terms were fit separately within strata. The age range in the strata is indicated by the suffix following the underscore on the age category label. Strata in which the age range was < 5 years included only a linear age term, otherwise a quadratic age term was included.
